# A Polygenic Risk Score for Coronary Artery Disease Improves the Prediction of Early-Onset Myocardial Infarction and Mortality in Men

**DOI:** 10.1101/2021.04.25.21256083

**Authors:** Hasanga D. Manikpurage, Aida Eslami, Nicolas Perrot, Zhonglin Li, Christian Couture, Patrick Mathieu, Yohan Bossé, Benoit J. Arsenault, Sébastien Thériault

## Abstract

**Background:** Several risk factors for coronary artery disease (CAD) have been described, some of which are genetically determined. The use of a polygenic risk score (PRS) could improve CAD risk assessment, but predictive accuracy according to age and sex is not well established.

**Methods:** A PRS_CAD_ including the weighted effects of >1.14 million SNPs associated with CAD was calculated in UK Biobank (n=408,422), using LDPred. Cox regressions were performed, stratified by age quartiles and sex, for incident MI and mortality, with a median follow-up of 11.0 years. Improvement in risk prediction of MI was assessed by comparing PRS_CAD_ to the pooled cohort equation with categorical net reclassification index using a 2% threshold (NRI^0.02^) and continuous NRI (NRI^>0^).

**Results:** From 7,746 incident MI cases and 393,725 controls, hazard ratio (HR) for MI reached 1.53 (95% CI [1.49-1.56], p=2.69e-296) per standard deviation (SD) increase of PRS_CAD_. PRS_CAD_ was significantly associated with MI in both sexes, with a stronger association in men (interaction p=0.002), particularly in those aged between 40-51 years (HR=2.00, 95% CI [1.86-2.16], p=1.93e-72). This group showed the highest reclassification improvement, mainly driven by the up-classification of cases (NRI^0.02^=0.199, 95% CI [0.157-0.248] and NRI^>0^=0.602, 95% CI [0.525-0.683]). From 23,982 deaths, HR for mortality was 1.08 (95% CI [1.06-1.09], p=5.46e-30) per SD increase of PRS_CAD_, with a stronger association in men (interaction p=1.60e-6).

**Conclusion:** Our PRS_CAD_ predicts MI incidence and all-cause mortality, especially in men aged between 40-51 years. PRS could optimize the identification and management of individuals at risk for CAD.

## INTRODUCTION

Coronary artery disease (CAD) is considered as a global health issue, responsible for a high morbidity and mortality worldwide, especially when presenting as a myocardial infarction (MI)^1^. CAD is heritable and has many recognized risk factors, several of which are genetically driven (e.g. body mass index, LDL-cholesterol, diabetes, blood pressure) while others are related to lifestyle or environment (e.g. smoking, nutritional behaviors, physical activity)^2^.

Current treatments for CAD include invasive procedures and preventive medical therapies. Numerous new molecules have been developed or are currently evaluated in clinical studies (e.g. PCSK9 inhibitors, lipoprotein(a) antisense)^3,4^ in order to reverse or slow down CAD progression through its causal risk factors. However, identifying and selecting patients who will benefit from these therapies remains challenging, especially considering the cost of these new treatments.

During the past decades, methods to identify high-risk patients have improved. From observational studies, several scores to quantify the risk of developing atherosclerotic cardiovascular disease (ASCVD) were proposed^5,6^. However, these scores do not consider genetic components and focus only on age-related risk factors or lifestyle habits observed at a given time point. Recent genome-wide association studies (GWAS), including data from up to >700 000 individuals, have uncovered numerous genetic loci associated with CAD^7,8^. These publicly available datasets enable the development of instruments that aggregate the effect of variants located throughout the genome. A polygenic risk score (PRS) can be estimated early in life and identify individuals with a high genetic risk. Recent algorithms, such as the one implemented in LDPred^9^, use Bayesian statistics to generate a posterior mean effect of genetic variants by accounting for nearby polymorphisms (i.e. in linkage disequilibrium), improving the predictive accuracy of the PRS.

To date, many PRS for CAD were proposed to establish the genetic risk profile of an individual^2,10–26^. While independently associated with CAD, the incremental value of these PRS to existing tools was shown to be relatively modest^18,19^. However, the thresholds to define individuals with a high polygenic risk have not been thoroughly explored in these studies. As shown previously, the use of a high PRS threshold could identify patients with risk similar to rare monogenic mutations associated with CAD^15,16^. The predictive accuracy of PRS for incident CAD has also not been reported according to age and sex strata.

Here, we evaluated the accuracy of a CAD PRS (PRS_CAD_) to predict incident MI and mortality in participants from UK Biobank followed for a median of 11.0 years. This study had three main objectives: (1) derive a PRS_CAD_ and evaluate its association with incident MI and mortality according to age and sex, (2) compare its predictive accuracy with other CAD risk factors and existing clinical tools and (3) propose a threshold to identify individuals at high risk who could benefit the most from early preventive measures.

We show that PRS_CAD_ is independently associated with incident MI and mortality. These associations are stronger in men aged between 40 and 51 years. We also suggest a threshold that could be used to identify individuals at high risk.

## METHODS

### Study Population and Genetic Data

UKB is linked to Hospital Episode Statistic (HES) data, as well as national death and cancer registries. These databases use normalized diagnostic codes from the International Classification of Diseases (ICD)-10^th^ revision and surgical procedure codes of OPSC-4 (Office of Population, Censuses and Surveys: Classification of Interventions and Procedures, version 4). To define prevalent CAD, we selected participants with ICD-10 codes for MI (I21.X, I22.X, I23.X, I24.1, or I25.2), for other acute ischemic heart disease (I24.0, I24.8-9) and for atherosclerotic / chronic ischemic heart disease (I25.0-25.1, I25.5-25.9). Procedure codes for coronary artery bypass grafting (K40.1–40.4, K41.1–41.4, K45.1–45.5), for coronary angioplasty, with or without stenting (K49.1–49.2, K49.8–49.9, K50.2, K75.1–75.4, K75.8–75.9) were also added to the CAD definition. We defined MI or revascularization based on MI ICD-10 codes, coronary artery bypass grafting and angioplasty procedure codes.

For incident analyses, we used first occurrences dates for MI (data-field 131298 – I21 ICD-10) provided by UKB until the end of June 2020. These data are retrieved from death registry, primary care information (available for ∼45% of UKB participants), hospital admission medical reports and self-report. Dates for all-cause mortality (data-field 40000) were provided until August 2020.

The estimation of 10-year risk of having a first ASCVD event was obtained by applying the pooled cohort equation (PCE)^5^. This equation combines the following common risk factors: ethnicity, sex, age, smoking status (current or not), HDL-cholesterol (HDL-C) levels, total cholesterol levels, diabetes status, systolic blood pressure and treatment against hypertension. Data collected or measured at the baseline assessment in UKB was used. In participants without direct HDL-C measurement, levels were estimated using the Friedewald equation for individuals with triglycerides levels <4.52 mmol/L (<400mg/dL). Otherwise, individuals with missing data for any risk factor were excluded (n =47,735).

Genetic data issued from UKB Phase 3 release were obtained from two genotyping arrays: Affymetrix UK Biobank Axiom and UKBiLEVE Axiom. Quality control procedures, phasing and imputation were centrally performed using UK10K panel, Haplotype Reference Consortium resource and 1000 Genomes Phase 3 panel as references^27^. Samples with a call rate <95%, an outlier heterozygosity rate, a sex mismatch, a non-white British ancestry, a third-degree relatives excess (>10), or not used for relatedness calculation, were excluded.

### Polygenic Risk Score for CAD

A polygenic score estimating the CAD genetic risk (PRS_CAD_) was built by using the LDPred software (v1.0.08)^9^. Our study population was divided into two groups: a training dataset of 5,000 randomly selected participants and a validation group including the rest of the cohort (n=403,422, Supplemental Table 1).

The LDPred algorithm uses a Bayesian approach to generate a posterior mean effect for each variant described in a GWAS, by using information on linkage disequilibrium (LD) of variants found in a reference panel. We used the summary statistics issued from the CAD GWAS meta-analysis of the CARDIoGRAMplusC4D^7^ consortium (including 60,801 cases and 123,504 controls). For the LD reference panel, we used data from the 503 European samples from the 1000 Genomes Project (phase 3, version 5)^28^.

Variants with info score R2 < 0.3 (i.e. quality of imputation), minor allele frequency (MAF) < 0.01, multi-allelic, insertion or deletions and with MAF discrepancies > 10% between all datasets were excluded. Only SNPs included in the Phase 3 HapMap Consortium (∼1.2 million) were kept for the PRS derivation as recommended^9^. This computational algorithm also considers the fraction of causal variants via a tuning parameter called rho, which is unknown for any given trait. A range of rho values (0.1 – 0.03 – 0.01 – 0.003 – 0.001 – 0.0003) were explored using the training dataset as recommended^9^.

The final PRS_CAD_ aggregates the re-weighted effect of 1,145,511 SNPs. From here, the PRS_CAD_ was z-score normalized for further statistical analysis.

### Statistical Analysis

To evaluate the association between PRS_CAD_ and prevalent CAD and MI, logistic regressions adjusted for age, sex and the first ten genetic principal components (PCs) were performed. We also tested the independence between the PRS_CAD_ and other CAD-related risk factors, namely LDL-cholesterol, body mass index, systolic blood pressure, diabetes, smoking status and the derived PCE variable.

Cox proportional hazard regression models were performed to evaluate the association of PRS_CAD_ with the incidence of a first MI and all-cause mortality. Dates of recruitment were used as the starting time and all participants with events occurring before recruitment were excluded. As data collection timeline differs for each outcome, we selected dates for the end of follow-up according to the frequency distribution in time (Figure 1 and Supplemental Figures 1-2). For incident MI, the end of follow-up date was selected within the last two years of the follow-up; each month event frequency was screened and if there was a decrease exceeding 60% of the mean of the previous three-month event frequency, the last day of the three-month period was chosen as the end date (Supplemental Figure 1). For all-cause mortality, we used the last day of February 2020 as the end of follow-up date, to avoid the subsequent sharp increase in mortality likely secondary to the COVID-19 pandemic (Supplemental Figure 2). Schoenfeld tests were performed as a prior to Cox regressions in order to verify proportionality assumption and Schoenfeld residuals were visually inspected. We stratified our analysis by quartiles of age and by sex. Models were adjusted for age, sex and the first ten genetic PCs in age-stratified analysis and only for age and the first ten genetic PCs in sex-stratified analyses. Models including the derived PCE variable were also tested.

**Figure 1:**
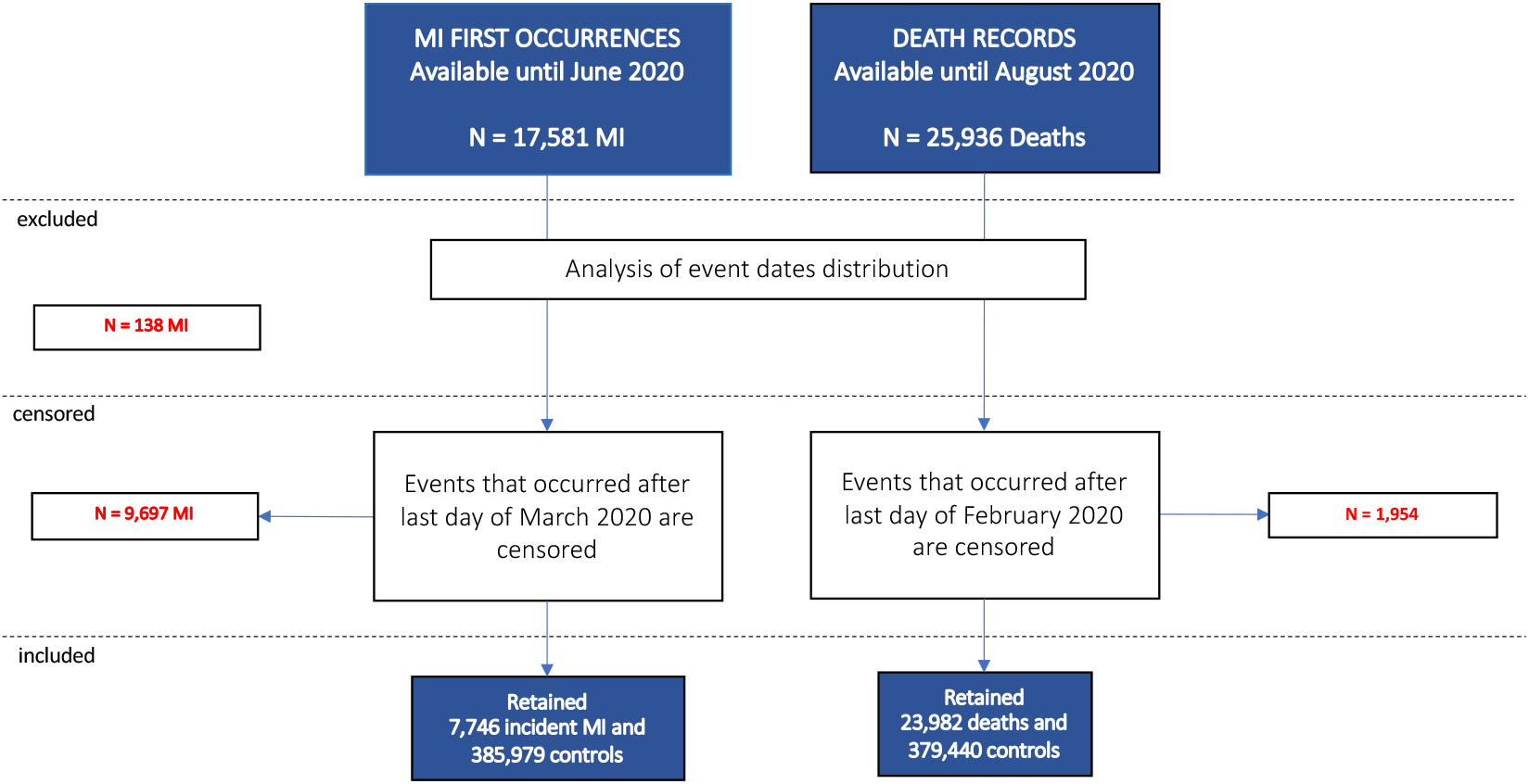
Study design for incident analyses in the validation group. Overview of participants’ selection for incident Myocardial Infarction (MI) and all-cause mortality.

To identify thresholds for a high genetic risk, we compared the individuals above the 80^th^, 90^th^, 95^th^, 97.5^th^ and 99^th^ percentile of the PRS distribution to the rest of the validation group.

To explore survival and incidence of event probabilities, we performed Kaplan-Meier analysis. Tertiles of PRS_CAD_ were used to define genetic risk categories, as well as individuals below and above the 90^th^ percentile of PRS_CAD_. Statistical significance through genetic risk categories were assessed via log-rank tests.

The discrimination of the predictive models was assessed using Harrell’s C-statistic and 95% confidence intervals (CI) were calculated by bootstrapping. PRS_CAD_ predictive performance was evaluated when added to PCE, using categorized net reclassification improvement (NRI), with a risk probabilities threshold of 2% (NRI^0.02^) obtained with Kaplan-Meier estimates for a period of 10 years. The risk threshold was selected based on the proportion of incident MI in the cohort. Continuous NRI (NRI^>0^) were also calculated to assess the strength of the instrument as suggested by Pencina et al^29^ and Kerr et al^30^. Confidence intervals (95%) for NRI^0.02^ and NRI^>0^ were obtained from bootstrapping.

All statistical analyses were conducted using R (version 3.5.1) and GraphPad Prism 9. P-values <0.05 were considered as significant.

## RESULTS

### PRS_CAD_ derivation and validation in UKB

The complete dataset includes 408,422 participants from European ancestry, with a mean age of 56.9 ± 8.0 years at recruitment and 54.1% of women. From Hospital Episode Statistics data, we have identified 32,694 cases of CAD (8.00%), 14,924 cases of MI (3.65%) and 20,769 cases of MI or revascularization (5.08%) (Supplemental Table 1).

A PRS_CAD_ including 1,145,511 SNPs was calculated in the training dataset including 5,000 participants. The optimal rho value was 0.01 (Supplemental Table 2) with a corresponding odds ratio for CAD (OR_CAD_) per SD increase of PRS_CAD_ reaching 1.48 (95% CI [1.29 to 1.70], p = 3.45e-08). The PRS_CAD_ was calculated in the validation group, where it was normally distributed (Anderson-Darling Normality Test, p = 0.1468, Supplemental Figure 3).

In the validation group, including 32,475 CAD cases and 370,947 controls, the OR_CAD_ was 1.56 (95% CI, [1.54 to 1.58, p < 10e-300) per SD increase of PRS_CAD_ (Figure 2). The area under the receiver operating characteristic curve (AUC) was 0.766 for the model including PRS_CAD_, age, sex and the first ten genetic principal components (Supplemental Table 3). OR_CAD_ per SD increase of PRS_CAD_ was higher in men (1.62 – 95% CI [1.59 to 1.64], p <10e-300) compared to women (1.45 – 95% CI [1.42 to 1.48], p = 3.65e-255; p = 1.21e-18 for interaction) (Figure 2). Likewise, OR for MI (OR_MI_) and OR for MI-Revascularization (OR_MI-REVASC_) per SD increase of PRS_CAD_ were higher in men than women (p = 2.39e-06 and p = 0.003 for interaction) (Figure 2).

**Figure 2:**
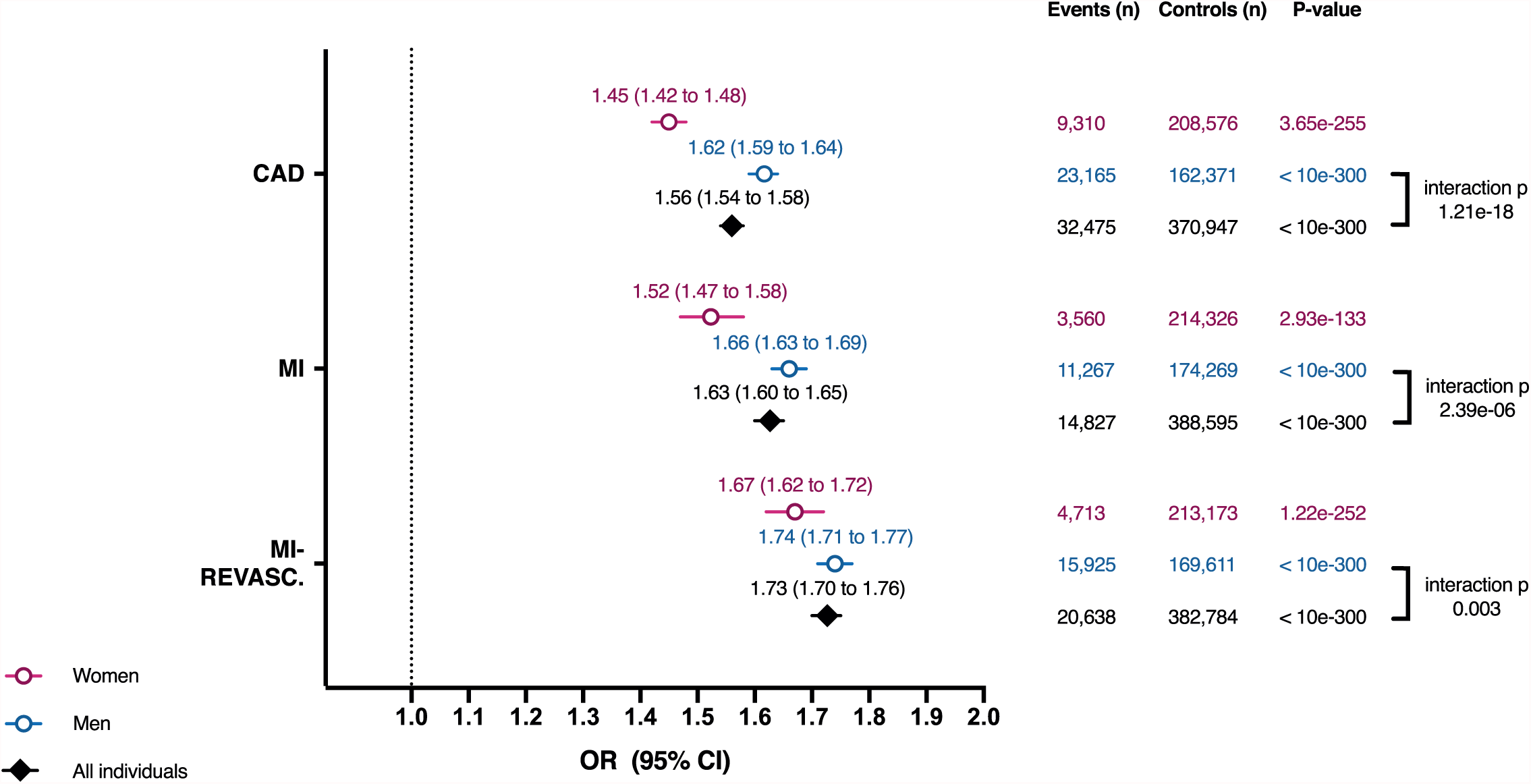
Association between PRS_CAD_ and prevalence of cardiovascular outcomes. Data are presented as estimated odds ratios (OR) per SD increase of PRS_CAD_ with their 95% confidence intervals (95% CI) and two-sided P-values from logistic regressions, adjusted for age, sex (only for all individuals) and the first ten genetic principal components. Significance of sex interaction terms is indicated. CAD: Coronary Artery Disease; MI: Myocardial Infarction; MI-REVASC: MI or Revascularisation Procedure.

The association between PRS_CAD_ and the cardiovascular outcomes was similar in models adjusted for body mass index, LDL-cholesterol, HDL-cholesterol, total cholesterol, systolic blood pressure, diabetes and smoking status (Supplemental Table 3). Similar results were obtained when adjusting for the PCE-derived 10-year risk of first ASCVD variable (Supplemental Table 3).

### PRS_CAD_ association with incidence of MI

To determine the predictive accuracy of PRS_CAD_, we evaluated its association with the incidence of a first MI. A total of 7,746 first MI occurred during the follow-up (median of 11.0 years) (Table 1). There was a significant decrease in PRS_CAD_ in the third and fourth age quartiles (Supplemental Figure 4). The risk of having a first MI was positively associated with PRS_CAD_. When including all age groups and both sexes, adjusted hazard ratios (HR) for MI (HR_MI_) per SD increase of PRS_CAD_ was 1.53 (95% CI [1.49 to 1.56], p = 2.69e-296) (Supplemental Table 4).

**Table 1.** Characteristics of the validation group, stratified by age and sex.

| Categories | Sub-Categories | Validation Group | Quartile 1 Group |  | Quartile 2 Group |  | Quartile 3 Group |  | Quartile 4 Group |  |
| --- | --- | --- | --- | --- | --- | --- | --- | --- | --- | --- |
|  |  | All | Women | Men | Women | Men | Women | Men | Women | Men |
| n (%) |  | 403,422 | 55,289 (54.82) | 45,567 (45.18) | 56,704 (56.22) | 44,151 (43.78) | 54,737 (54.27) | 46,118 (45.73) | 51,156 (50.72) | 49,700 (49.28) |
| Age, y (± SD) | | 56.93 (8.00) | 46.29 (3.07) | 46.16 (3.10) | 55.24 (2.24) | 55.26 (2.24) | 61.52 (1.43) | 61.54 (1.44) | 66.83 (1.79) | 66.79 (1.78) |
| BMI, kg/m<sup>2</sup> (± SD) | | 27.42 (4.76) | 26.49 (5.35) | 27.65 (4.32) | 27.11 (5.30) | 28.00 (4.42) | 27.32 (5.03) | 27.96 (4.23) | 27.32 (4.77) | 27.81 (3.97) |
| Blood Pressure, mmHg (± SD) | Diastolic | 82.27 (10.11) | 79.16 (10.03) | 83.75 (10.04) | 81.27 (9.94) | 85.23 (9.97) | 81.34 (9.77) | 84.56 (9.88) | 80.85 (9.83) | 83.23 (9.94) |
|  | Systolic | 138.30 (18.64) | 125.69 (15.71) | 135.08 (14.62) | 133.63 (17.89) | 139.85 (16.54) | 139.51 (18.61) | 143.38 (17.65) | 144.60 (19.30) | 146.58 (18.41) |
| Blood lipids, mmol/L (± SD) | LDL-C | 3.57 (0.87) | 3.34 (0.77) | 3.62 (0.82) | 3.72 (0.84) | 3.60 (0.85) | 3.80 (0.89) | 3.45 (0.87) | 3.73 (0.92) | 3.28 (0.87) |
|  | HDL-C | 1.45 (0.38) | 1.55 (0.37) | 1.25 (0.32) | 1.62 (0.40) | 1.27 (0.34) | 1.51 (0.40) | 1.28 (0.35) | 1.60 (0.40) | 1.27 (0.35) |
|  | TC | 5.71 (1.15) | 5.45 (0.97) | 5.65 (1.07) | 6.00 (1.08) | 5.64 (1.11) | 6.12 (1.14) | 5.46 (1.14) | 6.03 (1.19) | 5.24 (1.14) |
|  | APO-B | 1.07 (0.24) | 0.96 (0.22) | 1.05 (0.24) | 1.06 (0.23) | 1.06 (0.24) | 1.08 (0.24) | 1.02 (0.24) | 1.06 (0.25) | 0.98 (0.24) |
| nmol/L (± SD) | Lp(a) | 44.09 (49.45) | 43.11 (48.80) | 43.20 (49.49) | 44.91 (49.78) | 43.29 (49.28) | 45.16 (49.73) | 43.84 (49.41) | 45.50 (49.80) | 43.34 (49.19) |
| Hypertension, n (%) |  | 90,542 (22.44) | 5,832 (10.55) | 7,223 (15.85) | 12,136 (21.40) | 13,771 (31.19) | 17,771 (32.47) | 19,746 (42.82) | 22,795 (44.56) | 26,782 (53.89) |
| Diabetes, n (%) |  | 20,522 (5.09) | 1,914 (3.46) | 2,408 (5.28) | 2,955 (5.21) | 4,212 (9.54) | 3,721 (6.90) | 5,651 (12.25) | 4,691 (9.17) | 7,556 (15.20) |
| Current Smoker n (%) |  | 40,763 (10.10) | 6,398 (11.57) | 6,925 (15.20) | 5,329 (9.40) | 5,616 (12.72) | 3,998 (7.30) | 4,979 (10.80) | 3,138 (6.13) | 4,380 (8.81) |
| 10-year risk of first ASCVD event (PCE), % (± SD) | | 8.39 (7.66) | 1.31 (1.47) | 4.23 (3.55) | 2.97 (2.05) | 9.03 (4.72) | 5.70 (2.98) | 14.47 (5.97) | 10.45 (4.83) | 21.24 (7.51) |
| Mean PRS<sub>CAD</sub> (± SD) | | 6.8e-06 (1.00) | 0.0047 (1.00) | 0.0144 (1.00) | 0.0044 (1.01) | -0.002 (0.99) | -0.0015 (1.00) | -0.0038 (1.00) | -0.003 (1.00) | -0.013 (0.99) |
| Incident MI, n (%) |  | 7,746 (1.92) | 208 (0.38) | 681 (1.49) | 427 (0.75) | 1,175 (2.66) | 662 (1.20) | 1,562 (3.39) | 998 (1.95) | 2,033 (4.09) |
| All-cause Mortality, n (%) |  | 23,982 (5.94) | 830 (1.50) | 1,011 (2.21) | 1,649 (2.91) | 2,182 (4.94) | 2,651 (4.84) | 3,906 (8.47) | 4,474 (8.75) | 7,279 (14.65) |
| Median Follow-up, y (± IQR) | | 11.04 (1.40) | 11.13 (1.36) | 11.12 (1.39) | 11.12 (1.29) | 11.10 (1.33) | 11.05 (1.39) | 10.99 (1.44) | 10.98 (1.41) | 10.87 (1.56) |

Continuous variables are presented with mean values ± SD.
ApoB: Apolipoprotein B; ASCVD: Atherosclerotic cardiovascular disease; BMI: Body mass index; HDL-C: High density lipoprotein cholesterol; IQR: Interquartile range; LDL-C: Low density lipoprotein cholesterol; Lp(a): Lipoprotein (a); MI: Myocardial infarction; PCE: Pooled cohort equation; PRS<sub>CAD</sub>: Polygenic risk score for coronary artery disease; SD: Standard deviation; TC: Total cholesterol

In sex and age-stratified analyses, adjusted HR_MI_ for women (n = 2,295 MI) varied between 1.57 (quartile 1, 95% CI [1.37 to 1.79], p = 9.04e-11) and 1.40 (quartile 3, 95% CI [1.30 to 1.51], p = 7.90e-18) per SD increase of PRS_CAD_ (Figure 3A and Supplemental Table 4). In men (n = 5,451 MI), a stronger association was observed in the first quartile of age, with an adjusted HR_MI_ of 2.00 (95% CI [1.86 to 2.16], p = 1.93e-72) versus 1.48 (95% CI [1.41 to 1.54], p = 1.04e-66) for the last quartile, per SD increase of PRS_CAD_ (Figure 3B and Supplemental Table 4). Models adding the PCE-derived variable as a covariate did not show significant modification in HR_MI_ for both sexes (Supplemental Table 5). Overall, there was a significant sex interaction (p = 0.002). When stratifying by age, significant sex interactions were only observed in the first (p = 0.001) and second (p = 0.012) quartiles.

**Figure 3:**
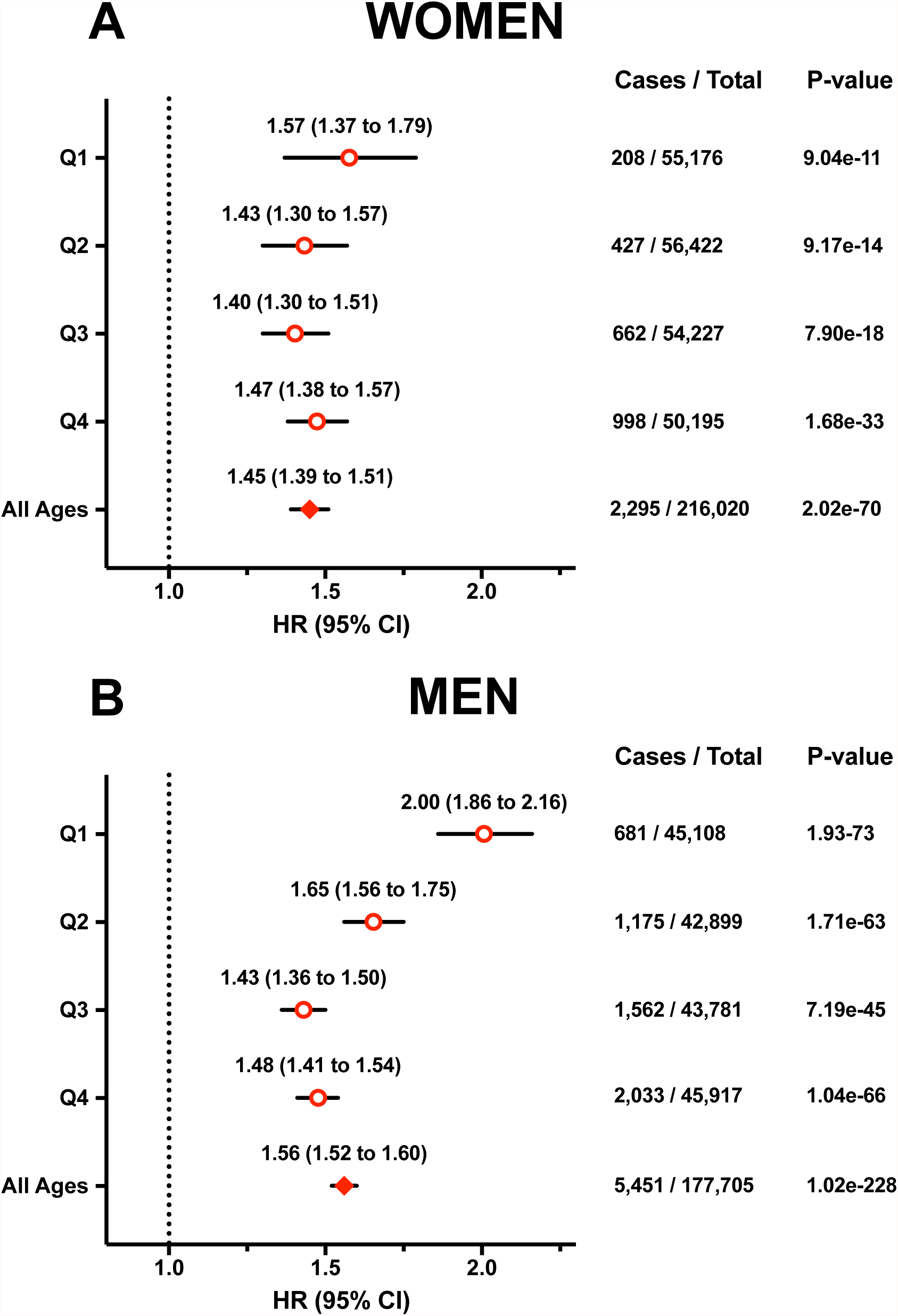
Association between PRS_CAD_ and MI incidence. Cox regression results were obtained from models adjusted for age and the first ten genetic principal components (PCs) in women (A) and in men (B), by quartile of age (Q1 to Q4) and for the whole group (all ages). Data are presented as estimated hazard ratios (HR) for incident MI per SD increase of PRS_CAD_, with 95% confidence intervals (95% CI).

There was an inverse correlation between PRS_CAD_ and age at first incident MI, with a Pearson’s R value of -0.094 (95% CI [-0.116 to -0.0719], p = 1.14e-16). The correlation was only significant in men (Pearson’s R value of -0.125 (95% CI [-0.151 to -0.0992], p = 1.44e-20; women: Pearson’s R value = -0.0162, 95% CI [-0.0571 to 0.0247], p = 0.44) (Supplemental Figure 5).

For each category stratified by both age and sex, we also verified the impact of being in the highest percentiles of the PRS_CAD_ distribution on HR_MI_. Overall, the risk of incident MI increased gradually across percentiles for all quartiles of age, although confidence intervals were wider for higher thresholds (Figure 4). The 90^th^ percentile threshold corresponded to a HR_MI_ above 2 in both sexes while higher thresholds showed modest increases in risk (Figure 4 and Supplemental Table 6). At the 90^th^ percentile threshold, the strongest associations were observed in the first quartile of age for both sexes, with a HR_MI_ of 2.78 (95% CI [2.01 to 3.82]) in women and 3.45 (95% CI [2.92 to 4.09]) in men (Figure 4 and Supplemental Table 6).

**Figure 4:**
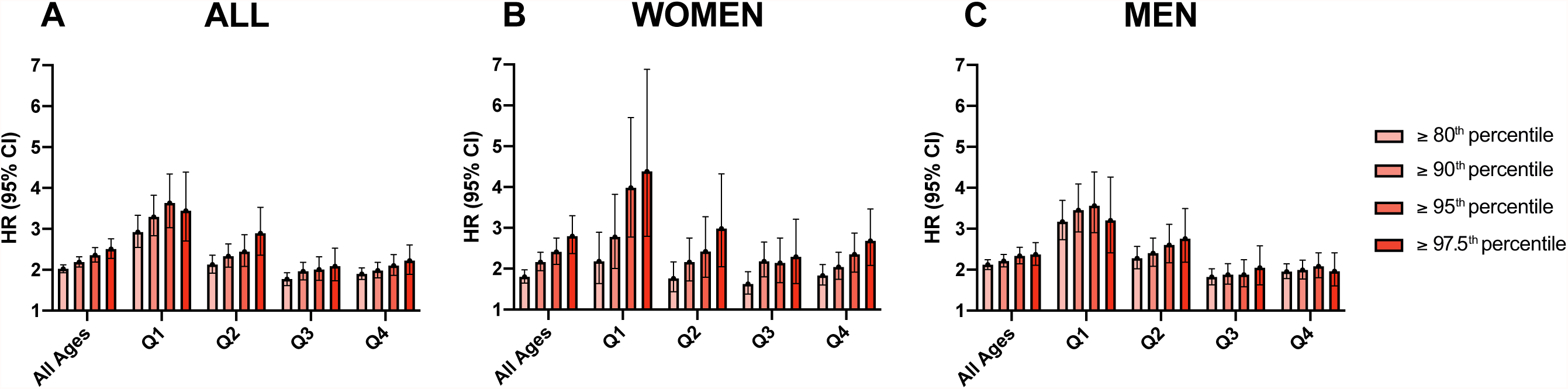
MI incidence in individuals at high genetic risk. Cox regression results were obtained for individuals above different percentile thresholds of PRS_CAD_ (ranging between the 80^th^ and the 97.5^th^ percentile). Hazard ratios (HR) for MI incidence with their 95% confidence intervals (95% CI) were compared in all individuals (A), and separately in women (B) and in men (C), for all ages and each quartile of age (Q1 to Q4).

Kaplan-Meier estimates of cumulative events for MI were consistent with Cox regression results. When divided in tertiles, PRS_CAD_ was significantly associated with a higher incidence, in all individuals (Figure 5A– Log-rank test, p <0.0001), but also in both women and men (respectively Figures 5B and 5C – Log-rank test, p <0.0001). Taken together, a wider separation was observed for cumulative events curves in men (Supplemental Figure 6). After 10 years of follow-up, estimates of cumulative events for MI ranged between 0.62 % (low PRS) and 1.31 % (high PRS) in women, and between 1.62 % (low PRS) and 4.21 % (high PRS) in men (Figures 5B and 5C). For individuals above the 90^th^ percentile of PRS_CAD_, estimates of cumulative events for MI reached 1.76% and 5.44% respectively for women and men (Supplemental Figure 7 – Log-rank test, p <0.0001).

**Figure 5:**
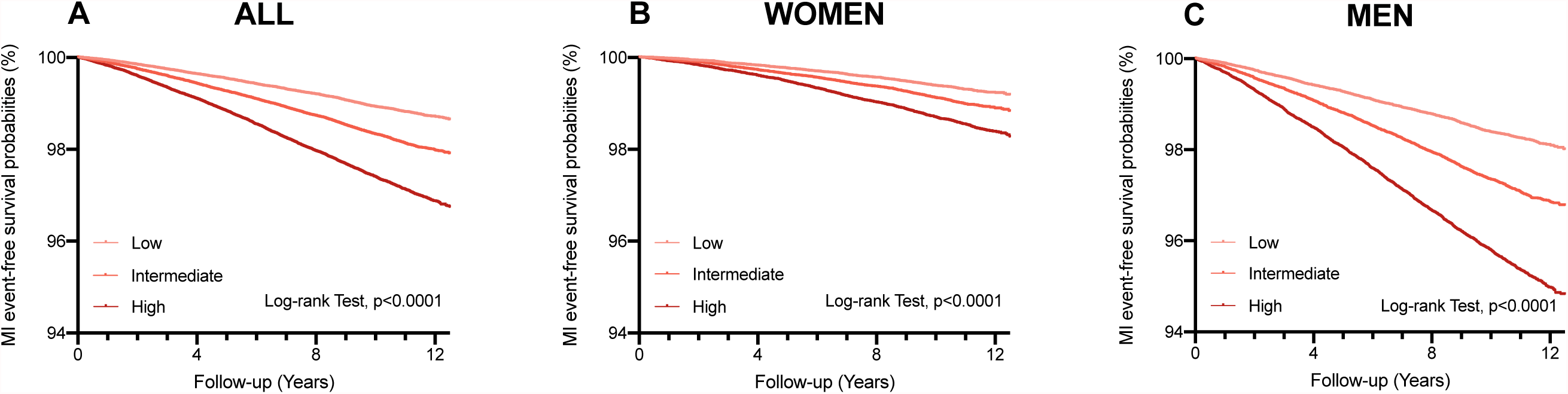
Event free probability for incident MI according to genetic risk. Event-free probability curves for MI were obtained from Kaplan-Meier estimates for PRS_CAD_ tertiles (low, intermediate, high) in all individuals (A) and separately in women (B) and men (C). Only individuals without an event before recruitment were included. The median follow-up was 11.0 years. Log-rank test, p-values <0.0001 for A, B and C.

Discrimination was assessed in Cox regression models including various covariables using Harrell’s C-Statistic (Table 2). The model including PCE-derived variable, PRS_CAD_ and the first ten genetic principal components (model 5) showed higher discrimination in all individuals (0.748 - 95% CI [0.743 to 0.754]) when compared to the model including PCE only (model 4) (0.740 - 95% CI [0.734 to 0.745]). Higher C-statistics were observed in model 5 for all age groups in men, ranging between 0.747 (quartile 1, 95% CI [0.731 to 0.768]) and 0.654 (quartile 4, 95% CI [0.643 to 0.669]), but only in quartile 4 in women (0.673 – 95% CI [0.657 to 0.694]).

**Table 2.** Discrimination using Harrell’s C-statistic in Cox regression models for MI Incidence.

| Sex Group | Age Group | Harrell's C-Statistic |  |  |  |  |
| --- | --- | --- | --- | --- | --- | --- |
|  |  | Model 1:<br>AGE + SEX | Model 2:<br>PRS<sub>CAD</sub> + PCs | Model 3:<br>PRS<sub>CAD</sub> + AGE + SEX + PCs | Model 4:<br>PCE | Model 5:<br>PRS<sub>CAD</sub> + PCE + PCs |
| All | (All ages) from 40 to 73 | 0.699 (0.723 to 0.734) | 0.613 (0.607 to 0.619) | 0.729 (0.724 to 0.735) | 0.740 (0.734 to 0.745) | 0.748 (0.743 to 0.754) |
|  | (Q1) from 40 to 51 | 0.701 (0.743 to 0.775) | 0.680 (0.664 to 0.701) | 0.760 (0.746 to 0.777) | 0.795 (0.782 to 0.810) | 0.748 (0.733 to 0.769) |
|  | (Q2) from 52 to 58 | 0.672 (0.703 to 0.728) | 0.625 (0.613 to 0.640) | 0.717 (0.704 to 0.730) | 0.745 (0.733 to 0.757) | 0.737 (0.726 to 0.752) |
|  | (Q3) from 59 to 63 | 0.643 (0.672 to 0.694) | 0.598 (0.588 to 0.611) | 0.684 (0.674 to 0.695) | 0.707 (0.696 to 0.718) | 0.714 (0.703 to 0.725) |
|  | (Q4) from 64 to 73 | 0.618 (0.653 to 0.673) | 0.607 (0.598 to 0.618) | 0.664 (0.656 to 0.675) | 0.671 (0.660 to 0.681) | 0.698 (0.689 to 0.708) |
| Women | (All ages) from 40 to 71 | 0.668 (0.658 to 0.678) | 0.601 (0.590 to 0.614) | 0.694 (0.685 to 0.706) | 0.728 (0.718 to 0.738) | 0.718 (0.708 to 0.733) |
|  | (Q1) from 40 to 51 | 0.604 (0.563 to 0.641) | 0.620 (0.601 to 0.672) | 0.655 (0.631 to 0.705) | 0.781 (0.750 to 0.815) | 0.689 (0.671 to 0.750) |
|  | (Q2) from 52 to 58 | 0.539 (0.511 to 0.566) | 0.601 (0.583 to 0.636) | 0.609 (0.592 to 0.645) | 0.700 (0.671 to 0.730) | 0.699 (0.678 to 0.728) |
|  | (Q3) from 59 to 63 | 0.514 (0.498 to 0.534) | 0.598 (0.583 to 0.624) | 0.599 (0.586 to 0.627) | 0.655 (0.632 to 0.676) | 0.652 (0.636 to 0.682) |
|  | (Q4) from 64 to 71 | 0.548 (0.529 to 0.566) | 0.611 (0.596 to 0.633) | 0.621 (0.605 to 0.641) | 0.637 (0.618 to 0.655) | 0.673 (0.657 to 0.694) |
| Men | (All ages) from 40 to 73 | 0.614 (0.607 to 0.621) | 0.622 (0.614 to 0.630) | 0.671 (0.665 to 0.678) | 0.675 (0.668 to 0.682) | 0.707 (0.700 to 0.714) |
|  | (Q1) from 40 to 51 | 0.582 (0.563 to 0.602) | 0.702 (0.686 to 0.724) | 0.718 (0.702 to 0.737) | 0.730 (0.710 to 0.749) | 0.747 (0.731 to 0.768) |
|  | (Q2) from 52 to 58 | 0.536 (0.519 to 0.552) | 0.640 (0.626 to 0.658) | 0.644 (0.631 to 0.661) | 0.661 (0.644 to 0.678) | 0.692 (0.679 to 0.709) |
|  | (Q3) from 59 to 63 | 0.523 (0.507 to 0.536) | 0.604 (0.592 to 0.620) | 0.608 (0.596 to 0.624) | 0.635 (0.621 to 0.650) | 0.657 (0.645 to 0.671) |
|  | (Q4) from 64 to 73 | 0.521 (0.509 to 0.535) | 0.611 (0.601 to 0.625) | 0.614 (0.604 to 0.629) | 0.608 (0.594 to 0.621) | 0.654 (0.643 to 0.669) |

95% Confidence intervals were obtained by bootstrapping
PRS<sub>CAD</sub>: Polygenic risk score for coronary artery disease; PCE: Pooled cohort equation; PCs: First ten genetic principal components

Categorized net reclassification improvement (NRI) was calculated using a threshold of 2% (NRI^0.02^) of predicted 10-year risk of MI. The addition of PRS_CAD_ to the Cox proportional hazard models including PCE resulted in a NRI^0.02^ of 0.0452 (95% CI, [0.0333 to 0.0573]), whereas continuous NRI (NRI^>0^) was equal to 0.324 (95% CI [0.297 to 0.349]) (Table 3 and Supplemental Table 7).

**Table 3.**
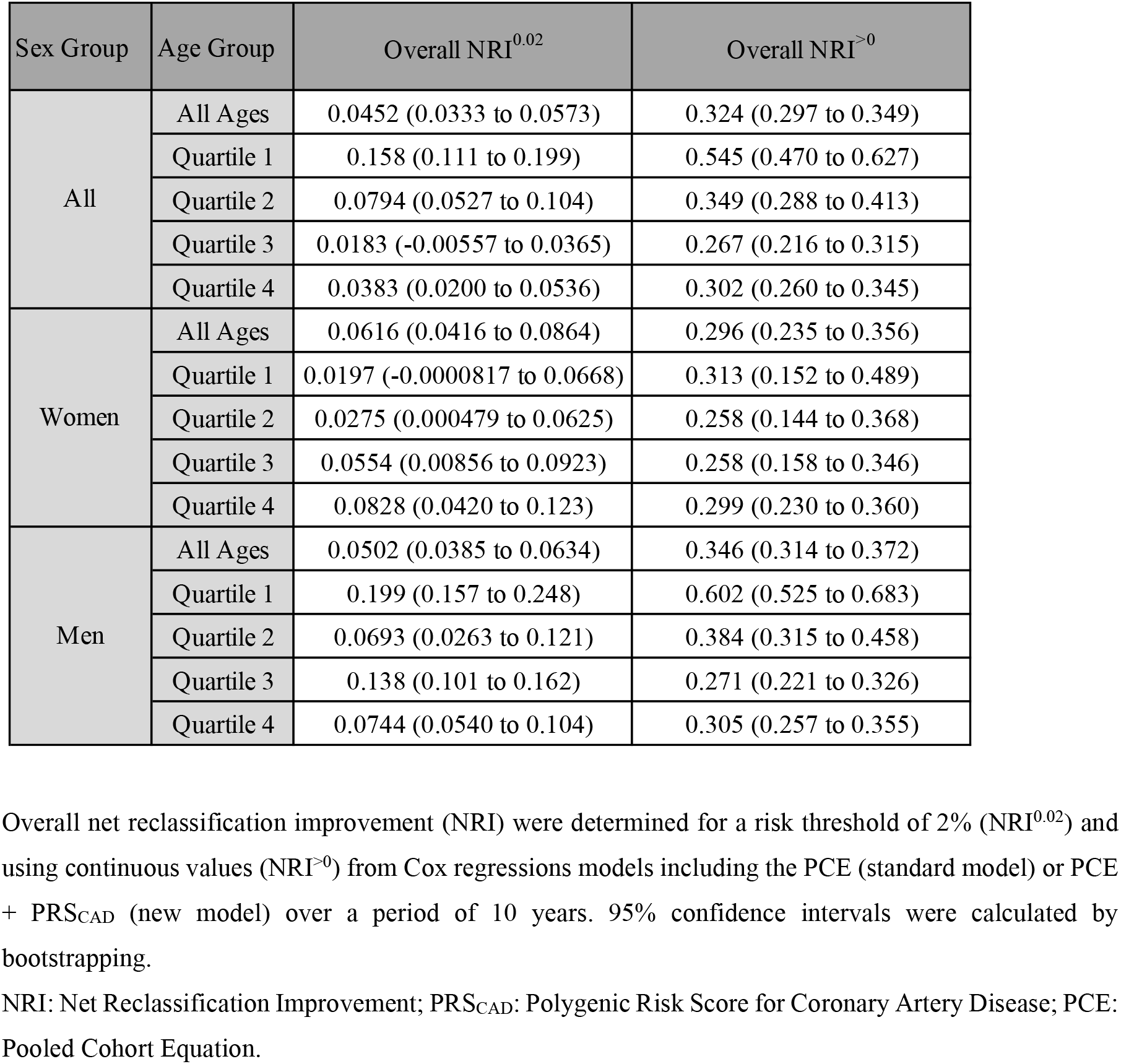
Net reclassification improvement of PRS_CAD_ for MI incidence.

| Sex Group | Age Group | Overall NRI<sup>0.02</sup> | Overall NRI<sup>>0</sup> |
| --- | --- | --- | --- |
| All | All Ages | 0.0452 (0.0333 to 0.0573) | 0.324 (0.297 to 0.349) |
|  | Quartile 1 | 0.158 (0.111 to 0.199) | 0.545 (0.470 to 0.627) |
|  | Quartile 2 | 0.0794 (0.0527 to 0.104) | 0.349 (0.288 to 0.413) |
|  | Quartile 3 | 0.0183 (-0.00557 to 0.0365) | 0.267 (0.216 to 0.315) |
|  | Quartile 4 | 0.0383 (0.0200 to 0.0536) | 0.302 (0.260 to 0.345) |
| Women | All Ages | 0.0616 (0.0416 to 0.0864) | 0.296 (0.235 to 0.356) |
|  | Quartile 1 | 0.0197 (-0.0000817 to 0.0668) | 0.313 (0.152 to 0.489) |
|  | Quartile 2 | 0.0275 (0.000479 to 0.0625) | 0.258 (0.144 to 0.368) |
|  | Quartile 3 | 0.0554 (0.00856 to 0.0923) | 0.258 (0.158 to 0.346) |
|  | Quartile 4 | 0.0828 (0.0420 to 0.123) | 0.299 (0.230 to 0.360) |
| Men | All Ages | 0.0502 (0.0385 to 0.0634) | 0.346 (0.314 to 0.372) |
|  | Quartile 1 | 0.199 (0.157 to 0.248) | 0.602 (0.525 to 0.683) |
|  | Quartile 2 | 0.0693 (0.0263 to 0.121) | 0.384 (0.315 to 0.458) |
|  | Quartile 3 | 0.138 (0.101 to 0.162) | 0.271 (0.221 to 0.326) |
|  | Quartile 4 | 0.0744 (0.0540 to 0.104) | 0.305 (0.257 to 0.355) |

Overall net reclassification improvement (NRI) were determined for a risk threshold of 2% (NRI<sup>0.02</sup>) and using continuous values (NRI<sup>>0</sup>) from Cox regressions models including the PCE (standard model) or PCE + PRS<sub>CAD</sub> (new model) over a period of 10 years. 95% confidence intervals were calculated by bootstrapping.
NRI: Net Reclassification Improvement; PRS<sub>CAD</sub>: Polygenic Risk Score for Coronary Artery Disease; PCE: Pooled Cohort Equation.

When sex-stratified, NRI^0.02^ reached 0.0616 (95% CI [0.0416 to 0.0864]) for women and 0.0502 (95% CI, [0.0385 to 0.0634]) for men, while NRI^>0^ reached respectively 0.296 (95% CI [0.235 to 0.356]) and 0.346 (95% CI 0.314 to 0.372) (Table 3 and Supplemental Table 7).

Age-stratified analysis showed different results in women and men. First, NRI^0.02^ was low in women when stratified by age, likely due to an overall smaller number of cases (Table 3, Supplemental Table 7 and Supplemental Figure 8). In men, NRI^0.02^ was higher in the first quartile of age (0.199 – 95% CI [0.157 to 0.248]), mainly driven by the reclassification of events in the high-risk category (Supplemental Figure 8B), and decreased over the other quartiles (Table 3 and Supplemental Table 7). NRI^>0^ showed at least modest improvement in all groups, ranging from 0.271 to 0.602 (Table 3 and Supplemental Table 7).

### PRS_CAD_ association with mortality

We have evaluated the impact of PRS_CAD_ on all-cause mortality. 23,982 deaths have occurred until March 2020 in the validation group. For all individuals, HR for mortality (HR_MORTALITY_) per SD increase of PRS_CAD_ reached 1.08 (95% CI [1.06 to 1.09], p = 5.46e-30) (Supplemental Table 8).

A sex interaction within all participants was observed, with a stronger association observed in men (p = 1.65e-06, Figure 6). In women (n=9,604 deaths), HR_MORTALITY_ per SD increase of PRS_CAD_ was significant (1.04 - 95% CI [1.02 to 1.06], p = 3.17e-04), but when stratified by age quartiles, only in the first and last were significant (Figure 6A and Supplemental Table 8). In men (n = 14,378 deaths), HR_MORTALITY_ per SD increase of PRS_CAD_ were significant for all age groups; the strongest association was observed in the youngest group (aged between 40 and 51 years) with a corresponding HR of 1.15 (95% CI [1.08 to 1.22], p = 1.64e-05) (Figure 6B and Supplemental Table 8).

**Figure 6:**
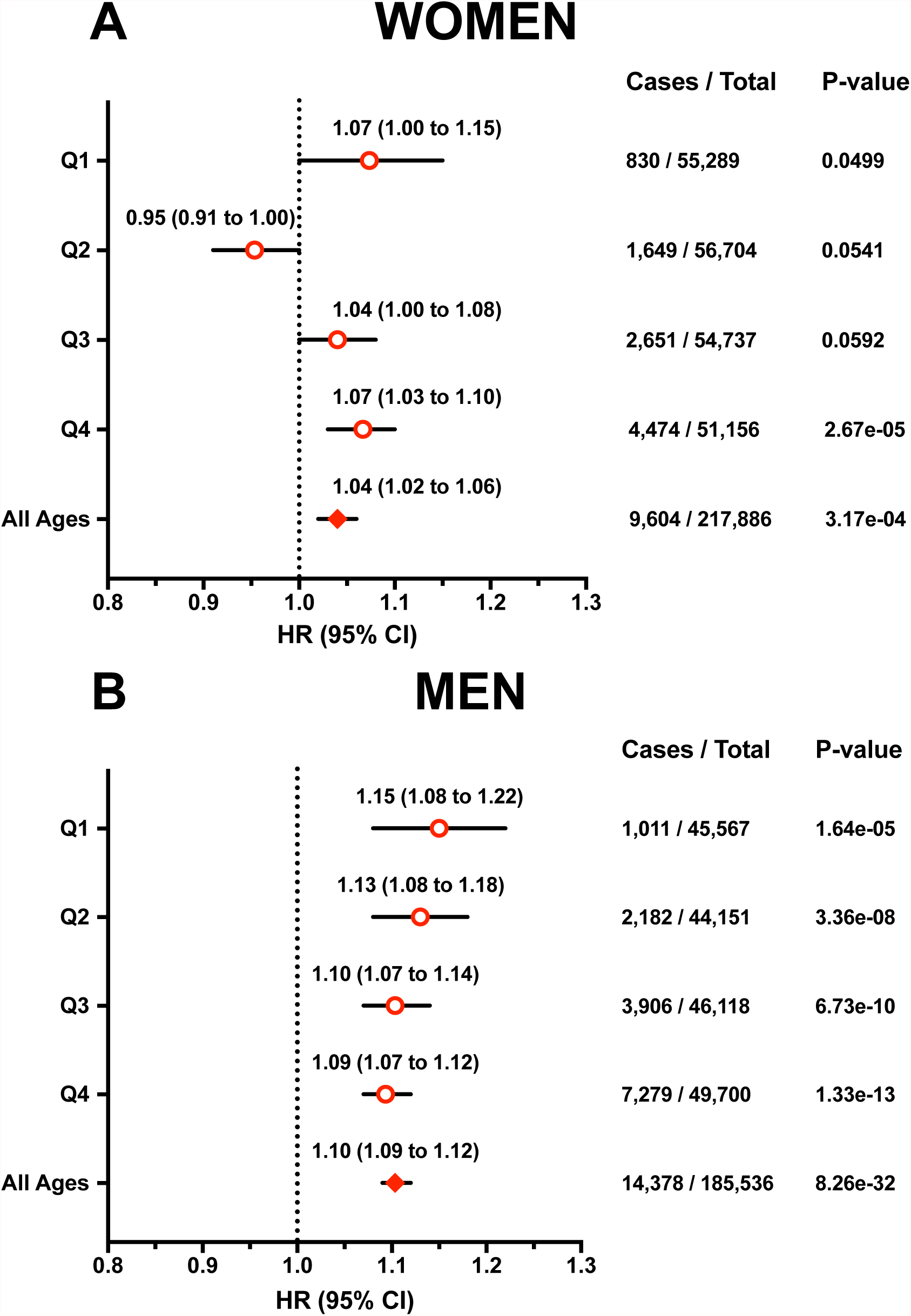
Association between PRS_CAD_ and all-cause mortality. Cox regression results were obtained from models adjusted for age and the first ten genetic principal components in women (A) and in men (B), by quartiles of age (Q1 to Q4) and for the whole group (all ages). Data are presented as estimated hazard ratios (HR) for mortality per SD increase of PRS_CAD_, with their 95% confidence interval (95% CI).

As observed for incident MI, being in the highest percentiles of PRS_CAD_ was associated with a significantly higher risk of mortality (Supplemental Figure 9 and Supplemental Table 9). In all individuals at the 90^th^ percentile threshold, HR_MORTALITY_ reached 1.18 (95% CI [1.13 to 1.23], p = 6.86e-16), whereas in women and men, HR_MORTALITY_ were respectively 1.16 (95% CI [1.09 to 1.24], p = 3.76e-06) and 1.19 (95% CI [1.13 to 1.26], p = 2.05e-11). When age and sex stratified, the strongest associations were found in the first age quartile, with a corresponding HR_MORTALITY_ of 1.42 (95% CI [1.19 to 1.70], p = 1.35e-04) in men (Supplemental Figure 9 and Supplemental Table 9).

Kaplan-Meier curves showed modest but significant separation of survival curves among tertiles of PRS_CAD_ in women (Log-rank test, p = 0.02, Figure 7B), and a significant separation in survival curves was observed in men (Log-rank test, p < 0.0001, Figure 7C).

**Figure 7:**
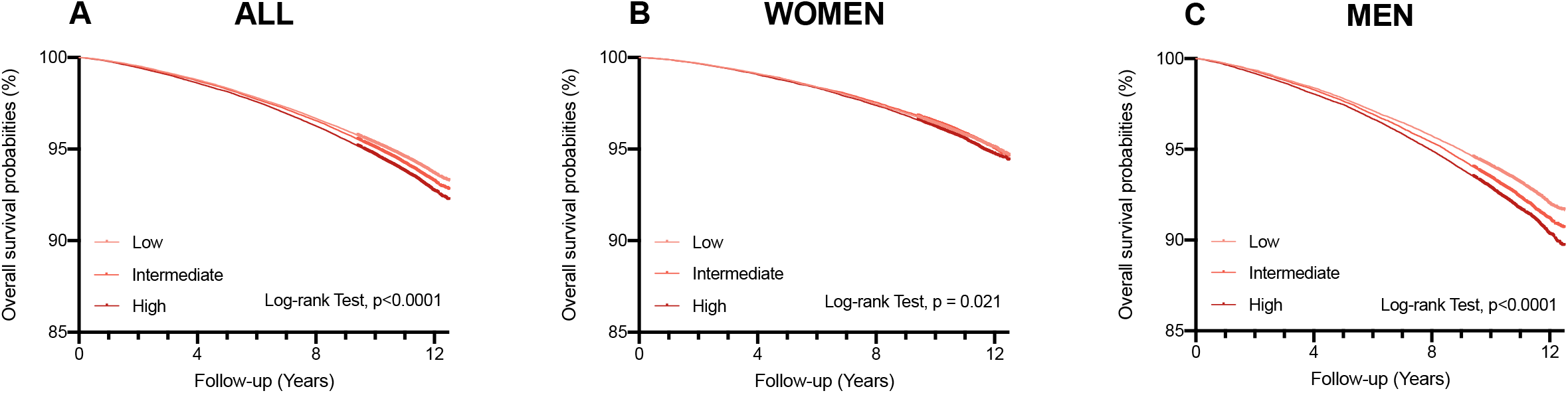
Survival probability according to genetic risk. Overall survival probability curves were obtained from Kaplan-Meier estimates over a median follow-up of 11.0 years, for tertiles of PRS_CAD_ (low, intermediate, high) in all individuals (A), and separately in women (B) and in men (C). Log-rank test, p-values <0.0001 for A and C, and 0.02 for B.

After 10 years of follow-up, cumulative mortality estimates ranged between 3.61% (low PRS) and 3.78% (high PRS) in women, and between 5.92% (low PRS) and 7.11% (high PRS) in men. Individuals above the 90^th^ percentile threshold presented overall 10-year cumulative mortality of 4.11% and 7.41%, respectively for women and men (Supplemental Figure 10).

## DISCUSSION

We derived a polygenic risk score for CAD using a recognized Bayesian approach and evaluated its predictive accuracy for incident myocardial infarction and all-cause mortality, in various age and sex groups using recent data from UK Biobank.

Our study supports 4 major conclusions.

First, the association between PRS_CAD_ and incident MI varies according to age and sex. Younger men (aged between 40 and 51 years) showed significantly higher HR_MI_ compared to the other groups tested. Recently, sex-stratified Cox regressions using different PRS_CAD_ in UKB revealed higher HRs for incident CAD in men compared to women^25,26^, although the analysis was not performed in all age groups. Another study in UKB with a mean follow-up period of 6.2 years showed that men with a high PRS_CAD_ are at a significantly higher risk^17^. The better performance observed in men could be due to the overrepresentation of men as cases in CAD GWAS. The use of sex-specific association data could potentially improve the predictive accuracy of PRS, notably for women.

Our results suggest a more important genetic component in early-onset events, as the prediction accuracy of MI incidence was the highest in the youngest quartiles of age in both women and men. This last observation is in line with a previous study showing that a genetic risk score of 182 variants was strongly associated with early-onset CAD (before 40 years old)^15^. A recent study in the FinnGen cohort reported that using the top decile of a PRS_CAD_ including >6 million SNPs could identify 12.6% of coronary heart disease cases missed by clinical risk scores in individuals under 55 years old, compared to only 2.5% of cases in older individuals^22^. We also report a significant inverse correlation between PRS_CAD_ and age at first incident MI in men, supporting the better performance to predict early-onset events.

Second, PRS_CAD_ was significantly associated with all-cause mortality in both sexes, with a stronger association in men. This is consistent with a recent study that showed an association between a PRS_CAD_ (207 SNPs) and all-cause mortality in men only in UKB^23^. We also observed a small but significant difference in PRS_CAD_ between age groups, with older individuals having on average a lower PRS. This finding could support a potential impact of PRS_CAD_ on longevity.

Third, PRS_CAD_ brings complementary support in the identification of patients at high risk of MI, independently of traditional risk factors and clinical scores. We observed significant continuous NRI when adding the PRS to models including only clinical risk factors in all age and sex groups, with a stronger improvement in younger men, corroborating recent findings^26^. Our results are also in line with observations from a recent analysis in the Malmö Diet and Cancer Study, in which PCE-derived risk was categorized and compared with a PRS_CAD_ including >6 million SNPs. The study showed a categorized NRI of 0.165 for CAD incidence over a 10-year period^24^. Our results contrast with the conclusions of two recent studies^18,19^. Mosley et al^18^ showed no significant improvement in reclassification with models including both PCE and a PRS_CAD_ of >6 million SNPs (versus PCE alone) in two cohorts (ARIC and MESA), even if the PRS was associated with the incidence of coronary heart disease. The discrepancy with our results could be in part due to the older population in these cohorts, which respectively included individuals with a mean age at baseline of 62.9 and 61.8 years (compared to 56.9 years in this study). The study by Elliott et al^19^ in UKB, with a median follow-up of 8 years, showed a modest but significant categorized-NRI for incident CAD (risk threshold of 7.5%) when adding a PRS_CAD_ of 1 million SNPs (derived using the lassosum method). Notably, the PRS had a higher predictive accuracy in participants <55 years, although reclassification analyses were not performed in specific age groups. In our study, discriminative capacity and NRI were significantly higher in the younger group of men, supporting a potential utility to predict early-onset events. In fact, a PRS that can be measured from birth could improve the predictive performance of clinical scores which rely heavily on age and have lower predictive accuracy in young individuals.

Fourth, we propose the use of a threshold to identify patients at high genetic risk of MI. We demonstrate that hazard ratios for MI for individuals in the top percentiles of the PRS_CAD_ increase gradually. Women and men above the 90^th^ percentile of PRS_CAD_ have a more than two-fold increase in risk for incident MI and a 15 to 20% increase in risk of all-cause mortality. We therefore suggest the 90^th^ percentile of the distribution as a threshold to identify patients at high risk, in line with other studies^20,22^. A similar threshold was previously used to identify patients with a high polygenic risk who benefit more from lipid-lowering treatments^12,14,20,21^. Notably, a recent study showed that individuals with a PRS_CAD_ including >6 millions of SNPs above the 90^th^ percentile had a higher benefit from alirocumab, a PCSK9 inhibitor, with significant reduction of absolute and relative risk. The data from the latter study was obtained from ODYSSEY OUTCOME, a clinical trial including patients with a previous diagnosis of CAD^20^. Similar results were observed in statin trials for primary prevention^12,14^. All in all, the 90^th^ percentile of PRS_CAD_ could be used as a threshold to identify high-risk individuals for selection in clinical trials and the use of more aggressive preventative measures.

Our study has limitations. First, our analyses were restricted to events occurring after the age of 40, the minimum age at recruitment in UKB. In addition, individuals with a very high genetic risk could be underrepresented in the older groups since they are more likely to have early-onset events. Second, this study is limited to individuals of European ancestry. Results need to be validated in other ethnic groups. Third, the number of events was lower for women than men, which may have decreased the power to detect significant associations. Fourth, the PCE is validated for ASCVD and not specifically to predict MI. MI was selected as the main outcome for incident analyses because of its severity and more robust phenotype.

In conclusion, we show that a polygenic risk score for CAD is independently associated with incident MI and all-cause mortality, and has a better predictive accuracy in men between 40 and 51 years old. The use of a PRS threshold to identify individuals at high risk of an early-onset MI should be considered.

## Data Availability

Data that support our findings are available through appropriate application to UK Biobank (UKB).Data access permission for this study was granted under UKB application 25205. Computational codes are available, without compromising sensitive individual-level data, from the corresponding author upon reasonable request.

## ACKNOWLEDGEMENTS

We would like to thank all study participants and scientists involved in the UK Biobank study.

## REFERENCES

1. GBD 2015 Mortality and Causes of Death Collaborators. Global, regional, and national life expectancy, all-cause mortality, and cause-specific mortality for 249 causes of death, 1980-2015: a systematic analysis for the Global Burden of Disease Study 2015. Lancet (London, England) 2016;388:1459–1544. doi:10.1016/S0140-6736(16)31012-1.

2. Khera A v., Emdin CA, Drake I, Natarajan P, Bick AG, Cook NR, et al. Genetic risk, adherence to a healthy lifestyle, and coronary disease. New England Journal of Medicine 2016;375:2349–2358. doi:10.1056/NEJMoa1605086.

3. Libby P, Buring JE, Badimon L, Hansson GK, Deanfield J, Bittencourt MS, et al. Atherosclerosis. Nature Reviews Disease Primers 2019;5:56. doi:10.1038/s41572-019-0106-z.

4. Michos ED, McEvoy JW, Blumenthal RS. Lipid Management for the Prevention of Atherosclerotic Cardiovascular Disease. New England Journal of Medicine 2019;381:1557–1567. doi:10.1056/nejmra1806939.

5. Goff DC, Lloyd-Jones DM, Bennett G, Coady S, D’Agostino RB, Gibbons R, et al. 2013 ACC/AHA Guideline on the Assessment of Cardiovascular Risk. Journal of the American College of Cardiology 2014;63:2935–2959. doi:10.1016/j.jacc.2013.11.005.

6. Lloyd-Jones DM, Braun LT, Ndumele CE, Smith SC, Sperling LS, Virani SS, et al. Use of Risk Assessment Tools to Guide Decision-Making in the Primary Prevention of Atherosclerotic Cardiovascular Disease: A Special Report from the American Heart Association and American College of Cardiology. Circulation 2019;139:E1162–E1177. doi:10.1161/CIR.0000000000000638.

7. Nikpay M, Goel A, Won HH, Hall LM, Willenborg C, Kanoni S, et al. A comprehensive 1000 Genomes-based genome-wide association meta-analysis of coronary artery disease. Nature Genetics 2015;47:1121–1130. doi:10.1038/ng.3396.

8. Nelson CP, Goel A, Butterworth AS, Kanoni S, Webb TR, Marouli E, et al. Association analyses based on false discovery rate implicate new loci for coronary artery disease. Nature Genetics 2017;49:1385–1391. doi:10.1038/ng.3913.

9. Vilhjálmsson BJ, Yang J, Finucane HK, Gusev A, Lindström S, Ripke S, et al. Modeling Linkage Disequilibrium Increases Accuracy of Polygenic Risk Scores. The American Journal of Human Genetics 2015;97:576–592. doi:10.1016/j.ajhg.2015.09.001.

10. Ripatti S, Tikkanen E, Orho-Melander M, Havulinna AS, Silander K, Sharma A, et al. A multilocus genetic risk score for coronary heart disease: Case-control and prospective cohort analyses. The Lancet 2010;376:1393–1400. doi:10.1016/S0140-6736(10)61267-6.

11. Ding K, Bailey KR, Kullo IJ. Genotype-informed estimation of risk of coronary heart disease based on genome-wide association data linked to the electronic medical record. BMC Cardiovascular Disorders 2011;11. doi:10.1186/1471-2261-11-66.

12. Mega JL, Stitziel NO, Smith JG, Chasman DI, Caulfield MJ, Devlin JJ, et al. Genetic risk, coronary heart disease events, and the clinical benefit of statin therapy: An analysis of primary and secondary prevention trials. The Lancet 2015;385:2264–2271. doi:10.1016/S0140-6736(14)61730-X.

13. Tada H, Melander O, Louie JZ, Catanese JJ, Rowland CM, Devlin JJ, et al. Risk prediction by genetic risk scores for coronary heart disease is independent of self-reported family history. European Heart Journal 2016;37:561–567. doi:10.1093/eurheartj/ehv462.

14. Natarajan P, Young R, Stitziel NO, Padmanabhan S, Baber U, Mehran R, et al. Polygenic risk score identifies subgroup with higher burden of atherosclerosis and greater relative benefit from statin therapy in the primary prevention setting. Circulation 2017;135:2091–2101. doi:10.1161/CIRCULATIONAHA.116.024436.

15. Thériault S, Lali R, Chong M, Velianou JL, Natarajan MK, Paré G. Polygenic Contribution in Individuals With Early-Onset Coronary Artery Disease. Circulation Genomic and Precision Medicine 2018;11:e001849. doi:10.1161/CIRCGEN.117.001849.

16. Khera A v, Chaffin M, Aragam KG, Haas ME, Roselli C, Choi SH, et al. Genome-wide polygenic scores for common diseases identify individuals with risk equivalent to monogenic mutations 2018;50.

17. Inouye M, Abraham G, Nelson CP, Wood AM, Sweeting MJ, Dudbridge F, et al. Genomic Risk Prediction of Coronary Artery Disease in 480,000 Adults. Journal of the American College of Cardiology 2018;72:1883–1893. doi:10.1016/j.jacc.2018.07.079.

18. Mosley JD, Gupta DK, Tan J, Yao J, Wells QS, Shaffer CM, et al. Predictive Accuracy of a Polygenic Risk Score Compared With a Clinical Risk Score for Incident Coronary Heart Disease. JAMA 2020;323:627–635. doi:10.1001/jama.2019.21782.

19. Elliott J, Bodinier B, Bond TA, Chadeau-Hyam M, Evangelou E, Moons KGM, et al. Predictive Accuracy of a Polygenic Risk Score-Enhanced Prediction Model vs a Clinical Risk Score for Coronary Artery Disease. JAMA 2020;323:636–645. doi:10.1001/jama.2019.22241.

20. Damask A, Steg PG, Schwartz GG, Szarek M, Hagström E, Badimon L, et al. Patients with High Genome-Wide Polygenic Risk Scores for Coronary Artery Disease May Receive Greater Clinical Benefit from Alirocumab Treatment in the ODYSSEY OUTCOMES Trial. Circulation 2020:624–636. doi:10.1161/CIRCULATIONAHA.119.044434.

21. Marston NA, Kamanu FK, Nordio F, Gurmu Y, Roselli C, Sever PS, et al. Predicting Benefit from Evolocumab Therapy in Patients with Atherosclerotic Disease Using a Genetic Risk Score. Circulation 2020:616–623. doi:10.1161/CIRCULATIONAHA.119.043805.

22. Mars N, Koskela JT, Ripatti P, Kiiskinen TTJ, Havulinna AS, Lindbohm J v., et al. Polygenic and clinical risk scores and their impact on age at onset and prediction of cardiometabolic diseases and common cancers. Nature Medicine 2020;26:549–557. doi:10.1038/s41591-020-0800-0.

23. Meisner A, Kundu P, Zhang YD, Lan L v, Kim S, Ghandwani D, et al. Combined Utility of 25 Disease and Risk Factor Polygenic Risk Scores for Stratifying Risk of All-Cause Mortality. American Journal of Human Genetics 2020;107:418–431. doi:10.1016/j.ajhg.2020.07.002.

24. Hindy G, Aragam KG, Ng K, Chaffin M, Lotta LA, Baras A, et al. Genome-wide polygenic score, clinical risk factors, and long-term trajectories of coronary artery disease. Arteriosclerosis, Thrombosis, and Vascular Biology 2020:2738–2746. doi:10.1161/ATVBAHA.120.314856.

25. Huang Y, Hui Q, Gwinn M, Hu Y, Quyyumi AA, Vaccarino V, et al. Sexual Differences in Genetic Predisposition of Coronary Artery Disease. Circulation Genomic and Precision Medicine 2021;14:e003147. doi:10.1161/CIRCGEN.120.003147.

26. Riveros-Mckay F, Weale ME, Moore R, Selzam S, Krapohl E, Sivley RM, et al. An Integrated Polygenic Tool Substantially Enhances Coronary Artery Disease Prediction. Circulation: Genomic and Precision Medicine 2021. doi:10.1161/circgen.120.003304.

27. Bycroft C, Freeman C, Petkova D, Band G, Elliott LT, Sharp K, et al. The UK Biobank resource with deep phenotyping and genomic data. Nature 2018;562:203–209. doi:10.1038/s41586-018-0579-z.

28. Auton A, Abecasis GR, Altshuler DM, Durbin RM, Bentley DR, Chakravarti A, et al. A global reference for human genetic variation. Nature 2015;526:68–74. doi:10.1038/nature15393.

29. Pencina MJ, D’Agostino RB, Steyerberg EW. Extensions of net reclassification improvement calculations to measure usefulness of new biomarkers. Statistics in Medicine 2011;30:11–21. doi:10.1002/sim.4085.

30. Kerr KF, Wang Z, Janes H, McClelland RL, Psaty BM, Pepe MS. Net Reclassification Indices for Evaluating Risk Prediction Instruments: A critical review. Epidemiology 2014;25:114–121. doi:10.1097/EDE.0000000000000018.

