## Supplemental Tables for "A Polygenic Risk Score for Coronary Artery Disease Improves the Prediction of Early-Onset Myocardial Infarction and Mortality in Men"

**Supplemental Table 1: Characteristics of the cohort.** ApoB indicates Apolipoprotein B; ASCVD, Atherosclerotic Cardiovascular Disease; BMI, Body Mass Index; HDL-C, High Density Lipoprotein Cholesterol; HES, Hospital Episode Statistics; IQR, Interquartile Range; LDL-C, Low Density Lipoprotein Cholesterol; Lp(a), Lipoprotein (a); MI, Myocardial Infarction; PCE, Pooled Cohort Equation; SD, Standard Deviation; TC, Total Cholesterol

| Categories | Sub-categories | Whole Cohort | Training Subset | Validation Subset |
|---|---|---|---|---|
| n (%) | All | 408,422 | 5,000 | 403,422 |
| | Women | 220,802 (54.06) | 2,916 (58.32) | 217,886 (54.01) |
| | Men | 187,620 (45.94) | 2,084 (41.68) | 185,536 (45.99) |
| Age at baseline, y (± SD) | All | 56.91 (8.00) | 55.97 (7.95) | 56.93 (8.00) |
| | Women | 56.72 (7.92) | 55.95 (7.90) | 56.73 (7.92) |
| | Men | 57.15 (8.09) | 55.99 (8.03) | 57.16 (8.09) |
| BMI, kg/m2 (± SD) | | 27.42 (4.75) | 26.97 (4.52) | 27.42 (4.76) |
| Blood Pressure, mmHg (± SD) | Diastolic | 82.27 (10.11) | 82.12 (10.09) | 82.27 (10.11) |
| | Systolic | 138.29 (18.64) | 137.22 (18.66 | 138.30 (18.64) |
| Blood lipids, mmol/L (± SD) | LDL-C | 3.57 (0.87) | 3.72 (0.81) | 3.57 (0.87) |
| | HLD-C | 1.45 (0.38) | 1.49 (0.38) | 1.45 (0.38) |
| | TC | 5.71 (1.14) | 5.92 (1.06) | 5.71 (1.15) |
| | APO-B | 1.03 (0.24) | 1.07 (0.23) | 1.07 (0.24) |
| nmol/L (± SD) | Lp(a) | 44.09 (49.45) | 44.63 (49.97) | 44.09 (49.45) |
| Hypertension, n (%) | | 91,201 (22.33) | 659 (13.18) | 90,542 (22.44) |
| Diabetes, n (%) | | 20,620 (5.05) | 98 (1.96) | 20,522 (5.09) |
| Current Smoker at baseline, n (%) | | 41,253 (10.10) | 490 (9.80) | 40,763 (10.10) |
| 10-year risk of first ASCVD event (PCE) | n (%) | 360,407 (88.24) | 4,720 (94.40) | 355,687 (88.17) |
| | Risk % (± SD) | 8.38 (7.66) | 7.36 (7.15) | 8.39 (7.66) |
| HES CAD Outcome, n (%) | Prevalent (all cases) | 32,694 (8.00) | 219 (4.38) | 32,475 (8.05) |
| HES MI Outcome, n (%) | Prevalent (all cases) | 14,924 (3.65) | 97 (1.94) | 14,827 (3.67) |
| HES MI-Revascularisation Outcome, n (%) | Prevalent (all cases) | 20,769 (5.08) | 131 (2.62) | 20,638 (5.12) |
| Mortality, n (%) Before March 2020 | All-cause | 24,229 (5.93) | 247 (4.94) | 23,982 (5.94) |
| Follow-up, y (± IQR) | | 11.04 (1.40) | 11.10 (1.35) | 11.04 (1.40) |

**Supplemental Table 2: Association between $PRS_{CAD}$ and prevalent CAD in the training dataset.** Non-adjusted and adjusted models are used in logistic regression with prevalent cardiovascular outcomes. Data are presented as logistic-regression odd ratios (OR) per SD increase of $PRS_{CAD}$ with their 95% confidence intervals (95% CI), logistic regression P-value significance and corresponding area under the receiver operating characteristic curve (AUC) values. MI, Myocardial Infarction; MI-REVASC: MI or Revascularisation Procedure; $PRS_{CAD}$, Polygenic Risk Score for Coronary Artery Disease; PCE, Pooled Cohort Equation; PCs, First Ten Genetic Principal Components; CI: Confidence Interval

| LDPred Rho Threshold | Covariables | CAD (n = 219) | | | MI (n = 97) | | | MI-REVASC. (n = 131) | | |
|---|---|---|---|---|---|---|---|---|---|---|
| | | OR (95% CI) | P-value | AUC | OR (95% CI) | P-value | AUC | OR (95% CI) | P-value | AUC |
| 0.1 | $PRS_{CAD}$ | 1.36 (1.19 to 1.56) | 9.15E-06 | 0.588 | 1.27 (1.03 to 1.55) | 2.21E-02 | 0.570 | 1.45 (1.21 to 1.72) | 3.59E-05 | 0.603 |
| | $PRS_{CAD}$ + PCs | 1.36 (1.19 to 1.56) | 1.12E-05 | 0.599 | 1.27 (1.03 to 1.55) | 2.26E-02 | 0.603 | 1.45 (1.21 to 1.73) | 3.81E-05 | 0.615 |
| | $PRS_{CAD}$ + AGE | 1.38 (1.20 to 1.58) | 6.90E-06 | 0.691 | 1.27 (1.04 to 1.56) | 2.06E-02 | 0.663 | 1.46 (1.22 to 1.74) | 2.91E-05 | 0.691 |
| | $PRS_{CAD}$ + AGE + SEX + PCs | 1.38 (1.20 to 1.59) | 4.47E-06 | 0.737 | 1.28 (1.04 to 1.57) | 1.71E-02 | 0.736 | 1.47 (1.23 to 1.75) | 2.16E-05 | 0.754 |
| 0.03 | $PRS_{CAD}$ | 1.42 (1.24 to 1.63) | 5.72E-07 | 0.599 | 1.36 (1.11 to 1.67) | 2.57E-03 | 0.591 | 1.53 (1.28 to 1.82) | 2.06E-06 | 0.619 |
| | $PRS_{CAD}$ + PCs | 1.42 (1.23 to 1.63) | 6.81E-07 | 0.609 | 1.36 (1.11 to 1.67) | 2.73E-03 | 0.620 | 1.53 (1.28 to 1.83) | 2.21E-06 | 0.631 |
| | $PRS_{CAD}$ + AGE | 1.43 (1.25 to 1.65) | 3.78E-07 | 0.695 | 1.37 (1.12 to 1.68) | 2.37E-03 | 0.671 | 1.54 (1.29 to 1.84) | 1.58E-06 | 0.698 |
| | $PRS_{CAD}$ + AGE + SEX + PCs | 1.44 (1.26 to 1.66) | 2.14E-07 | 0.740 | 1.38 (1.13 to 1.69) | 1.86E-03 | 0.741 | 1.55 (1.30 to 1.85) | 1.06E-06 | 0.760 |
| 0.01 | $PRS_{CAD}$ | 1.44 (1.26 to 1.65) | 1.78E-07 | 0.600 | 1.43 (1.17 to 1.76) | 5.10E-04 | 0.597 | 1.57 (1.32 to 1.87) | 5.26E-07 | 0.621 |
| | $PRS_{CAD}$ + PCs | 1.44 (1.26 to 1.66) | 1.94E-07 | 0.611 | 1.43 (1.17 to 1.75) | 5.76E-04 | 0.627 | 1.57 (1.32 to 1.88) | 5.68E-07 | 0.634 |
| | $PRS_{CAD}$ + AGE | 1.46 (1.27 to 1.68) | 7.73E-08 | 0.694 | 1.44 (1.18 to 1.77) | 4.34E-04 | 0.672 | 1.59 (1.33 to 1.90) | 3.14E-07 | 0.698 |
| | $PRS_{CAD}$ + AGE + SEX + PCs | 1.48 (1.29 to 1.70) | 3.45E-08 | 0.742 | 1.46 (1.19 to 1.78) | 3.10E-04 | 0.744 | 1.61 (1.35 to 1.92) | 1.75E-07 | 0.762 |
| 0.003 | $PRS_{CAD}$ | 1.43 (1.25 to 1.64) | 3.14E-07 | 0.594 | 1.45 (1.18 to 1.77) | 3.69E-04 | 0.586 | 1.54 (1.29 to 1.84) | 1.50E-06 | 0.606 |
| | $PRS_{CAD}$ + PCs | 1.44 (1.25 to 1.65) | 3.15E-07 | 0.609 | 1.44 (1.18 to 1.77) | 4.37E-04 | 0.621 | 1.54 (1.29 to 1.84) | 1.60E-06 | 0.623 |
| | $PRS_{CAD}$ + AGE | 1.46 (1.27 to 1.68) | 9.70E-08 | 0.692 | 1.46 (1.19 to 1.79) | 2.89E-04 | 0.669 | 1.57 (1.31 to 1.87) | 7.36E-07 | 0.692 |
| | $PRS_{CAD}$ + AGE + SEX + PCs | 1.48 (1.29 to 1.71) | 3.67E-08 | 0.739 | 1.47 (1.20 to 1.81) | 1.99E-04 | 0.744 | 1.59 (1.33 to 1.90) | 3.65E-07 | 0.759 |
| 0.001 | $PRS_{CAD}$ | 1.37 (1.19 to 1.58) | 7.59E-06 | 0.583 | 1.39 (1.13 to 1.71) | 1.67E-03 | 0.576 | 1.43 (1.20 to 1.71) | 8.20E-05 | 0.588 |
| | $PRS_{CAD}$ + PCs | 1.38 (1.20 to 1.58) | 7.54E-06 | 0.601 | 1.38 (1.13 to 1.70) | 1.98E-03 | 0.613 | 1.43 (1.20 to 1.71) | 8.69E-05 | 0.608 |
| | $PRS_{CAD}$ + AGE | 1.40 (1.22 to 1.61) | 2.51E-06 | 0.688 | 1.40 (1.14 to 1.72) | 1.31E-03 | 0.665 | 1.45 (1.21 to 1.73) | 4.28E-05 | 0.684 |
| | $PRS_{CAD}$ + AGE + SEX + PCs | 1.42 (1.23 to 1.63) | 1.13E-06 | 0.734 | 1.41 (1.15 to 1.73) | 1.01E-03 | 0.740 | 1.47 (1.23 to 1.75) | 2.48E-05 | 0.751 |
| 0.0003 | $PRS_{CAD}$ | 1.12 (0.98 to 1.29) | 8.83E-02 | 0.533 | 0.97 (0.79 to 1.19) | 7.60E-01 | 0.513 | 1.05 (0.89 to 1.25) | 5.61E-01 | 0.510 |
| | $PRS_{CAD}$ + PCs | 1.13 (0.99 to 1.30) | 7.58E-02 | 0.564 | 0.98 (0.80 to 1.20) | 8.42E-01 | 0.573 | 1.06 (0.89 to 1.26) | 5.07E-01 | 0.548 |
| | $PRS_{CAD}$ + AGE | 1.13 (0.99 to 1.30) | 7.18E-02 | 0.674 | 0.98 (0.81 to 1.20) | 8.82E-01 | 0.651 | 1.07 (0.90 to 1.27) | 4.76E-01 | 0.664 |
| | $PRS_{CAD}$ + AGE + SEX + PCs | 1.13 (0.98 to 1.29) | 8.50E-02 | 0.725 | 0.98 (0.80 to 1.20) | 8.11E-01 | 0.727 | 1.06 (0.89 to 1.26) | 5.29E-01 | 0.735 |

**Supplemental Table 3: Association between $PRS_{CAD}$ and prevalent cardiovascular outcomes in mutlivariable models.** Non-adjusted and adjusted models are used in logistic regression with CAD, MI and MI-REVASC. outcomes. Data are presented as logistic-regression odd ratios (OR) per SD increase of $PRS_{CAD}$ and their 95% confidence intervals (95% CI), Nagelkerke's pseudo-$R^2$ corresponding area under the receiver operating characteristic curve (AUC) values.

BMI, Body Mass Index; HDL-C, High Density Lipoprotein Cholesterol; LDL-C, Low Density Lipoprotein Cholesterol; Lp(a), Lipoprotein (a); MI, Myocardial Infarction; MI-REVASC: MI or Revascularisation Procedure; PCE, Pooled Cohort Equation; PCs, First Ten Genetic Principal Components; $PRS_{CAD}$, Polygenic Risk Score for Coronary Artery Disease; TC, Total Cholesterol

| Outcome | Logisitic Regression Models | Event (n) | Control (n) | Total (n) | OR (95% CI) | Pseudo-$R^2$ | AUC |
|---|---|---|---|---|---|---|---|
| CAD | Model 1: AGE + SEX | 32,475 | 370,947 | 403,422 | NA | 0.127 | 0.742 |
| | Model 2: $PRS_{CAD}$ + PCs | 32,475 | 370,947 | 403,422 | 1.50 (1.49 to 1.52) | 0.0280 | 0.613 |
| | Model 3: $PRS_{CAD}$ + AGE + SEX + PCs | 32,475 | 370,947 | 403,422 | 1.56 (1.54 to 1.58) | 0.158 | 0.766 |
| | Model 4: PCE | 27,568 | 328,119 | 355,687 | NA | 0.107 | 0.749 |
| | Model 5: $PRS_{CAD}$ + PCE + PCs | 27,568 | 328,119 | 355,687 | 1.51 (1.49 to 1.53) | 0.132 | 0.758 |
| | Model 6: $PRS_{CAD}$ + AGE + SEX + PCs + LDL + HDL + TC + BMI + SBP + DIABETES + SMOKING | 27,516 | 327,628 | 355,144 | 1.55 (1.53 to 1.57) | 0.205 | 0.797 |
| MI | Model 1: AGE + SEX | 14,827 | 388,595 | 403,422 | NA | 0.0990 | 0.744 |
| | Model 2: $PRS_{CAD}$ + PCs | 14,827 | 388,595 | 403,422 | 1.58 (1.56 to 1.61) | 0.0280 | 0.629 |
| | Model 3: $PRS_{CAD}$ + AGE + SEX + PCs | 14,827 | 388,595 | 403,422 | 1.63 (1.60 to 1.65) | 0.129 | 0.772 |
| | Model 4: PCE | 12,517 | 343,170 | 355,687 | NA | 0.0800 | 0.748 |
| | Model 5: $PRS_{CAD}$ + PCE + PCs | 12,517 | 343,170 | 355,687 | 1.58 (1.55 to 1.61) | 0.105 | 0.761 |
| | Model 6: $PRS_{CAD}$ + AGE + SEX + PCs + LDL + HDL + TC + BMI + SBP + DIABETES + SMOKING | 12,495 | 342,649 | 355,144 | 1.60 (1.57 to 1.63) | 0.172 | 0.807 |
| MI-REVASC. | Model 1: AGE + SEX | 20,638 | 382,784 | 403,422 | NA | 0.123 | 0.756 |
| | Model 2: $PRS_{CAD}$ + PCs | 20,638 | 382,784 | 403,422 | 1.66 (1.63 to 1.68) | 0.0370 | 0.640 |
| | Model 3: $PRS_{CAD}$ + AGE + SEX + PCs | 20,638 | 382,784 | 403,422 | 1.73 (1.70 to 1.75) | 0.162 | 0.789 |
| | Model 4: PCE | 17,402 | 338,285 | 355,687 | NA | 0.0960 | 0.758 |
| | Model 5: $PRS_{CAD}$ + PCE + PCs | 17,402 | 338,285 | 355,687 | 1.67 (1.65 to 1.70) | 0.130 | 0.774 |
| | Model 6: $PRS_{CAD}$ + AGE + SEX + PCs + LDL + HDL + TC + BMI + SBP + DIABETES + SMOKING | 17,373 | 337,771 | 355,144 | 1.71 (1.68 to 1.74) | 0.205 | 0.817 |

**Supplemental Table 4: Association between $PRS_{CAD}$ and MI incidence in adjusted Cox regression models.** Adjusted Cox regression results from $PRS_{CAD}$ models including age and the first ten genetic principal components (PCs) as covariates were obtained in women and men, by quartiles of age (Q1 to Q4) and for the whole group (all age). Data are presented as estimated hazard ratios (HR) for incident MI outcome per SD increase of $PRS_{CAD}$, with their 95% confidence intervals (95% CI), and Cox regression P-value significance.

| Outcome | Sex Group | Age Group | Age Window | Max-Min Difference in Age Group | N Event | N Total | Ratio N Event / 100% of Event | HR (95% CI) | P-value |
|---|---|---|---|---|---|---|---|---|---|
| MI Incident | All | All Ages | from 40 to 73 | 33 | 7,746 | 393,725 | 100.00% | 1.53 (1.49 to 1.56) | 2.69E-296 |
| | | Quartile 1 | from 40 to 51 | 11 | 889 | 100,284 | 11.44% | 1.89 (1.77 to 2.02) | 1.15E-79 |
| | | Quartile 2 | from 51 to 58 | 7 | 1,602 | 99,321 | 20.68% | 1.59 (1.51 to 1.67) | 2.43E-74 |
| | | Quartile 3 | from 58 to 63 | 5 | 2,224 | 98,008 | 28.71% | 1.42 (1.36 to 1.48) | 4.22E-61 |
| | | Quartile 4 | from 63 to 73 | 10 | 3,031 | 96,112 | 39.13% | 1.48 (1.42 to 1.53) | 2.14E-98 |
| | Women | All Ages | from 40 to 71 | 31 | 2,295 | 216,020 | 100.00% | 1.45 (1.39 to 1.51) | 2.02E-70 |
| | | Quartile 1 | from 40 to 51 | 11 | 208 | 55,176 | 9.06% | 1.57 (1.37 to 1.79) | 9.04E-11 |
| | | Quartile 2 | from 51 to 58 | 7 | 427 | 56,422 | 18.61% | 1.43 (1.30 to 1.57) | 9.17E-14 |
| | | Quartile 3 | from 58 to 63 | 5 | 662 | 54,227 | 28.85% | 1.40 (1.30 to 1.51) | 7.90E-18 |
| | | Quartile 4 | from 63 to 71 | 8 | 998 | 50,195 | 43.49% | 1.47 (1.38 to 1.57) | 1.68E-33 |
| | Men | All Ages | from 40 to 73 | 33 | 5,451 | 177,705 | 100.00% | 1.56 (1.52 to 1.60) | 1.02E-228 |
| | | Quartile 1 | from 40 to 51 | 11 | 681 | 45,108 | 12.49% | 2.00 (1.86 to 2.16) | 1.93E-72 |
| | | Quartile 2 | from 51 to 58 | 7 | 1,175 | 42,899 | 21.56% | 1.65 (1.56 to 1.75) | 1.71E-63 |
| | | Quartile 3 | from 58 to 63 | 5 | 1,562 | 43,781 | 28.66% | 1.43 (1.36 to 1.50) | 7.19E-45 |
| | | Quartile 4 | from 63 to 73 | 10 | 2,033 | 45,917 | 37.30% | 1.48 (1.41 to 1.54) | 1.04E-66 |

**Supplemental Table 5: Comparison between adjusted Cox regression models for MI incidence including PCE and $PRS_{CAD}$.** Data are presented as estimated hazard ratios (HR) for incident MI outcome per SD increase of $PRS_{CAD}$, with their 95% confidence intervals (95% CI), and Cox regression p-value significance. *Sex adjustment was only performed in "all" group.

MI: Myocardial Infarction; $PRS_{CAD}$: Polygenic Risk Score for Coronary Artery Disease; PCE: Pooled Cohort Equation; PCs: First Ten Genetic Principal Components

| Outcome | Sex Group | Age Group | $PRS_{CAD}$ + AGE + SEX* + PCs | | | | $PRS_{CAD}$ + PCE + PCs | | | |
|---|---|---|---|---|---|---|---|---|---|---|
| | | | N Event | N Total | HR (95% CI) | P-value | N Event | N Total | HR (95% CI) | P-value |
| MI Incident | All | All Ages | 7,746 | 393,725 | 1.53 (1.49 to 1.56) | 2.69E-296 | 6,735 | 347,654 | 1.51 (1.48 to 1.55) | 5.81E-245 |
| | | Quartile 1 | 889 | 100,284 | 1.89 (1.77 to 2.02) | 1.15E-79 | 797 | 89,246 | 1.88 (1.75 to 2.01) | 1.61E-69 |
| | | Quartile 2 | 1,602 | 99,321 | 1.59 (1.51 to 1.67) | 2.43E-74 | 1,391 | 87,841 | 1.55 (1.47 to 1.64) | 8.64E-59 |
| | | Quartile 3 | 2,224 | 98,008 | 1.42 (1.36 to 1.48) | 4.22E-61 | 1,938 | 86,339 | 1.40 (1.34 to 1.46) | 8.17E-49 |
| | | Quartile 4 | 3,031 | 96,112 | 1.48 (1.42 to 1.53) | 2.14E-98 | 2,609 | 84,228 | 1.47 (1.41 to 1.53) | 7.01E-82 |
| | Women | All Ages | 2,295 | 216,020 | 1.45 (1.39 to 1.51) | 2.02E-70 | 1,976 | 191,065 | 1.43 (1.37 to 1.5) | 4.34E-57 |
| | | Quartile 1 | 208 | 55,176 | 1.57 (1.37 to 1.79) | 9.04E-11 | 190 | 49,124 | 1.55 (1.34 to 1.79) | 2.52E-09 |
| | | Quartile 2 | 427 | 56,422 | 1.43 (1.30 to 1.57) | 9.17E-14 | 373 | 49,965 | 1.36 (1.23 to 1.51) | 1.72E-09 |
| | | Quartile 3 | 662 | 54,227 | 1.40 (1.30 to 1.51) | 7.90E-18 | 572 | 47,885 | 1.37 (1.26 to 1.49) | 5.42E-14 |
| | | Quartile 4 | 998 | 50,195 | 1.47 (1.38 to 1.57) | 1.68E-33 | 841 | 44,091 | 1.46 (1.37 to 1.57) | 1.88E-27 |
| | Men | All Ages | 5,451 | 177,705 | 1.56 (1.52 to 1.60) | 1.02E-228 | 4,759 | 156,589 | 1.54 (1.50 to 1.59) | 4.50E-189 |
| | | Quartile 1 | 681 | 45,108 | 2.00 (1.86 to 2.16) | 1.93E-72 | 607 | 40,122 | 1.99 (1.84 to 2.16) | 1.72E-63 |
| | | Quartile 2 | 1,175 | 42,899 | 1.65 (1.56 to 1.75) | 1.71E-63 | 1,018 | 37,876 | 1.62 (1.52 to 1.73) | 8.73E-51 |
| | | Quartile 3 | 1,562 | 43,781 | 1.43 (1.36 to 1.50) | 7.19E-45 | 1,366 | 38,454 | 1.41 (1.34 to 1.49) | 1.71E-36 |
| | | Quartile 4 | 2,033 | 45,917 | 1.48 (1.41 to 1.55) | 1.04E-66 | 1,768 | 40,137 | 1.47 (1.40 to 1.54) | 1.74E-55 |

**Supplemental Table 6: MI incidence in individuals at high genetic risk.** Adjusted Cox regression results were obtained for individuals above percentile thresholds (ranging between the 80th and the 99th percentile). Hazard ratios (HR) for MI incidence and their 95% confidence intervals (95% CI) were compared in all individuals, and separately in women and in men, for all ages and for each quartile of age (Q1 to Q4).

| Outcome | Sex Group | Age Group | Age Distribution | Percentile Thres. | N event | N total | HR (95% CI) | P-value |
|---|---|---|---|---|---|---|---|---|
| MI Incident | All | All Ages | from 40 to 73 | 80% | 2,528 | 76,217 | 2.02 (1.93 to 2.12) | 2.03E-185 |
| | | | | 90% | 1,454 | 37,919 | 2.19 (2.07 to 2.32) | 1.80E-159 |
| | | | | 95% | 806 | 18,881 | 2.36 (2.19 to 2.54) | 2.39E-117 |
| | | | | 97.50% | 435 | 9,409 | 2.51 (2.28 to 2.76) | 1.95E-77 |
| | | | | 99% | 211 | 3,727 | 3.04 (2.65 to 3.49) | 4.52E-57 |
| | | Quartile 1 | from 40 to 51 | 80% | 374 | 19,683 | 2.92 (2.55 to 3.33) | 9.30E-56 |
| | | | | 90% | 236 | 9,793 | 3.29 (2.83 to 3.82) | 2.68E-55 |
| | | | | 95% | 141 | 4,874 | 3.63 (3.03 to 4.34) | 1.24E-44 |
| | | | | 97.50% | 71 | 2,437 | 3.44 (2.70 to 4.39) | 1.86E-23 |
| | | | | 99% | 39 | 964 | 4.48 (3.25 to 6.17) | 6.13E-20 |
| | | Quartile 2 | from 51 to 58 | 80% | 543 | 19,322 | 2.13 (1.92 to 2.36) | 2.26E-46 |
| | | | | 90% | 317 | 9,616 | 2.33 (2.06 to 2.63) | 2.52E-41 |
| | | | | 95% | 173 | 4,794 | 2.44 (2.08 to 2.86) | 1.99E-28 |
| | | | | 97.50% | 102 | 2,382 | 2.89 (2.36 to 3.53) | 4.20E-25 |
| | | | | 99% | 48 | 946 | 3.45 (2.59 to 4.60) | 3.28E-17 |
| | | Quartile 3 | from 58 to 63 | 80% | 669 | 18,933 | 1.77 (1.61 to 1.93) | 1.02E-34 |
| | | | | 90% | 389 | 9,412 | 1.96 (1.75 to 2.18) | 2.99E-33 |
| | | | | 95% | 206 | 4,695 | 2.01 (1.74 to 2.32) | 1.34E-21 |
| | | | | 97.50% | 110 | 2,341 | 2.09 (1.73 to 2.53) | 4.61E-14 |
| | | | | 99% | 54 | 927 | 2.63 (2.00 to 3.44) | 2.54E-12 |
| | | Quartile 4 | from 63 to 73 | 80% | 953 | 18,270 | 1.90 (1.76 to 2.05) | 2.20E-60 |
| | | | | 90% | 528 | 9,084 | 1.98 (1.80 to 2.18) | 3.29E-46 |
| | | | | 95% | 289 | 4,517 | 2.10 (1.86 to 2.37) | 3.57E-33 |
| | | | | 97.50% | 155 | 2,248 | 2.22 (1.89 to 2.61) | 3.73E-22 |
| | | | | 99% | 75 | 887 | 2.64 (2.10 to 3.33) | 9.71E-17 |

| Outcome | Sex Group | Age Group | Age Distribution | Percentile Thres. | N event | N total | HR (95% CI) | P-value |
|---|---|---|---|---|---|---|---|---|
| MI Incident | Women | All Ages | from 40 to 71 | 80% | 704 | 42,500 | 1.80 (1.65 to 1.97) | 2.25E-38 |
| | | | | 90% | 436 | 21,166 | 2.16 (1.95 to 2.40) | 2.21E-47 |
| | | | | 95% | 252 | 10,549 | 2.41 (2.11 to 2.75) | 1.58E-39 |
| | | | | 97.50% | 148 | 5,253 | 2.80 (2.37 to 3.30) | 1.18E-33 |
| | | | | 99% | 75 | 2,086 | 3.51 (2.79 to 4.42) | 1.24E-26 |
| | | Quartile 1 | from 40 to 51 | 80% | 73 | 10,963 | 2.18 (1.64 to 2.89) | 9.07E-08 |
| | | | | 90% | 49 | 5,469 | 2.78 (2.01 to 3.82) | 4.32E-10 |
| | | | | 95% | 36 | 2,723 | 3.98 (2.78 to 5.70) | 5.17E-14 |
| | | | | 97.50% | 21 | 1,359 | 4.38 (2.79 to 6.88) | 1.40E-10 |
| | | | | 99% | 14 | 538 | 7.20 (4.18 to 12.4) | 1.02E-12 |
| | | Quartile 2 | from 51 to 58 | 80% | 130 | 11,155 | 1.76 (1.44 to 2.17) | 6.84E-08 |
| | | | | 90% | 82 | 5,561 | 2.16 (1.70 to 2.75) | 4.03E-10 |
| | | | | 95% | 48 | 2,774 | 2.42 (1.79 to 3.27) | 8.15E-09 |
| | | | | 97.50% | 30 | 1,381 | 2.98 (2.05 to 4.32) | 8.74E-09 |
| | | | | 99% | 16 | 549 | 3.93 (2.38 to 6.47) | 8.25E-08 |
| | | Quartile 3 | from 58 to 63 | 80% | 191 | 10,655 | 1.63 (1.38 to 1.93) | 1.08E-08 |
| | | | | 90% | 128 | 5,295 | 2.18 (1.80 to 2.65) | 2.24E-15 |
| | | | | 95% | 66 | 2,646 | 2.14 (1.66 to 2.75) | 5.01E-09 |
| | | | | 97.50% | 36 | 1,320 | 2.29 (1.64 to 3.21) | 1.28E-06 |
| | | | | 99% | 18 | 525 | 2.84 (1.78 to 4.54) | 1.25E-05 |
| | | Quartile 4 | from 63 to 71 | 80% | 311 | 9,728 | 1.83 (1.60 to 2.10) | 8.73E-19 |
| | | | | 90% | 182 | 4,838 | 2.04 (1.74 to 2.40) | 3.15E-18 |
| | | | | 95% | 108 | 2,402 | 2.35 (1.92 to 2.87) | 5.99E-17 |
| | | | | 97.50% | 63 | 1,192 | 2.68 (2.08 to 3.46) | 3.65E-14 |
| | | | | 99% | 29 | 473 | 3.00 (2.07 to 4.35) | 5.60E-09 |

| Outcome | Sex Group | Age Group | Age Distribution | Percentile Thres. | N event | N total | HR (95% CI) | P-value |
|---|---|---|---|---|---|---|---|---|
| MI Incident | Men | All Ages | from 40 to 73 | 80% | 1,843 | 33,698 | 2.12 (2.00 to 2.24) | 1.83E-150 |
| | | | | 90% | 1,041 | 16,730 | 2.21 (2.07 to 2.37) | 4.09E-117 |
| | | | | 95% | 570 | 8,316 | 2.34 (2.14 to 2.55) | 1.16E-81 |
| | | | | 97.50% | 296 | 4,147 | 2.37 (2.11 to 2.66) | 3.67E-47 |
| | | | | 99% | 139 | 1,639 | 2.77 (2.34 to 3.27) | 2.65E-32 |
| | | Quartile 1 | from 40 to 51 | 80% | 300 | 8,722 | 3.17 (2.73 to 3.69) | 1.80E-50 |
| | | | | 90% | 187 | 4,324 | 3.45 (2.92 to 4.09) | 4.34E-47 |
| | | | | 95% | 106 | 2,150 | 3.56 (2.90 to 4.39) | 3.24E-33 |
| | | | | 97.50% | 51 | 1,077 | 3.20 (2.41 to 4.26) | 1.43E-15 |
| | | | | 99% | 25 | 427 | 3.74 (2.51 to 5.58) | 1.01E-10 |
| | | Quartile 2 | from 51 to 58 | 80% | 420 | 8,160 | 2.28 (2.02 to 2.57) | 1.35E-41 |
| | | | | 90% | 242 | 4,048 | 2.40 (2.08 to 2.77) | 7.72E-34 |
| | | | | 95% | 137 | 2,008 | 2.60 (2.17 to 3.11) | 9.26E-26 |
| | | | | 97.50% | 74 | 999 | 2.76 (2.18 to 3.49) | 3.17E-17 |
| | | | | 99% | 34 | 395 | 3.14 (2.23 to 4.42) | 4.88E-11 |
| | | Quartile 3 | from 58 to 63 | 80% | 484 | 8,273 | 1.82 (1.63 to 2.02) | 1.25E-27 |
| | | | | 90% | 267 | 4,112 | 1.88 (1.65 to 2.15) | 6.41E-21 |
| | | | | 95% | 139 | 2,051 | 1.88 (1.58 to 2.24) | 1.18E-12 |
| | | | | 97.50% | 77 | 1,018 | 2.05 (1.63 to 2.58) | 8.29E-10 |
| | | | | 99% | 36 | 402 | 2.38 (1.71 to 3.32) | 2.69E-07 |
| | | Quartile 4 | from 63 to 73 | 80% | 658 | 8,526 | 1.95 (1.78 to 2.14) | 3.40E-45 |
| | | | | 90% | 361 | 4,231 | 1.99 (1.77 to 2.23) | 2.96E-32 |
| | | | | 95% | 197 | 2,099 | 2.08 (1.80 to 2.41) | 1.46E-22 |
| | | | | 97.50% | 96 | 1,052 | 1.96 (1.60 to 2.41) | 1.28E-10 |
| | | | | 99% | 48 | 412 | 2.46 (1.85 to 3.28) | 7.41E-10 |

**Supplemental Table 7: Net Reclassification Improvement of $PRS_{CAD}$ for MI incidence, sex and age stratified.** Net reclassification improvement for events (NRI Events), for non-events (NRI Non-Events) and overall net reclassification improvement (NRI) were determined with their 95% confidence intervals (95% CI), for a risk threshold of 2% ($NRI^{0.02}$) and for continuous NRI ($NRI^{>0}$) for all individuals, but also in women and in men. 95% CIs were calculated using bootstrapping method.

| Categorized Net Reclassification Improvement ($NRI^{0.02}$) | | | | |
|---|---|---|---|---|
| Sex Group | Age Group | $NRI^{0.02}$ Events | $NRI^{0.02}$ Non-Events | Overall $NRI^{0.02}$ |
| All | All Ages | 0.0669 (0.0535 to 0.0792) | -0.0217 (-0.0245 to -0.0180) | 0.0452 (0.0333 to 0.0573) |
| | Quartile 1 | 0.190 (0.133 to 0.242) | -0.0326 (-0.0451 to -0.0211) | 0.158 (0.111 to 0.199) |
| | Quartile 2 | 0.134 (0.0950 to 0.165) | -0.0544 (-0.0702 to -0.0327) | 0.0794 (0.0527 to 0.104) |
| | Quartile 3 | 0.0245 (0.000711 to 0.0425) | -0.00615 (-0.0129 to 0.000821) | 0.0183 (-0.00557 to 0.0365) |
| | Quartile 4 | -0.00488 (-0.0288 to 0.0101) | 0.0431 (0.0295 to 0.0637) | 0.0383 (0.0200 to 0.0536) |
| Women | All Ages | 0.0775 (0.0519 to 0.108) | -0.0159 (-0.0235 to -0.00828) | 0.0616 (0.0416 to 0.0864) |
| | Quartile 1 | 0.0198 (0.000 to 0.0673) | -0.0000562 (-0.00138 to 0.0000817) | 0.0197 (-0.0000817 to 0.0668) |
| | Quartile 2 | 0.0306 (0.00249 to 0.0686) | -0.00308 (-0.00702 to -0.00116) | 0.0275 (0.000479 to 0.0625) |
| | Quartile 3 | 0.0772 (0.0199 to 0.132) | -0.0218 (-0.0427 to -0.00767) | 0.0554 (0.00856 to 0.0923) |
| | Quartile 4 | 0.138 (0.086 to 0.184) | -0.0550 (-0.0700 to -0.0400) | 0.0828 (0.0420 to 0.123) |
| Men | All Ages | -0.0187 (-0.0345 to -0.00366) | 0.0689 (0.0548 to 0.0845) | 0.0502 (0.0385 to 0.0634) |
| | Quartile 1 | 0.331 (0.277 to 0.385) | -0.131 (-0.150 to -0.107) | 0.199 (0.157 to 0.248) |
| | Quartile 2 | -0.0917 (-0.155 to -0.00357) | 0.161 (0.0572 to 0.258) | 0.0693 (0.0263 to 0.121) |
| | Quartile 3 | -0.0522 (-0.0827 to -0.0313) | 0.190 (0.136 to 0.237) | 0.138 (0.101 to 0.162) |
| | Quartile 4 | -0.0287 (-0.0435 to -0.0150) | 0.103 (0.0723 to 0.141) | 0.0744 (0.0540 to 0.104) |
| Continuous Net Reclassification Improvement ($NRI^{>0}$) | | | | |
| All | All Ages | 0.158 (0.140 to 0.177) | 0.165 (0.152 to 0.178) | 0.324 (0.297 to 0.349) |
| | Quartile 1 | 0.287 (0.223 to 0.346) | 0.258 (0.230 to 0.290) | 0.545 (0.470 to 0.627) |
| | Quartile 2 | 0.168 (0.126 to 0.211) | 0.182 (0.149 to 0.212) | 0.349 (0.288 to 0.413) |
| | Quartile 3 | 0.124 (0.0850 to 0.157) | 0.143 (0.121 to 0.166) | 0.267 (0.216 to 0.315) |
| | Quartile 4 | 0.155 (0.123 to 0.187) | 0.147 (0.132 to 0.163) | 0.302 (0.260 to 0.345) |
| Women | All Ages | 0.137 (0.0989 to 0.176) | 0.159 (0.127 to 0.191) | 0.296 (0.235 to 0.356) |
| | Quartile 1 | 0.130 (0.00676 to 0.263) | 0.182 (0.119 to 0.248) | 0.313 (0.152 to 0.489) |
| | Quartile 2 | 0.118 (0.0317 to 0.205) | 0.140 (0.0953 to 0.189) | 0.258 (0.144 to 0.368) |
| | Quartile 3 | 0.130 (0.0520 to 0.198) | 0.129 (0.0892 to 0.162) | 0.258 (0.158 to 0.346) |
| | Quartile 4 | 0.147 (0.0937 to 0.199) | 0.152 (0.123 to 0.182) | 0.299 (0.230 to 0.360) |
| Men | All Ages | 0.172 (0.148 to 0.191) | 0.174 (0.162 to 0.187) | 0.346 (0.314 to 0.372) |
| | Quartile 1 | 0.321 (0.262 to 0.388) | 0.281 (0.251 to 0.313) | 0.602 (0.525 to 0.683) |
| | Quartile 2 | 0.188 (0.139 to 0.243) | 0.196 (0.168 to 0.224) | 0.384 (0.315 to 0.458) |
| | Quartile 3 | 0.118 (0.0807 to 0.160) | 0.152 (0.130 to 0.175) | 0.271 (0.221 to 0.326) |
| | Quartile 4 | 0.150 (0.114 to 0.187) | 0.155 (0.135 to 0.174) | 0.305 (0.257 to 0.355) |

**Supplemental Table 8: Association between $PRS_{CAD}$ and all-cause mortality in adjusted Cox regression models.** Adjusted Cox regression models including age and the first ten genetic principal components (PCs) as covariates were obtained all individuals, but also in women and in men, by quartiles of age (Q1 to Q4) and for the whole group (all age). Data are presented as estimated hazard ratios (HR) for mortality per SD increase of $PRS_{CAD}$ with their 95% confidence intervals (95% CI), and Cox regression P-value significance.

| Outcome | Sex Group | Age Group | Age Window | Max-Min Difference in Age Group | N Event | N Total | Ratio N Event / 100% of Event | HR (95% CI) | P-value |
|---|---|---|---|---|---|---|---|---|---|
| Mortality | All | All Ages | from 40 to 73 | 33 | 23,982 | 403,422 | 100.00% | 1.08 (1.06 to 1.09) | 5.46E-30 |
| Mortality | All | Quartile 1 | from 40 to 51 | 11 | 1,841 | 100,856 | 7.68% | 1.11 (1.06 to 1.16) | 6.40E-06 |
| Mortality | All | Quartile 2 | from 51 to 58 | 7 | 3,831 | 100,855 | 15.97% | 1.05 (1.01 to 1.08) | 4.22E-03 |
| Mortality | All | Quartile 3 | from 58 to 63 | 5 | 6,557 | 100,855 | 27.34% | 1.08 (1.05 to 1.10) | 2.33E-09 |
| Mortality | All | Quartile 4 | from 63 to 73 | 10 | 11,753 | 100,856 | 49.01% | 1.08 (1.06 to 1.10) | 3.78E-17 |
| Mortality | Women | All Ages | from 40 to 71 | 31 | 9,604 | 217,886 | 100.00% | 1.04 (1.02 to 1.06) | 3.17E-04 |
| Mortality | Women | Quartile 1 | from 40 to 51 | 11 | 830 | 55,289 | 8.64% | 1.07 (1.00 to 1.15) | 4.99E-02 |
| Mortality | Women | Quartile 2 | from 51 to 58 | 7 | 1,649 | 56,704 | 17.17% | 0.95 (0.91 to 1.00) | 5.41E-02 |
| Mortality | Women | Quartile 3 | from 58 to 63 | 5 | 2,651 | 54,737 | 27.60% | 1.04 (1.00 to 1.08) | 5.92E-02 |
| Mortality | Women | Quartile 4 | from 63 to 71 | 8 | 4,474 | 51,156 | 46.58% | 1.07 (1.03 to 1.10) | 2.67E-05 |
| Mortality | Men | All Ages | from 40 to 73 | 33 | 14,378 | 185,536 | 100.00% | 1.10 (1.09 to 1.12) | 6.26E-32 |
| Mortality | Men | Quartile 1 | from 40 to 51 | 11 | 1,011 | 45,567 | 7.03% | 1.15 (1.08 to 1.22) | 1.64E-05 |
| Mortality | Men | Quartile 2 | from 51 to 58 | 7 | 2,182 | 44,151 | 15.18% | 1.13 (1.08 to 1.18) | 3.36E-08 |
| Mortality | Men | Quartile 3 | from 58 to 63 | 5 | 3,906 | 46,118 | 27.17% | 1.10 (1.07 to 1.14) | 6.73E-10 |
| Mortality | Men | Quartile 4 | from 63 to 73 | 10 | 7,279 | 49,700 | 50.63% | 1.09 (1.07 to 1.12) | 1.33E-13 |

**Supplemental Table 9: Mortality in individuals at high genetic risk.** Adjusted Cox regression results were obtained for individuals belonging above percentile thresholds (ranging between the 80th and the 99th percentile). Hazard ratios (HR) for mortality and their 95% confidence intervals (95% CI) were compared in all individuals, but also separately in women and in men, for all ages and for each quartile of age (Q1 to Q4).

| Outcome | Sex Group | Age Group | Age Distribution | Percentile Thres. | N event | N total | HR (95% CI) | P-value |
|---|---|---|---|---|---|---|---|---|
| Mortality | All | All Ages | from 40 to 73 | 80% | 5,251 | 75,434 | 1.14 (1.11 to 1.18) | 1.22E-17 |
| | | | | 90% | 2,717 | 37,626 | 1.18 (1.13 to 1.23) | 6.86E-16 |
| | | | | 95% | 1,392 | 18,780 | 1.21 (1.15 to 1.28) | 3.49E-12 |
| | | | | 97.50% | 737 | 9,349 | 1.29 (1.20 to 1.39) | 8.27E-12 |
| | | | | 99% | 302 | 3,733 | 1.33 (1.18 to 1.49) | 1.07E-06 |
| | | Quartile 1 | from 40 to 51 | 80% | 444 | 19,728 | 1.28 (1.15 to 1.42) | 7.83E-06 |
| | | | | 90% | 246 | 9,840 | 1.39 (1.22 to 1.59) | 1.23E-06 |
| | | | | 95% | 137 | 4,906 | 1.54 (1.29 to 1.83) | 1.36E-06 |
| | | | | 97.50% | 75 | 2,447 | 1.67 (1.33 to 2.11) | 1.27E-05 |
| | | | | 99% | 28 | 981 | 1.52 (1.04 to 2.20) | 2.85E-02 |
| | | Quartile 2 | from 51 to 58 | 80% | 850 | 19,321 | 1.15 (1.06 to 1.24) | 3.81E-04 |
| | | | | 90% | 448 | 9,638 | 1.20 (1.09 to 1.33) | 2.18E-04 |
| | | | | 95% | 219 | 4,824 | 1.17 (1.02 to 1.34) | 2.73E-02 |
| | | | | 97.50% | 123 | 2,399 | 1.33 (1.11 to 1.59) | 2.02E-03 |
| | | | | 99% | 51 | 958 | 1.39 (1.06 to 1.84) | 1.89E-02 |
| | | Quartile 3 | from 58 to 63 | 80% | 1,400 | 18,771 | 1.09 (1.03 to 1.16) | 3.43E-03 |
| | | | | 90% | 709 | 9,377 | 1.09 (1.01 to 1.18) | 2.26E-02 |
| | | | | 95% | 376 | 4,667 | 1.17 (1.05 to 1.30) | 3.18E-03 |
| | | | | 97.50% | 196 | 2,326 | 1.21 (1.05 to 1.40) | 7.58E-03 |
| | | | | 99% | 81 | 928 | 1.27 (1.02 to 1.58) | 3.32E-02 |
| | | Quartile 4 | from 63 to 73 | 80% | 2,589 | 17,583 | 1.15 (1.10 to 1.20) | 4.87E-10 |
| | | | | 90% | 1,331 | 8,755 | 1.18 (1.11 to 1.25) | 2.33E-08 |
| | | | | 95% | 681 | 4,362 | 1.20 (1.11 to 1.29) | 5.37E-06 |
| | | | | 97.50% | 357 | 2,165 | 1.26 (1.13 to 1.40) | 2.01E-05 |
| | | | | 99% | 153 | 856 | 1.33 (1.14 to 1.56) | 4.13E-04 |

| Outcome | Sex Group | Age Group | Age Distribution | Percentile Thres. | N event | N total | HR (95% CI) | P-value |
|---|---|---|---|---|---|---|---|---|
| Mortality | Women | All Ages | from 40 to 71 | 80% | 2,044 | 41,534 | 1.09 (1.04 to 1.15) | 3.46E-04 |
| | | | | 90% | 1,081 | 20,708 | 1.16 (1.09 to 1.24) | 3.76E-06 |
| | | | | 95% | 559 | 10,336 | 1.20 (1.10 to 1.30) | 3.78E-05 |
| | | | | 97.50% | 300 | 5,148 | 1.30 (1.16 to 1.45) | 9.82E-06 |
| | | | | 99% | 119 | 2,060 | 1.28 (1.07 to 1.54) | 6.83E-03 |
| | | Quartile 1 | from 40 to 51 | 80% | 198 | 10,860 | 1.27 (1.08 to 1.49) | 3.72E-03 |
| | | | | 90% | 108 | 5,421 | 1.36 (1.11 to 1.66) | 3.19E-03 |
| | | | | 95% | 59 | 2,706 | 1.46 (1.12 to 1.90) | 5.03E-03 |
| | | | | 97.50% | 35 | 1,348 | 1.73 (1.23 to 2.43) | 1.48E-03 |
| | | | | 99% | 15 | 538 | 1.84 (1.10 to 3.07) | 1.92E-02 |
| | | Quartile 2 | from 51 to 58 | 80% | 335 | 11,006 | 1.02 (0.906 to 1.15) | 7.25E-01 |
| | | | | 90% | 177 | 5,494 | 1.09 (0.930 to 1.27) | 2.96E-01 |
| | | | | 95% | 87 | 2,749 | 1.05 (0.849 to 1.31) | 6.35E-01 |
| | | | | 97.50% | 49 | 1,369 | 1.20 (0.900 to 1.59) | 2.16E-01 |
| | | | | 99% | 18 | 550 | 1.10 (0.693 to 1.75) | 6.81E-01 |
| | | Quartile 3 | from 58 to 63 | 80% | 545 | 10,403 | 1.04 (0.946 to 1.14) | 4.24E-01 |
| | | | | 90% | 278 | 5,196 | 1.06 (0.936 to 1.20) | 3.59E-01 |
| | | | | 95% | 148 | 2,589 | 1.14 (0.963 to 1.34) | 1.29E-01 |
| | | | | 97.50% | 77 | 1,292 | 1.19 (0.949 to 1.49) | 1.32E-01 |
| | | | | 99% | 31 | 517 | 1.19 (0.838 to 1.70) | 3.27E-01 |
| | | Quartile 4 | from 63 to 71 | 80% | 969 | 9,263 | 1.12 (1.04 to 1.20) | 1.75E-03 |
| | | | | 90% | 519 | 4,597 | 1.21 (1.10 to 1.32) | 5.46E-05 |
| | | | | 95% | 268 | 2,290 | 1.23 (1.09 to 1.40) | 8.19E-04 |
| | | | | 97.50% | 146 | 1,133 | 1.34 (1.14 to 1.58) | 4.97E-04 |
| | | | | 99% | 58 | 454 | 1.31 (1.01 to 1.69) | 4.30E-02 |

| Outcome | Sex Group | Age Group | Age Distribution | Percentile Thres. | N event | N total | HR (95% CI) | P-value |
|---|---|---|---|---|---|---|---|---|
| Mortality | Men | All Ages | from 40 to 73 | 80% | 3,216 | 33,892 | 1.18 (1.13 to 1.22) | 3.44E-16 |
| | | | | 90% | 1,642 | 16,912 | 1.19 (1.13 to 1.26) | 2.05E-11 |
| | | | | 95% | 830 | 8,447 | 1.21 (1.13 to 1.30) | 9.32E-08 |
| | | | | 97.50% | 442 | 4,197 | 1.30 (1.18 to 1.43) | 7.26E-08 |
| | | | | 99% | 184 | 1,672 | 1.35 (1.17 to 1.56) | 4.62E-05 |
| | | Quartile 1 | from 40 to 51 | 80% | 247 | 8,867 | 1.30 (1.12 to 1.50) | 4.06E-04 |
| | | | | 90% | 137 | 4,420 | 1.42 (1.19 to 1.70) | 1.35E-04 |
| | | | | 95% | 77 | 2,202 | 1.58 (1.25 to 2.00) | 1.13E-04 |
| | | | | 97.50% | 40 | 1,100 | 1.62 (1.18 to 2.23) | 2.66E-03 |
| | | | | 99% | 15 | 441 | 1.48 (0.89 to 2.47) | 1.30E-01 |
| | | Quartile 2 | from 51 to 58 | 80% | 512 | 8,319 | 1.23 (1.12 to 1.36) | 3.44E-05 |
| | | | | 90% | 272 | 4,144 | 1.29 (1.14 to 1.47) | 8.27E-05 |
| | | | | 95% | 131 | 2,077 | 1.23 (1.03 to 1.47) | 2.19E-02 |
| | | | | 97.50% | 77 | 1,027 | 1.47 (1.17 to 1.85) | 8.50E-04 |
| | | | | 99% | 33 | 409 | 1.59 (1.13 to 2.24) | 8.33E-03 |
| | | Quartile 3 | from 58 to 63 | 80% | 857 | 8,367 | 1.13 (1.05 to 1.22) | 1.72E-03 |
| | | | | 90% | 427 | 4,185 | 1.11 (1.00 to 1.23) | 4.26E-02 |
| | | | | 95% | 228 | 2,078 | 1.19 (1.04 to 1.36) | 1.03E-02 |
| | | | | 97.50% | 117 | 1,036 | 1.22 (1.02 to 1.47) | 3.34E-02 |
| | | | | 99% | 51 | 411 | 1.34 (1.01 to 1.76) | 3.96E-02 |
| | | Quartile 4 | from 63 to 73 | 80% | 1,633 | 8,307 | 1.17 (1.11 to 1.24) | 2.41E-08 |
| | | | | 90% | 823 | 4,147 | 1.16 (1.08 to 1.25) | 4.55E-05 |
| | | | | 95% | 428 | 2,057 | 1.20 (1.09 to 1.32) | 3.09E-04 |
| | | | | 97.50% | 217 | 1,026 | 1.22 (1.06 to 1.39) | 4.30E-03 |
| | | | | 99% | 95 | 402 | 1.34 (1.10 to 1.64) | 4.35E-03 |
