## Supplemental Figures for "A Polygenic Risk Score for Coronary Artery Disease Improves the Prediction of Early-Onset Myocardial Infarction and Mortality in Men"

### **SUPPLEMENTAL TABLES**

**Supplemental Table 1: Characteristics of the cohort.** ApoB indicates Apolipoprotein B; ASCVD, Atherosclerotic Cardiovascular Disease; BMI, Body Mass Index; HDL-C, High Density Lipoprotein Cholesterol; HES, Hospital Episode Statistics; IQR, Interquartile Range; LDL-C, Low Density Lipoprotein Cholesterol; Lp(a), Lipoprotein (a); MI, Myocardial Infarction; PCE, Pooled Cohort Equation; SD, Standard Deviation; TC, Total Cholesterol

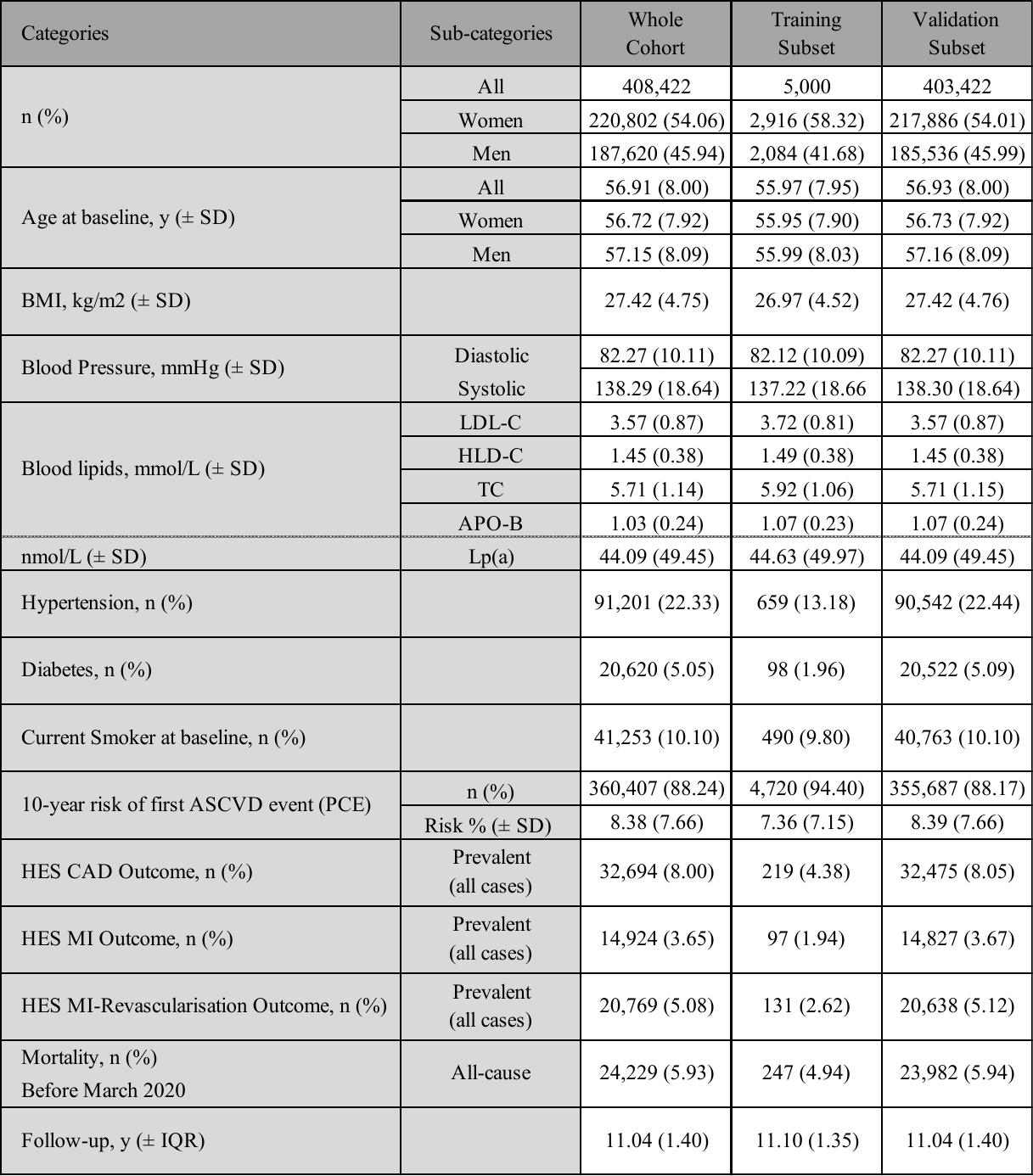

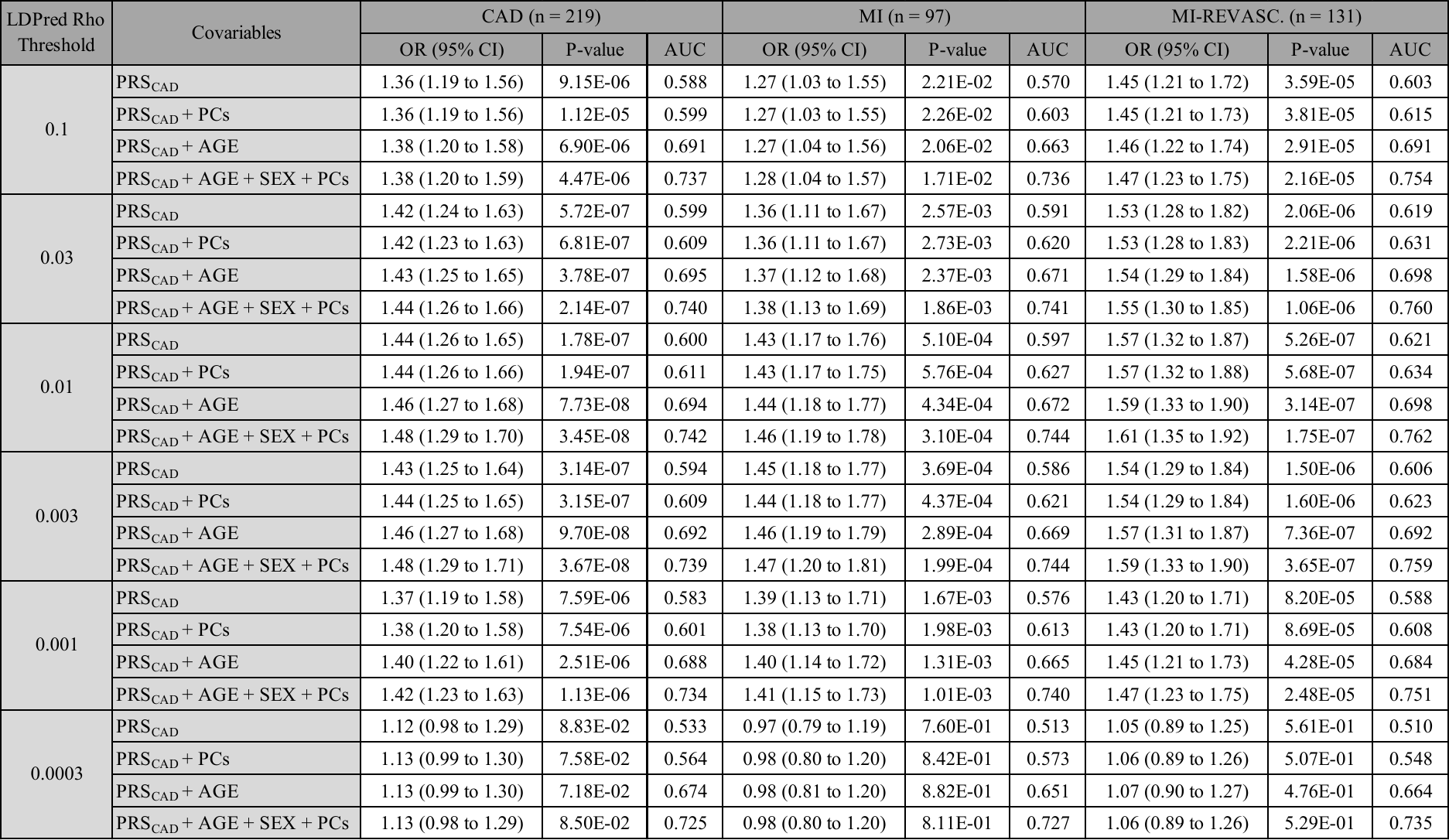
**Supplemental Table 2: Association between PRS_CAD_ and prevalent CAD in the training dataset.** Non-adjusted and adjusted models are used in logistic regression with prevalent cardiovascular outcomes. Data are presented as logistic-regression odd ratios (OR) per SD increase of PRS_CAD_ with their 95% confidence intervals (95% CI), logistic regression P-value significance and corresponding area under the receiver operating characteristic curve (AUC) values. MI, Myocardial Infarction; MI-REVASC: MI or Revascularisation Procedure; PRS_CAD_, Polygenic Risk Score for Coronary Artery Disease; PCE, Pooled Cohort Equation; PCs, First Ten Genetic Principal Components; CI: Confidence Interval

**Supplemental Table 3: Association between PRS_CAD_ and prevalent cardiovascular outcomes in mutlivariable models.** Non-adjusted and adjusted models are used in logistic regression with CAD, MI and MI-REVASC. outcomes. Data are presented as logistic-regression odd ratios (OR) per SD increase of PRS_CAD_ and their 95% confidence intervals (95% CI), Nagelkerke's pseudo-R^2^ corresponding area under the receiver operating characteristic curve (AUC) values.

BMI, Body Mass Index; HDL-C, High Density Lipoprotein Cholesterol; LDL-C, Low Density Lipoprotein Cholesterol; Lp(a), Lipoprotein (a); MI, Myocardial Infarction; MI-REVASC: MI or Revascularisation Procedure; PCE, Pooled Cohort Equation; PCs, First Ten Genetic Principal Components; PRS_CAD_, Polygenic Risk Score for Coronary Artery Disease; TC, Total Cholesterol

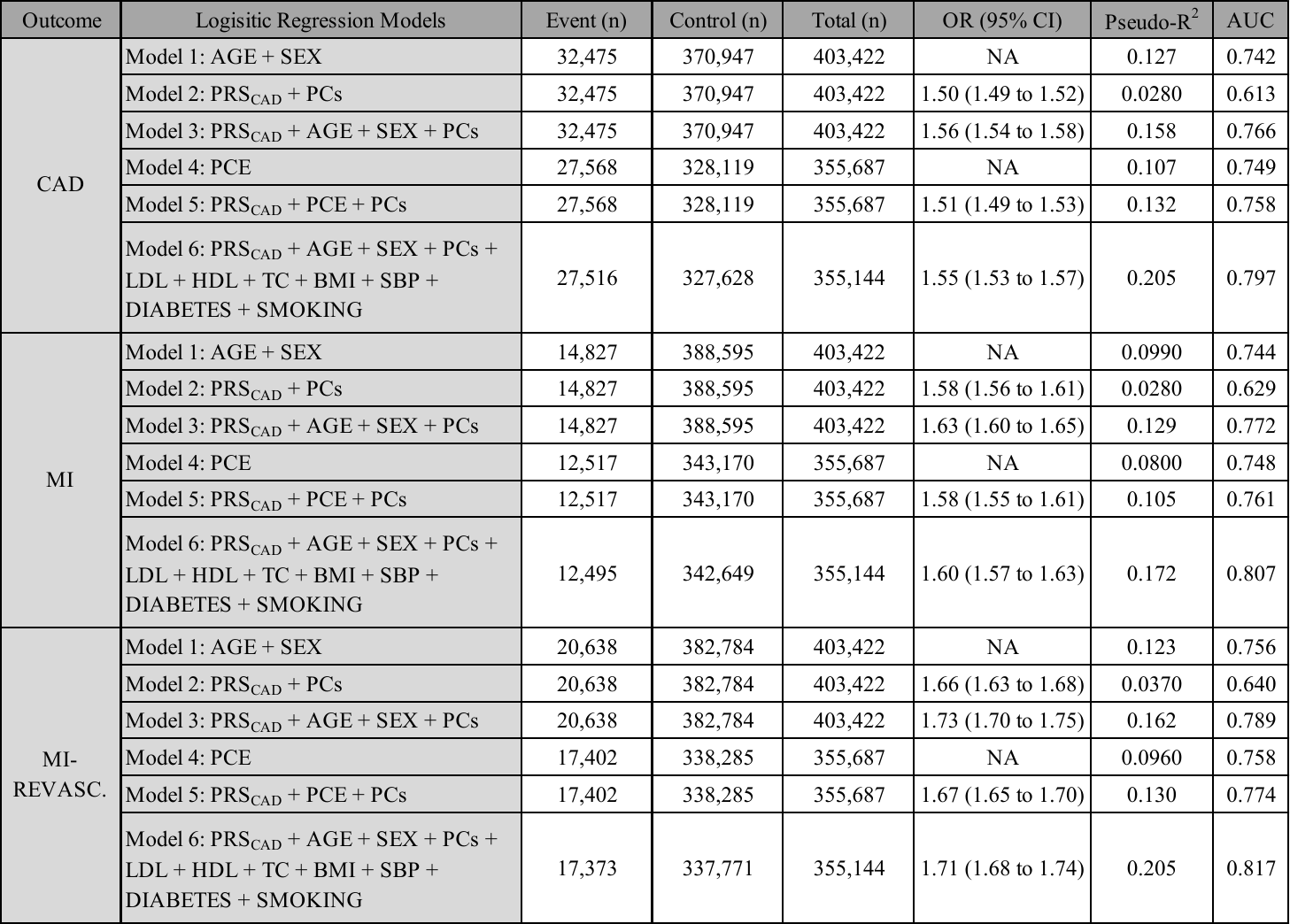

**Supplemental Table 4: Association between PRS_CAD_ and MI incidence in adjusted Cox regression models.** Adjusted Cox regression results from PRS_CAD_ models including age and the first ten genetic principal components (PCs) as covariates were obtained in women and men, by quartiles of age (Q1 to Q4) and for the whole group (all age). Data are presented as estimated hazard ratios (HR) for incident MI outcome per SD increase of PRS_CAD_, with their 95% confidence intervals (95% CI), and Cox regression P-value significance.

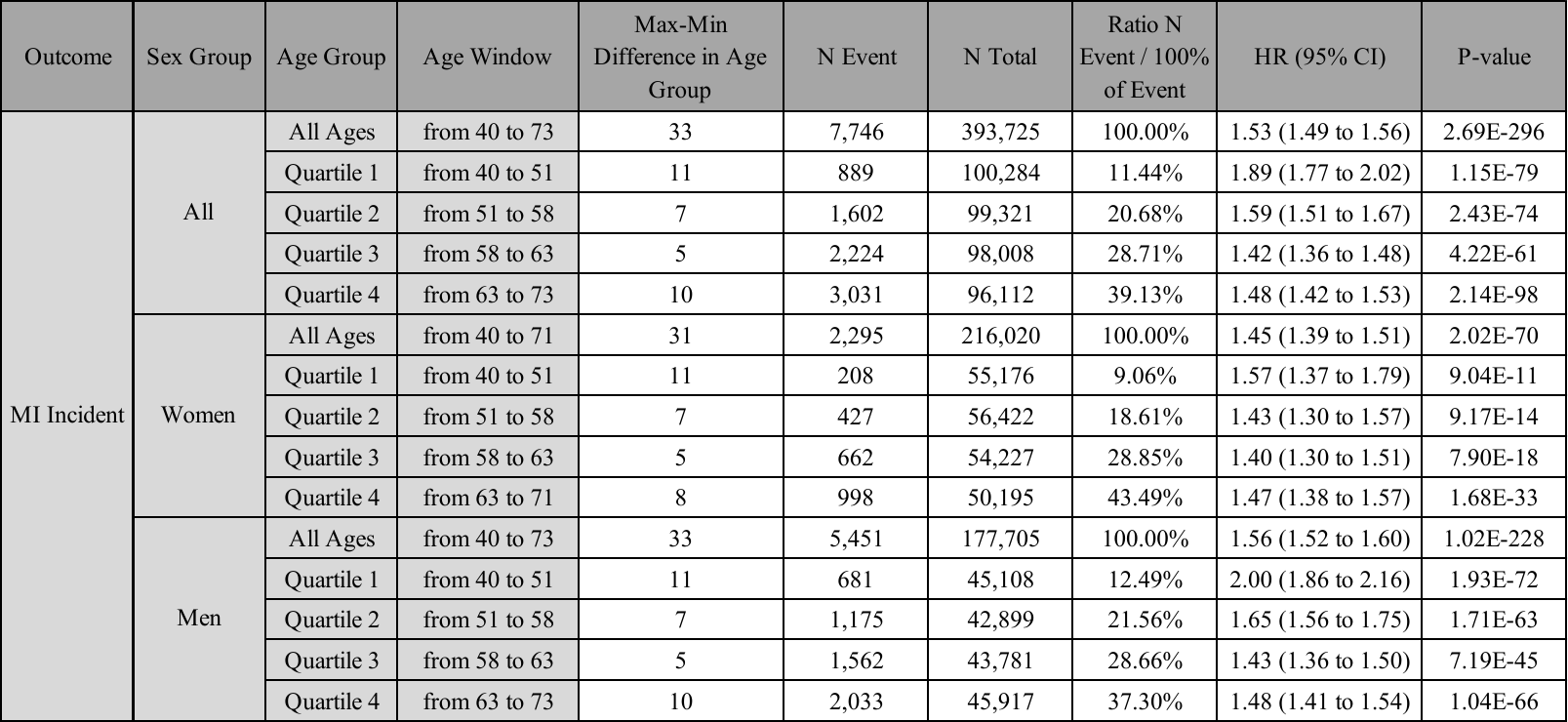

**Supplemental Table 5: Comparison between adjusted Cox regression models for MI incidence including PCE and PRS_CAD_.** Data are presented as estimated hazard ratios (HR) for incident MI outcome per SD increase of PRS_CAD_, with their 95% confidence intervals (95% CI), and Cox regression p-value significance. *Sex adjustment was only performed in “all” group.

MI: Myocardial Infarction; PRS_CAD_: Polygenic Risk Score for Coronary Artery Disease; PCE: Pooled Cohort Equation; PCs: First Ten Genetic Principal Components

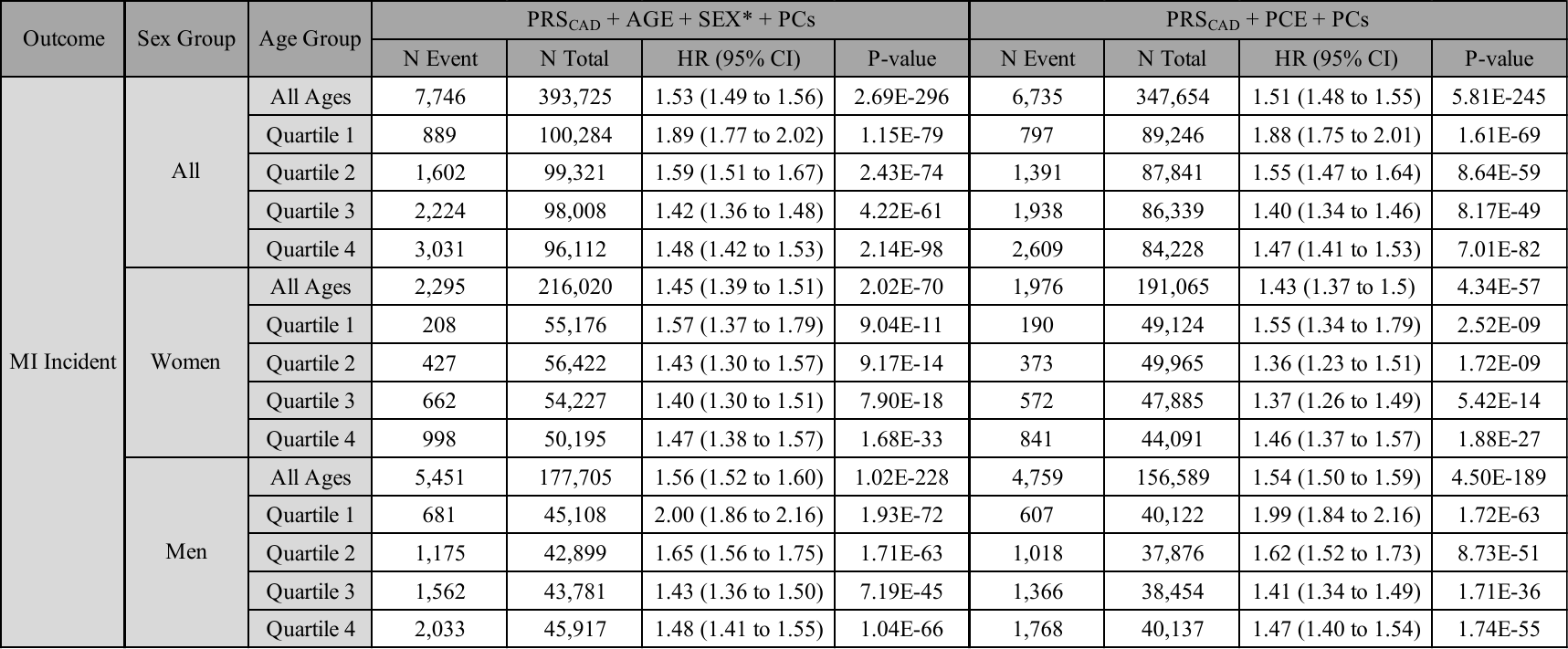

**Supplemental Table 6: MI incidence in individuals at high genetic risk.** Adjusted Cox regression results were obtained for individuals above percentile thresholds (ranging between the 80^th^ and the 99^th^ percentile). Hazard ratios (HR) for MI incidence and their 95% confidence intervals (95% CI) were compared in all individuals, and separately in women and in men, for all ages and for each quartile of age (Q1 to Q4).

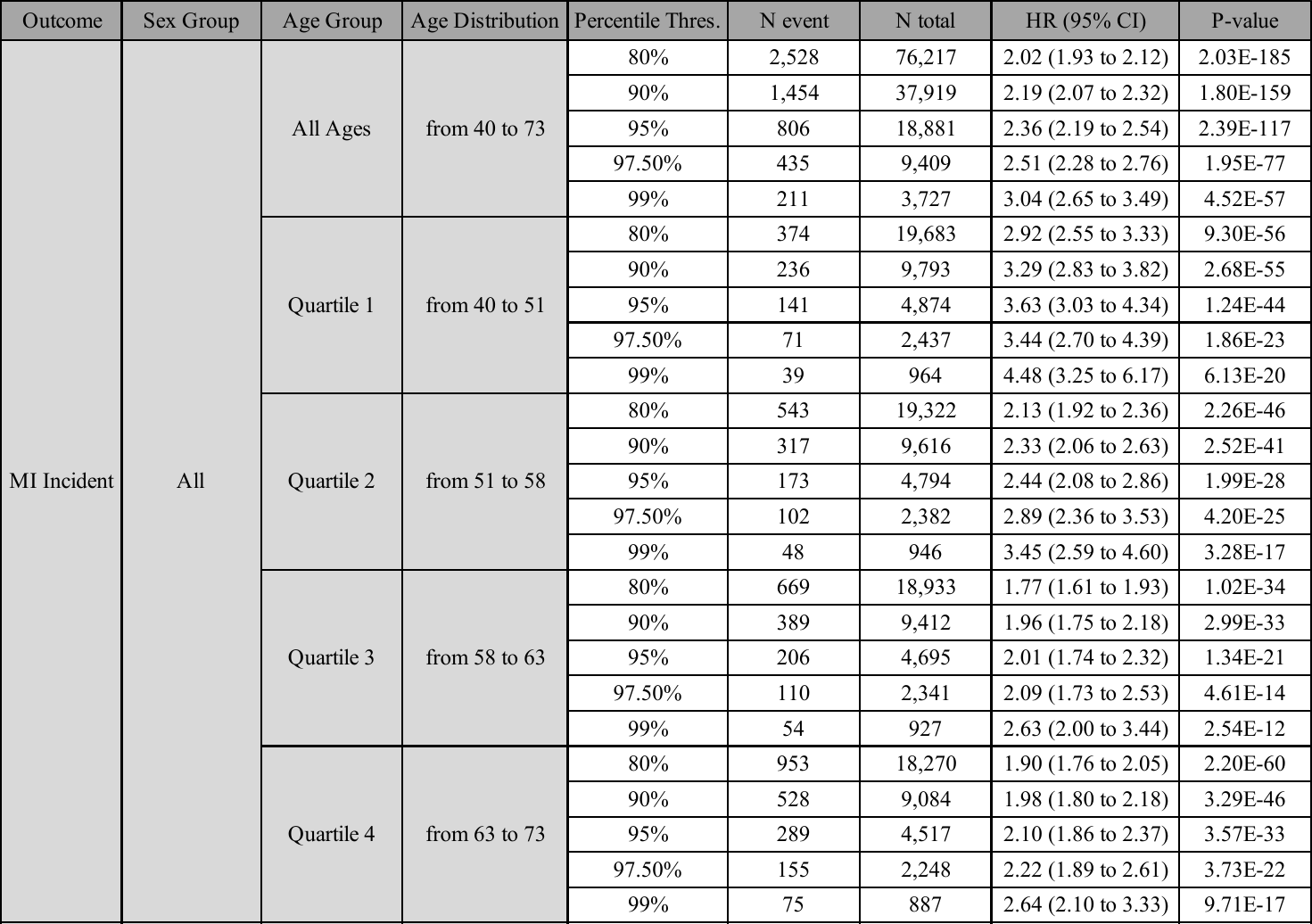

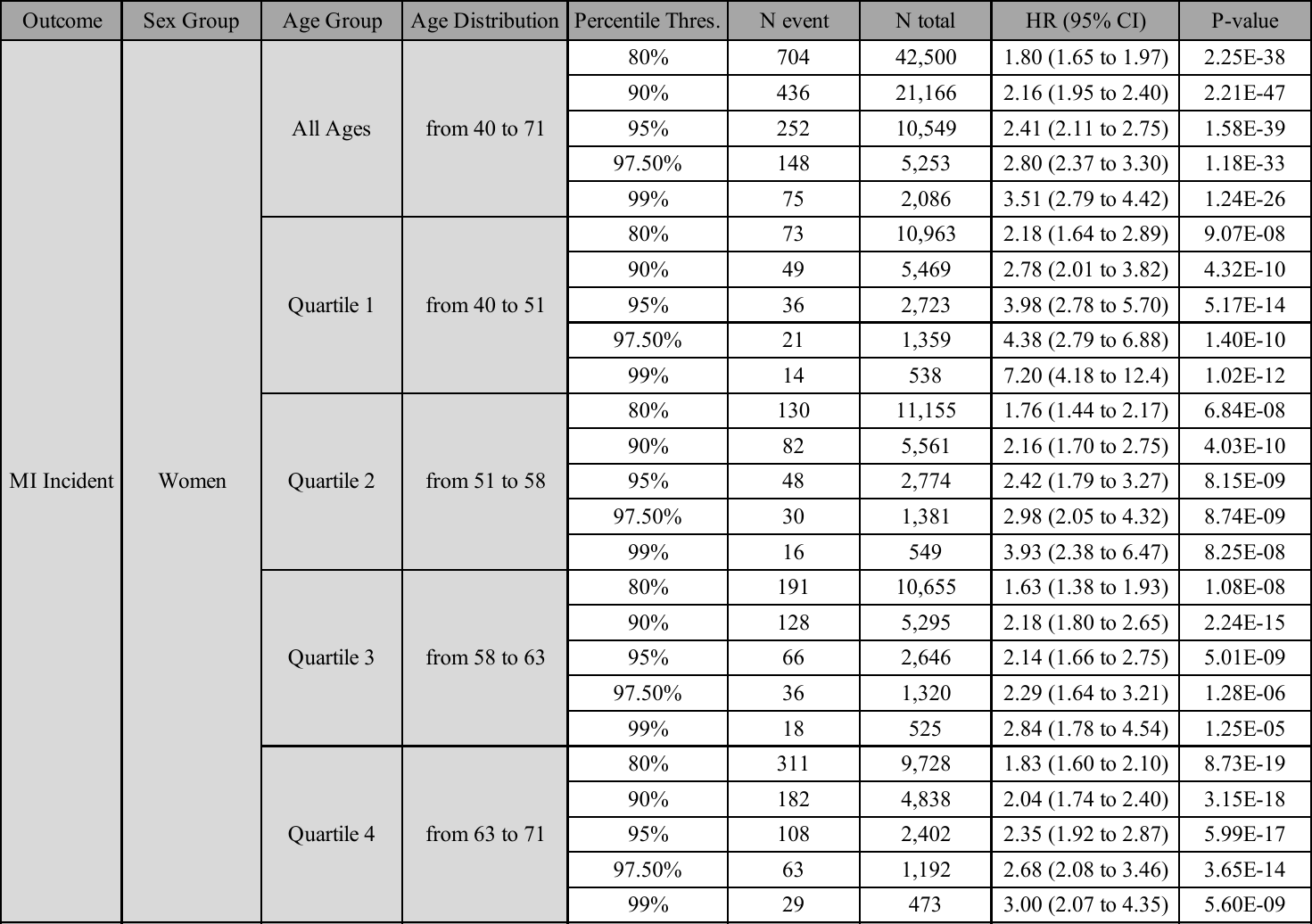

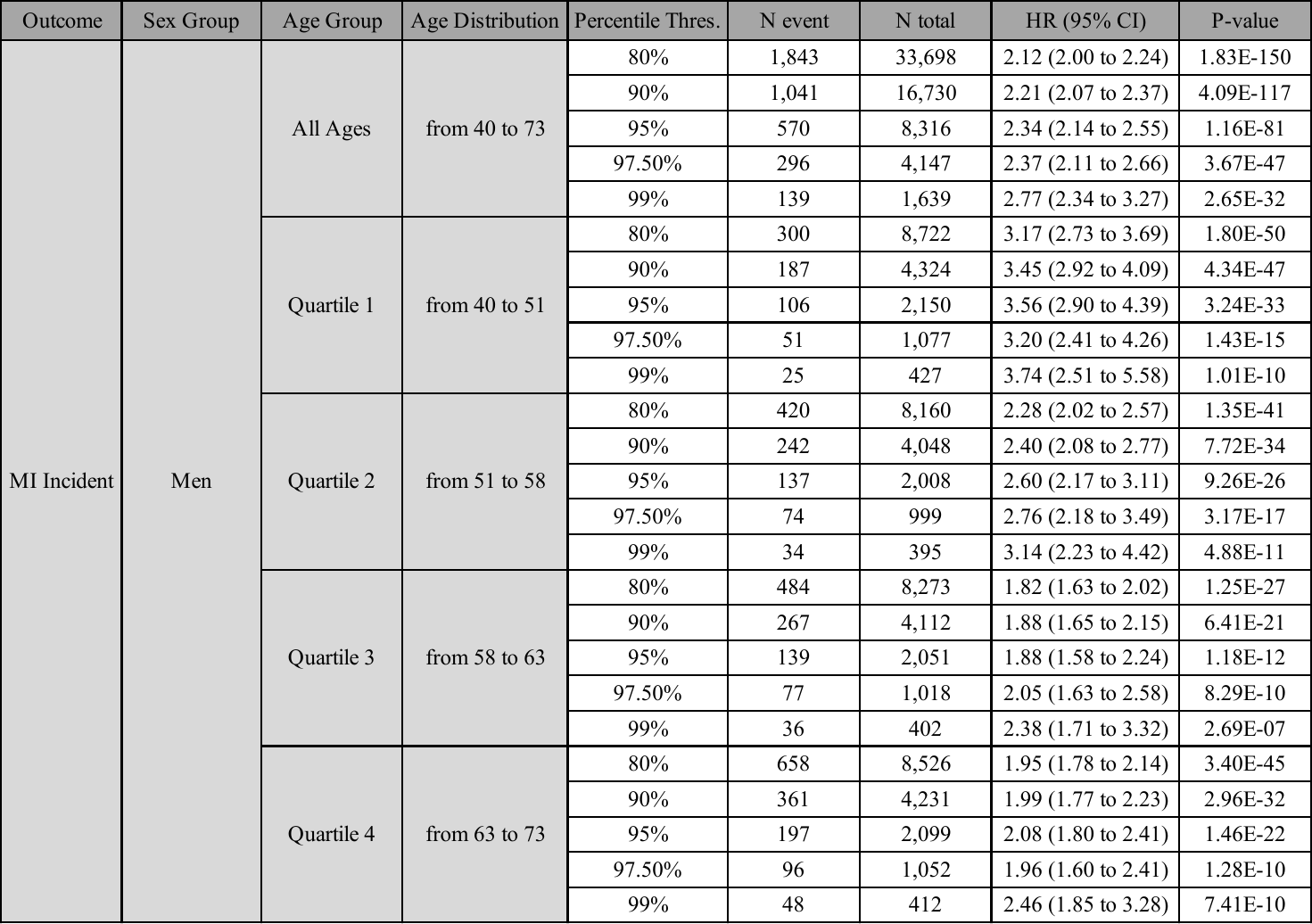

**Supplemental Table 7: Net Reclassification Improvement of PRS_CAD_ for MI incidence, sex and age stratified.** Net reclassification improvement for events (NRI Events), for non-events (NRI Non-Events) and overall net reclassification improvement (NRI) were determined with their 95% confidence intervals (95% CI), for a risk threshold of 2% (NRI^0.02^) and for continuous NRI (NRI^>0^) for all individuals, but also in women and in men. 95% CIs were calculated using bootstrapping method.

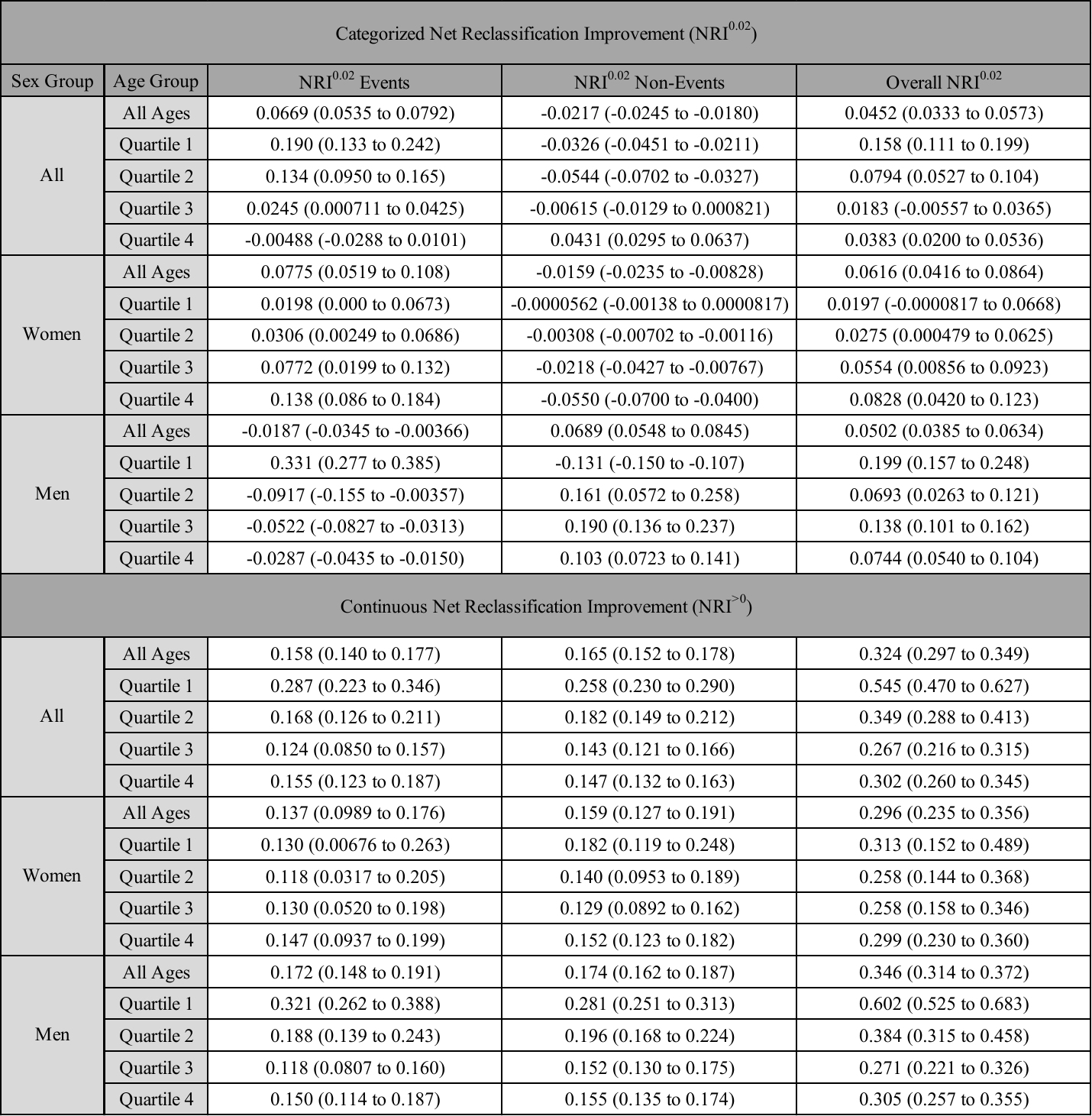

**Supplemental Table 8: Association between PRS_CAD_ and all-cause mortality in adjusted Cox regression models.** Adjusted Cox regression models including age and the first ten genetic principal components (PCs) as covariates were obtained all individuals, but also in women and in men, by quartiles of age (Q1 to Q4) and for the whole group (all age). Data are presented as estimated hazard ratios (HR) for mortality per SD increase of PRS_CAD_ with their 95% confidence intervals (95% CI), and Cox regression P-value significance.

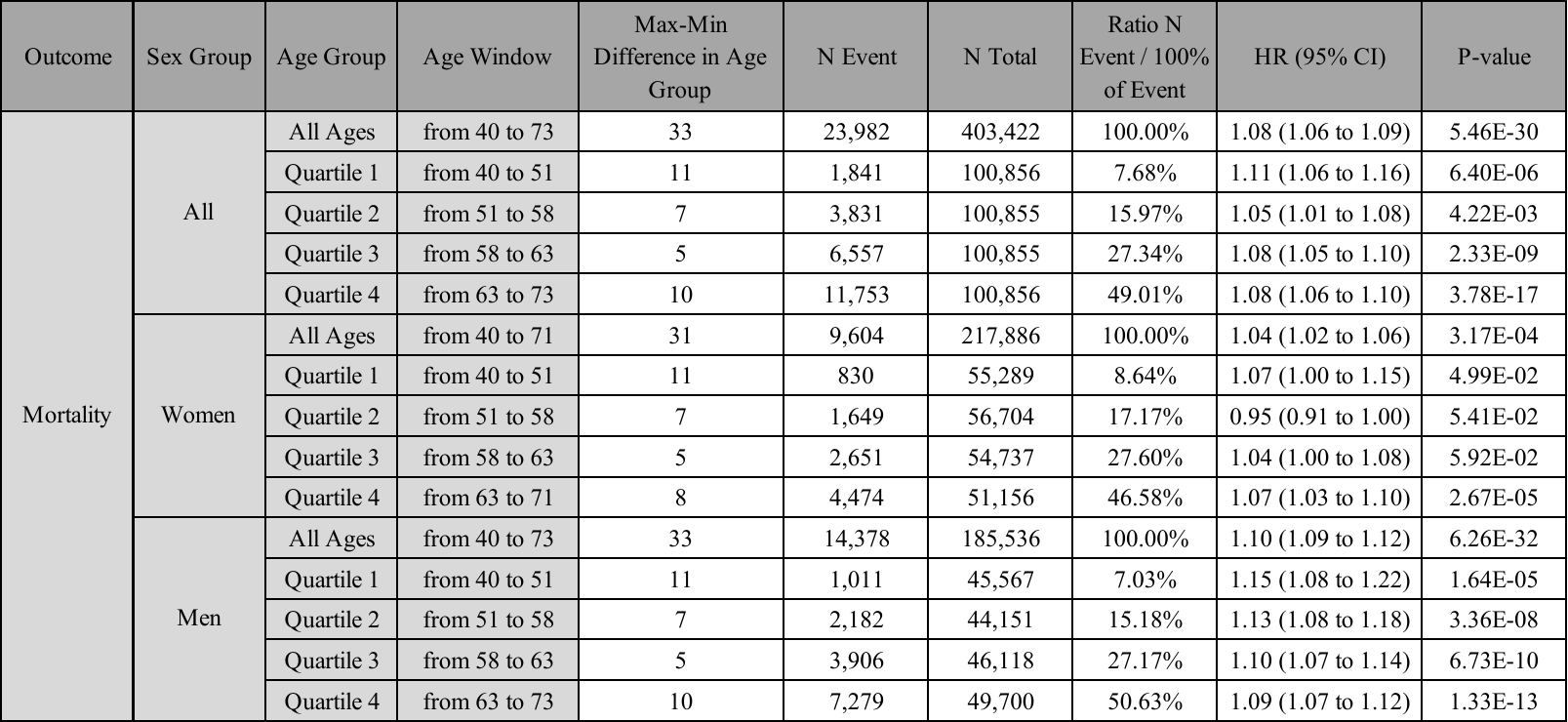

**Supplemental Table 9: Mortality in individuals at high genetic risk.** Adjusted Cox regression results were obtained for individuals belonging above percentile thresholds (ranging between the 80^th^ and the 99^th^ percentile). Hazard ratios (HR) for mortality and their 95% confidence intervals (95% CI) were compared in all individuals, but also separately in women and in men, for all ages and for each quartile of age (Q1 to Q4).

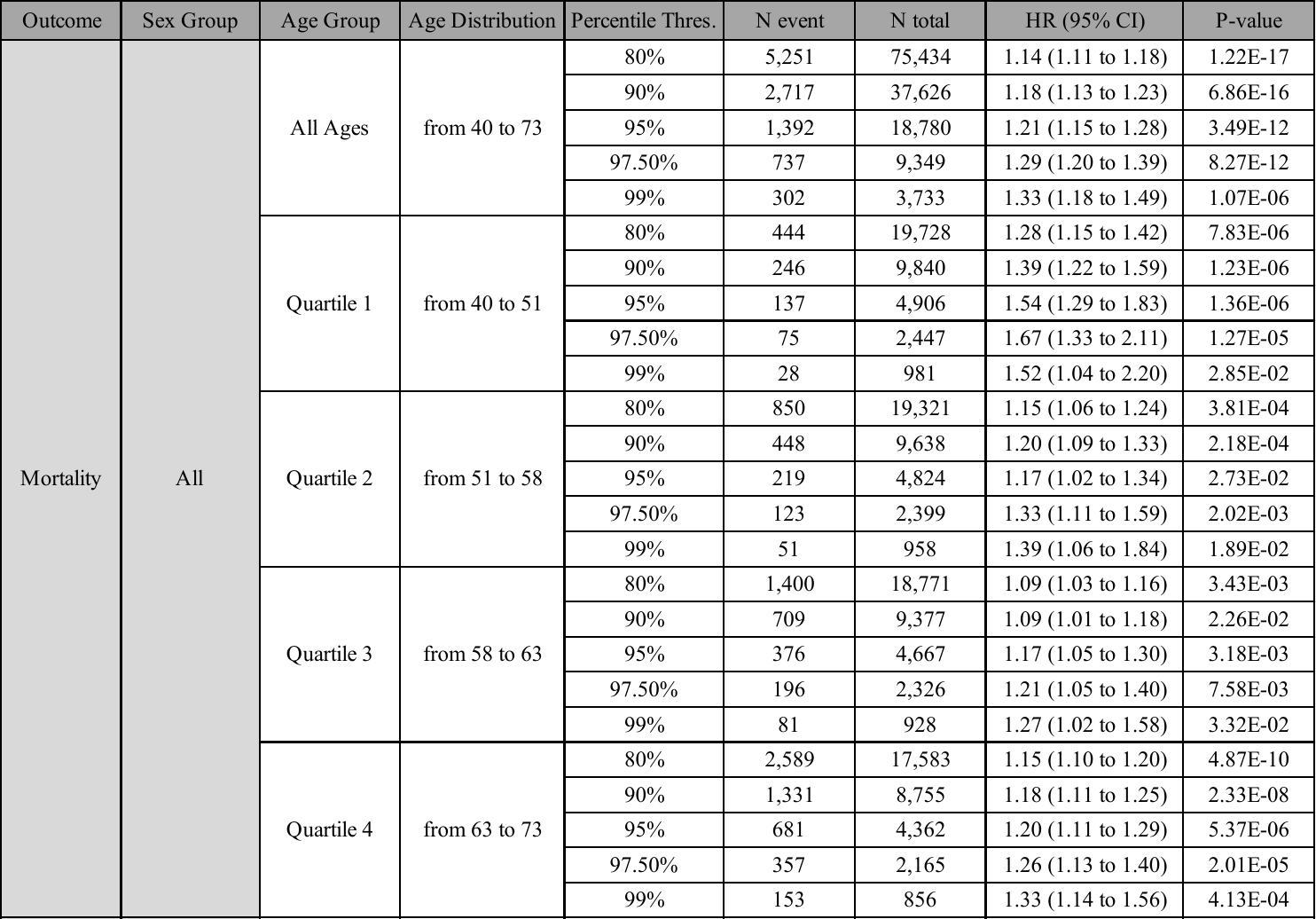

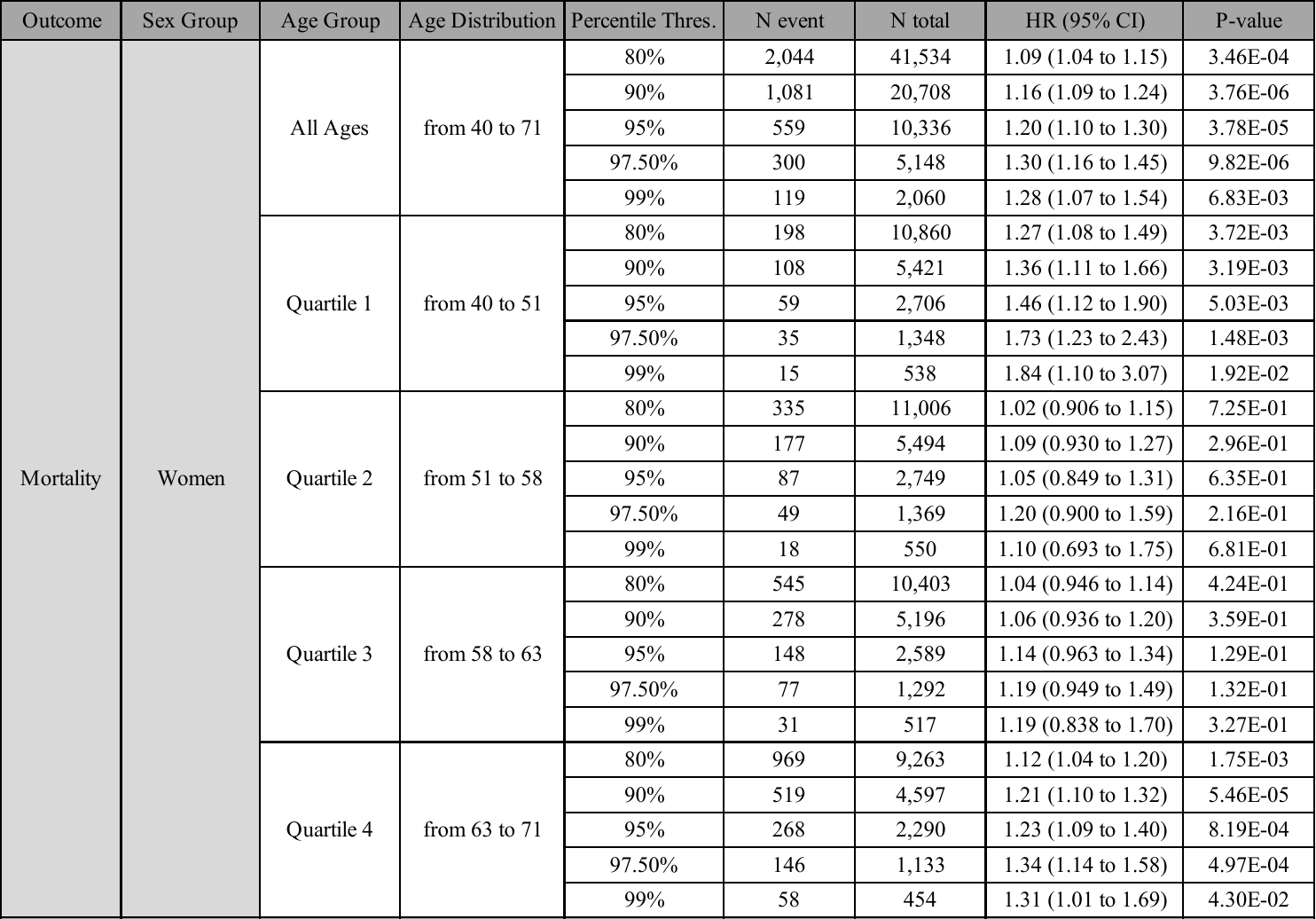

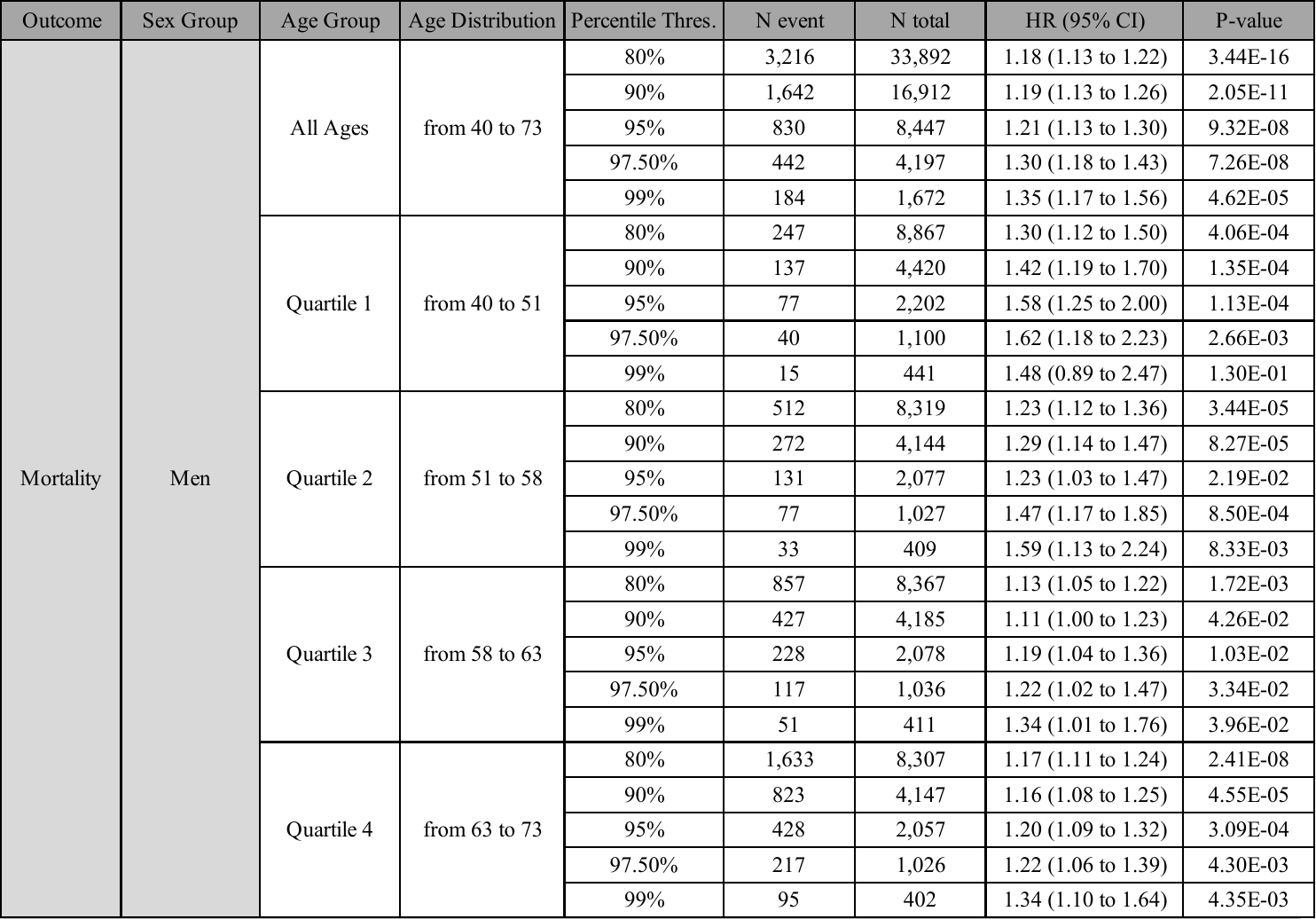

### **SUPPLEMENTAL FIGURES**

**Supplemental Figure 1: Incidence of first MI distribution.** Data are presented as counts of incident MI over time until August 2020. Censoring date was determined when the decrease of a month count exceeded 60% of the mean of the previous three months (within the two last years).

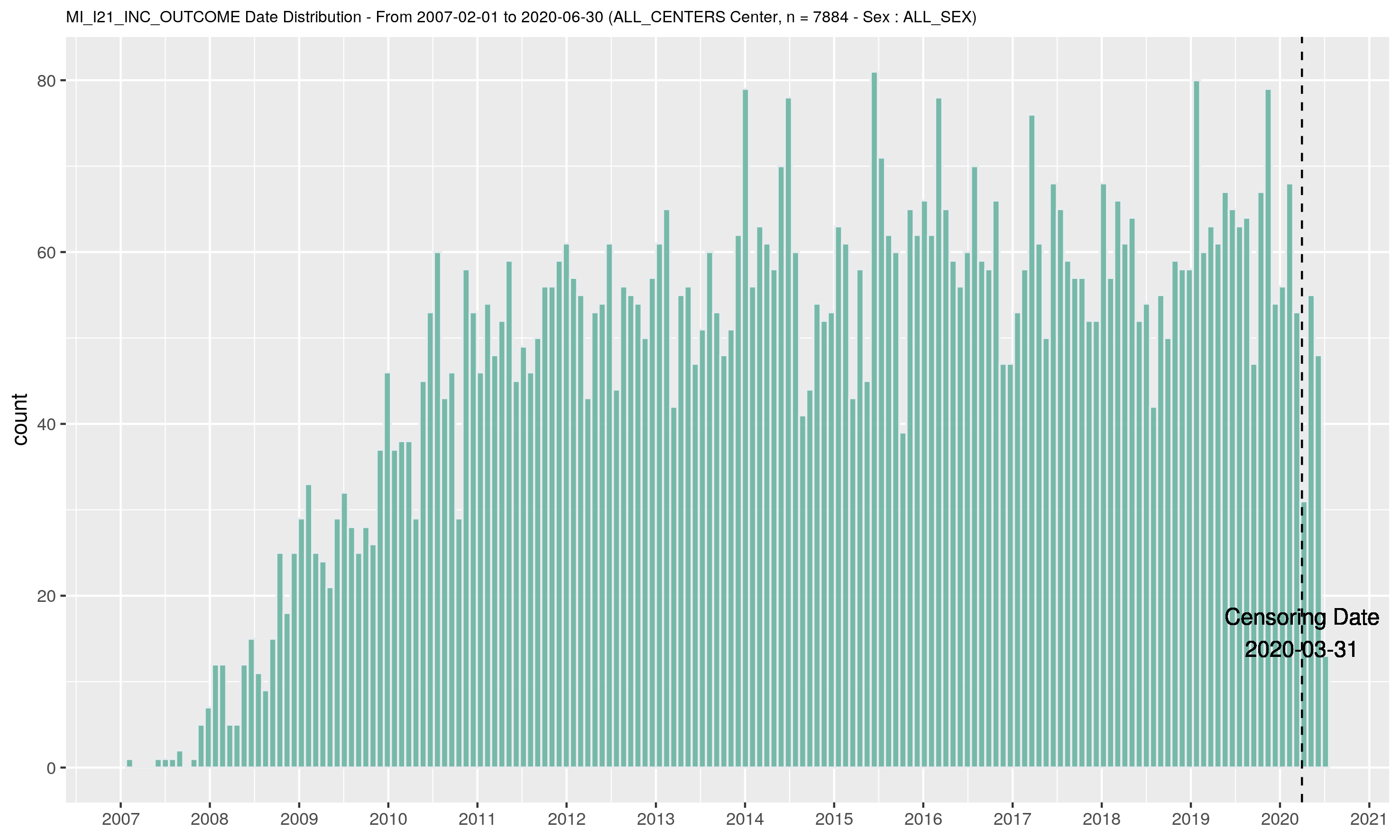

**Supplemental Figure 2: All-cause mortality distribution.** Data are presented as counts of deaths over time until September 2020. Censoring date was determined before the sharp increase in mortality, likely secondary to the COVID-19 pandemic.

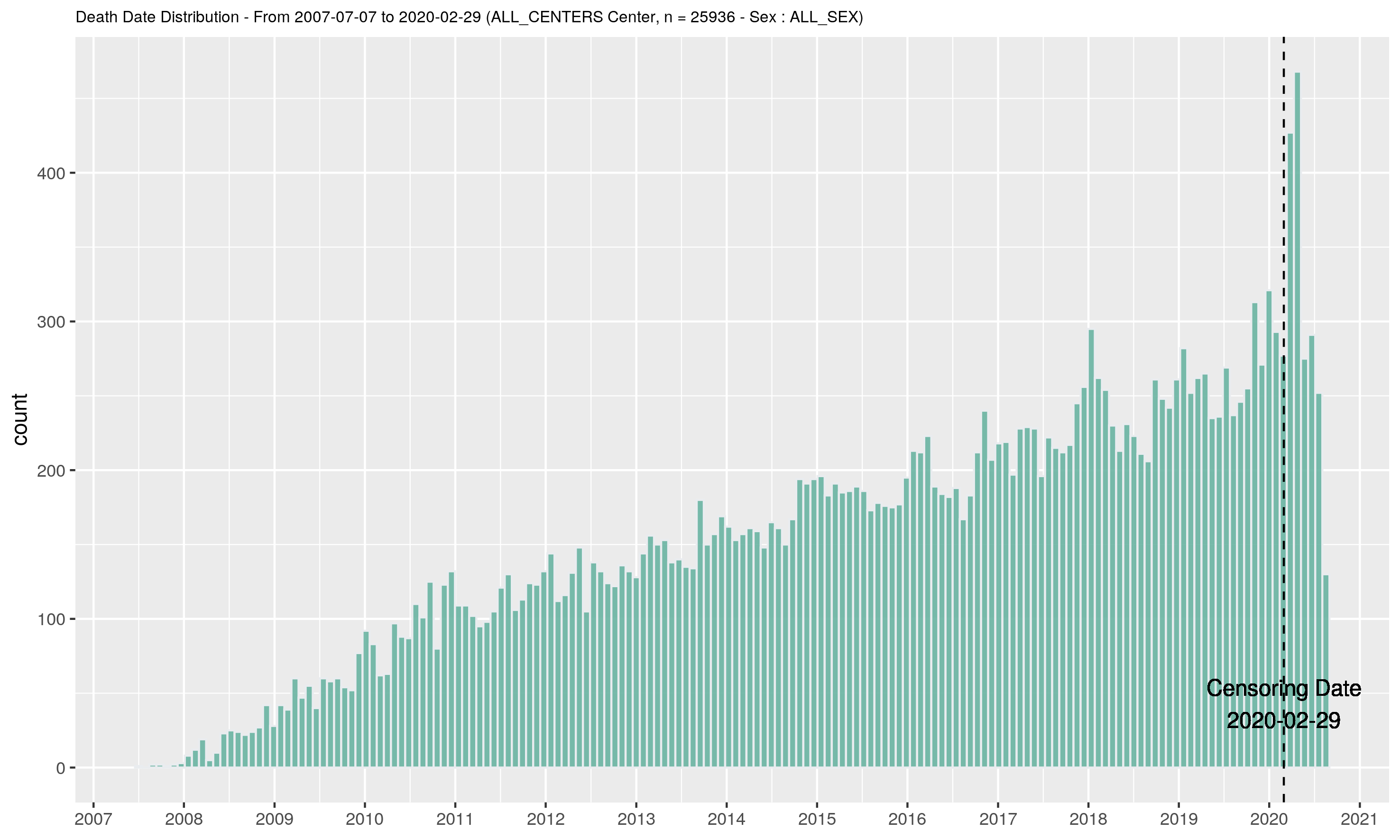

**Supplemental Figure 3: Frequency distribution of PRS_CAD_ in the validation group.** Anderson-Darling Normality Test, p-value = 0.1468.

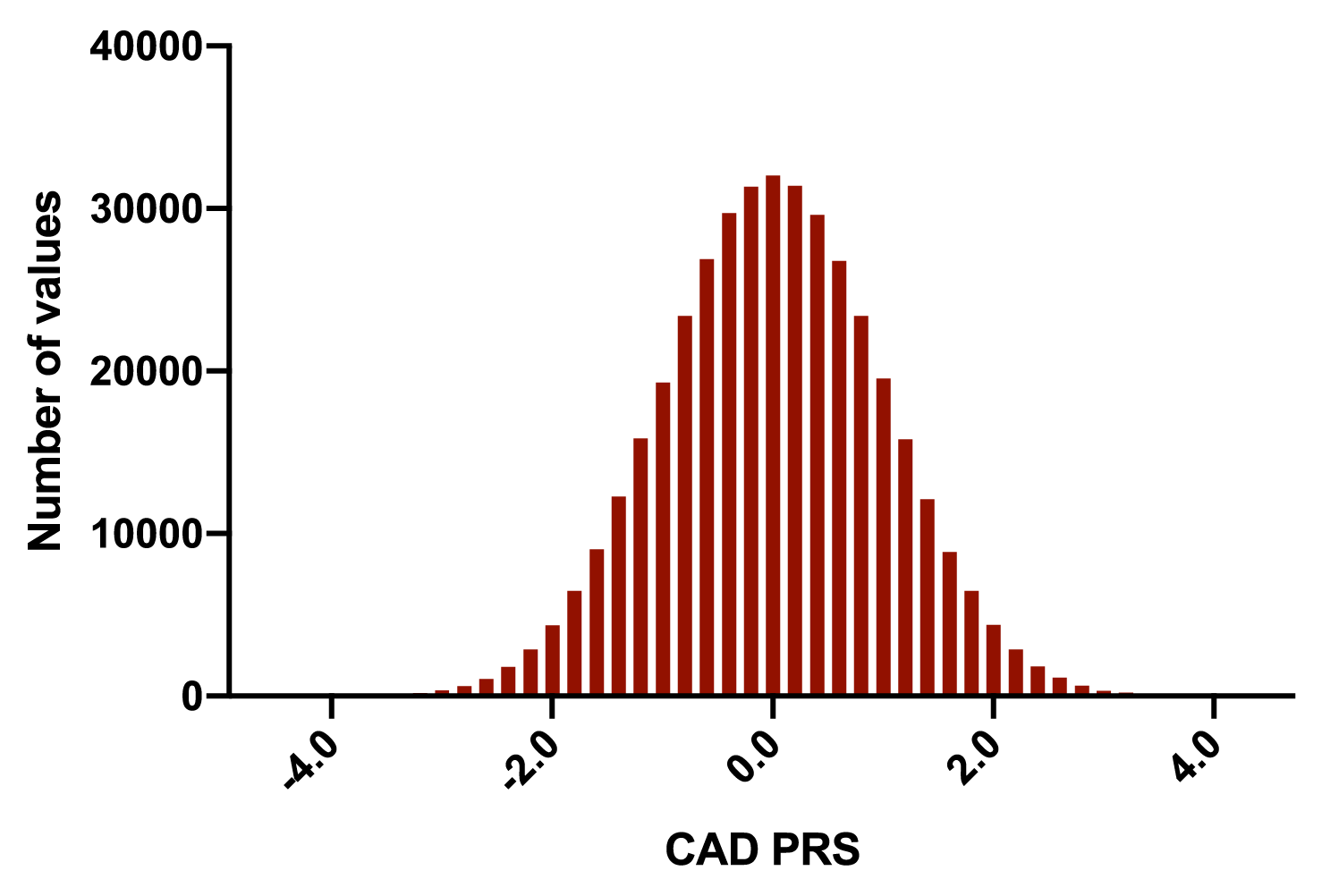

**Supplemental Figure 4: Comparisons of mean PRS_CAD_ for each age quartile.** Mean PRS_CAD_ (+/- SEM) are presented for each age quartile (Q1 to Q4). Two-sided P-values given for T-test: ** ; p < 0.01, *** ; p < 0.001).

SEM: Standard Error of the Mean

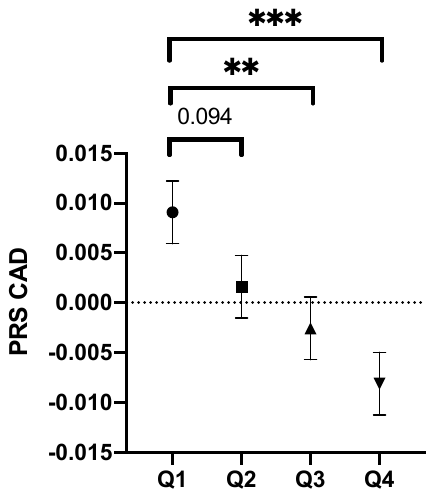

**Supplemental Figure 5: Correlation between age at first incident MI and PRS_CAD_, in women and men.** Correlation between age at first incident MI and PRS_CAD_. Pearson’s R value of -0.094 (95% CI [-0.116 to -0.0719], p = 1.14e-16) for all individuals. Men: Pearson’s R value of -0.125 (95% CI [-0.151 to -0.0992], p = 1.44e-20; women: Pearson’s R value = -0.0162, 95% CI [-0.0571 to 0.0247], p = 0.44).

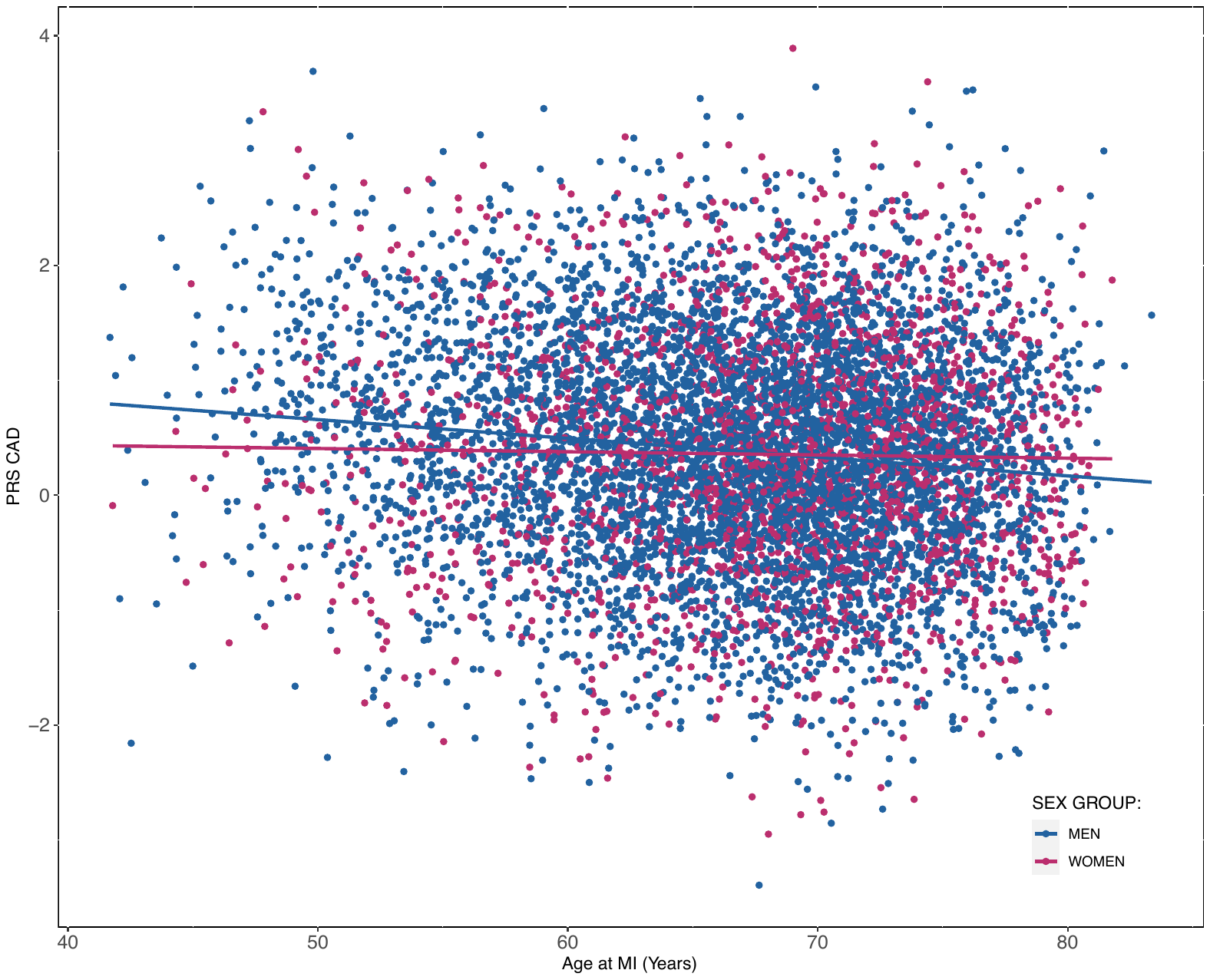

**Supplemental Figure 6: Event free probability for incident MI according to genetic risk in sex-stratified analysis.** Event-free probability curves for MI were obtained from Kaplan-Meier estimates for PRS_CAD_ tertiles (low, intermediate, high) separately in women and men. Only individuals without an event before recruitment were included. The median follow-up was 11.0 years. Log-rank test, p-value <0.0001

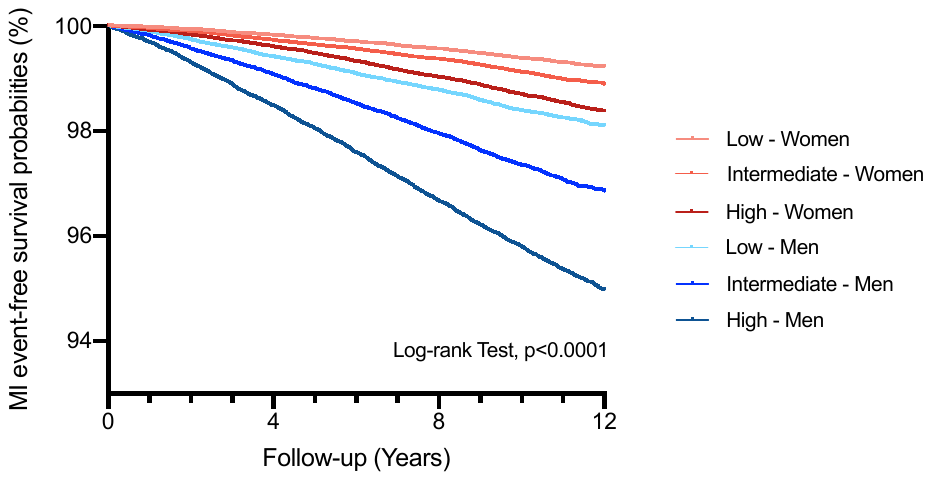

**Supplemental Figure 7: Impact of PRS_CAD_ on event-free probability for incident MI.** Event-free probabilities curves for MI were obtained from Kaplan-Meier estimates for PRS_CAD_ >90^th^ percentile in all individuals (A) and separately in women (B) and men (C). Only individuals without previous event (before recruitment) were included for a median follow-up of 11.0 years (Log-rank test, p-values <0.0001 for A, B and C).

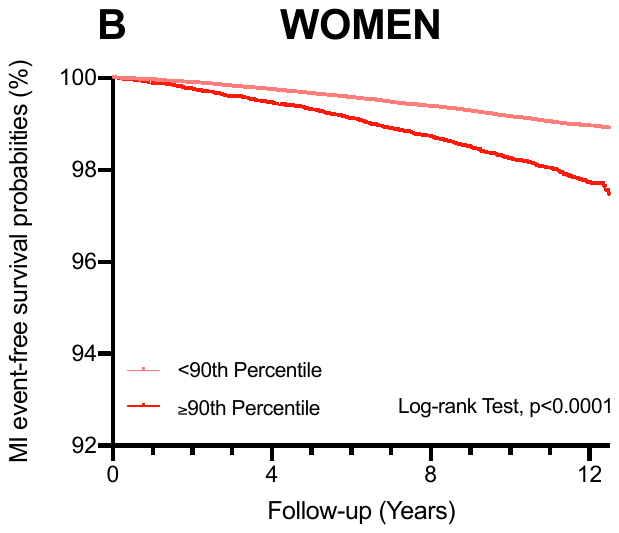

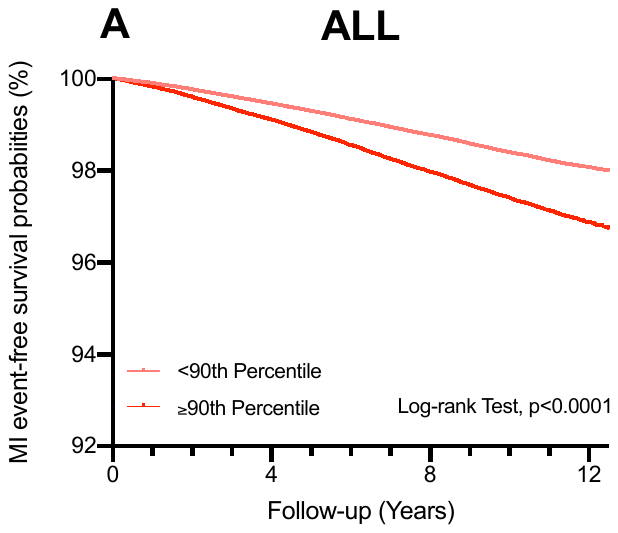

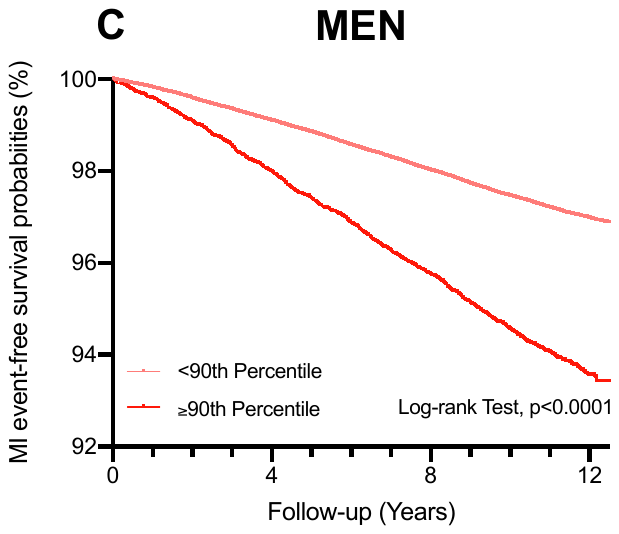

**Supplemental Figure 8: Proportions of participants down and up classified for MI incidence for all ages and in the first quartile of age.** Data are presented for all age groups (A) and for the first quartile of age (B). Number of individuals up and down classified between Cox regressions models including the PCE (standard model) or PCE + PRS_CAD_ (new model) over a period of 10 years.

PRS_CAD_: Polygenic Risk Score for Coronary Artery Disease; PCE: Pooled Cohort Equation.

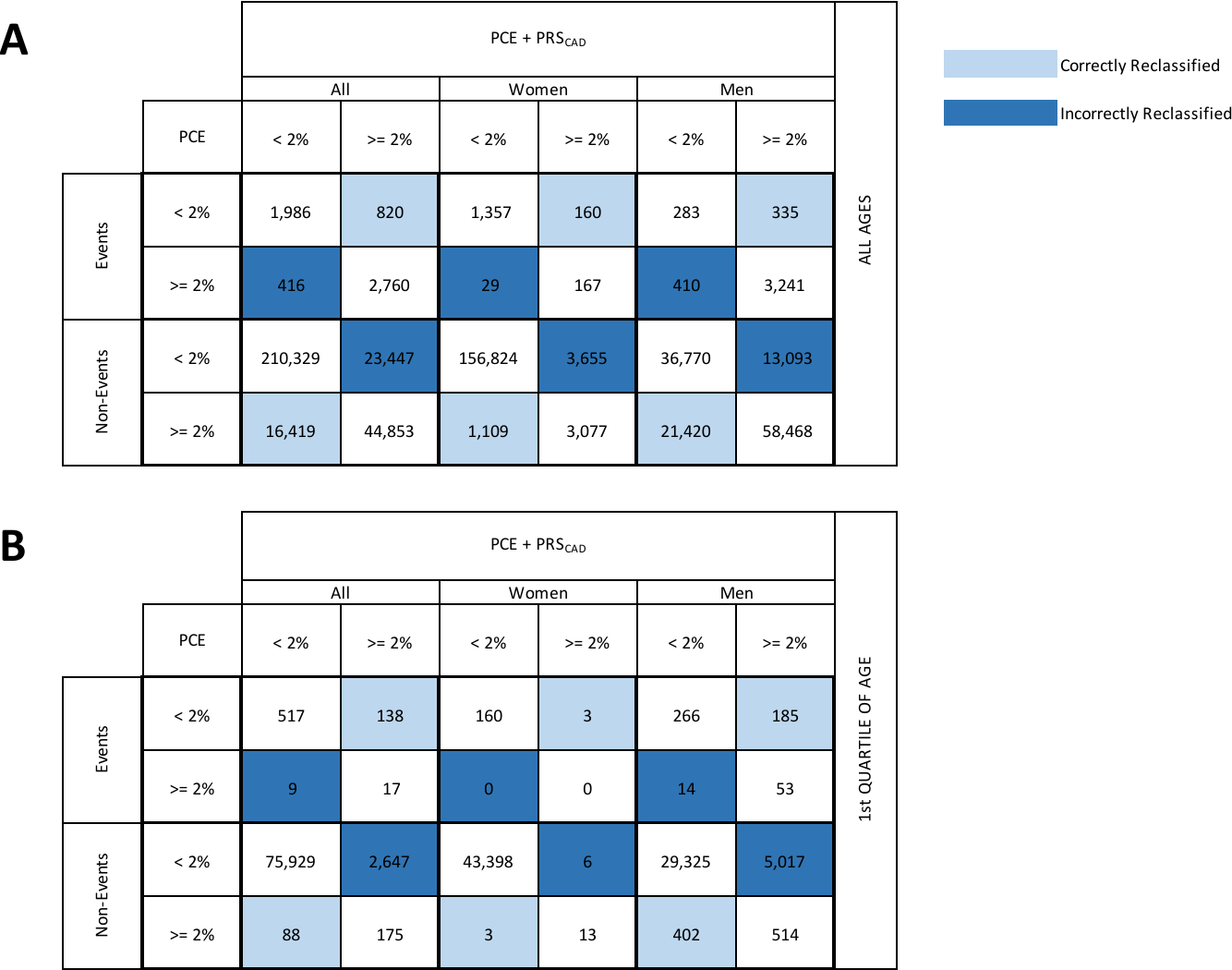

**Supplemental Figure 9: Mortality in individuals at high genetic risk.** Cox regression results were obtained for individuals above different percentile thresholds of PRS_CAD_ (ranging between the 80^th^ and the 97.5^th^ percentile). Hazard ratios (HR) for all-cause mortality and their 95% confidence intervals (95% CI) were compared in all individuals (A), and separately in women (B) and in men (C), for all ages and each quartile of age (Q1 to Q4).

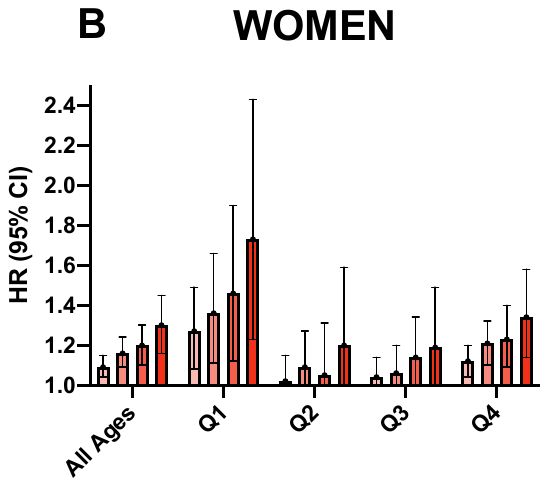

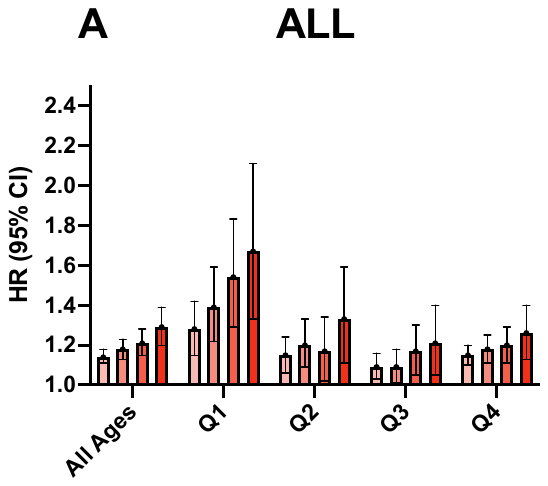

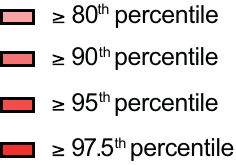

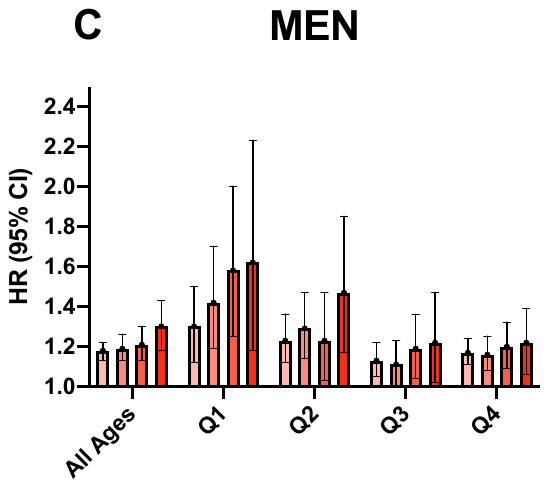

**Supplemental Figure 10: Impact of PRS_CAD_ on all-cause mortality.** Overall survival probabilities curves were obtained from Kaplan-Meier estimates for PRS_CAD_ >90^th^ percentile in all individuals (A) and separately in women (B) and men (C). Only individuals without previous event (before recruitment) were included for a median follow-up of 11.04 years (Log-rank test, p-values <0.0001 for A, B and C).

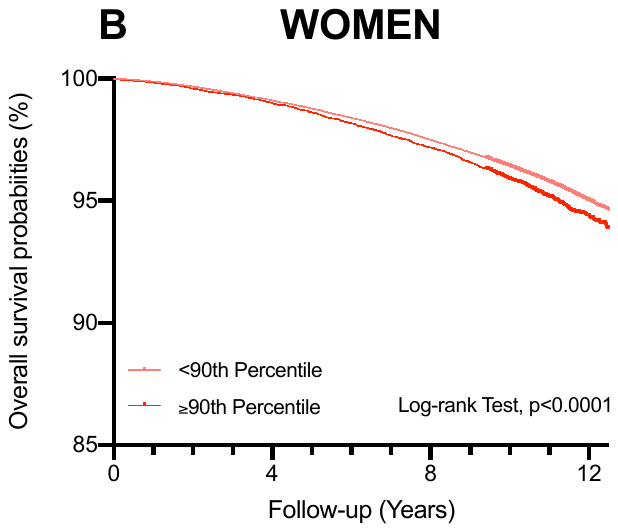

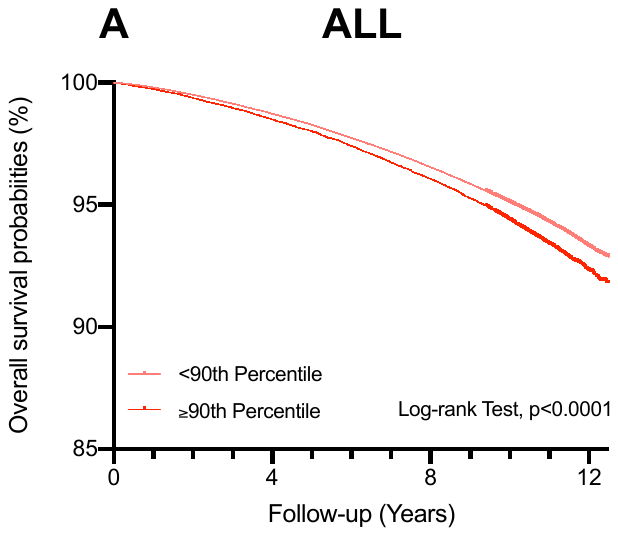

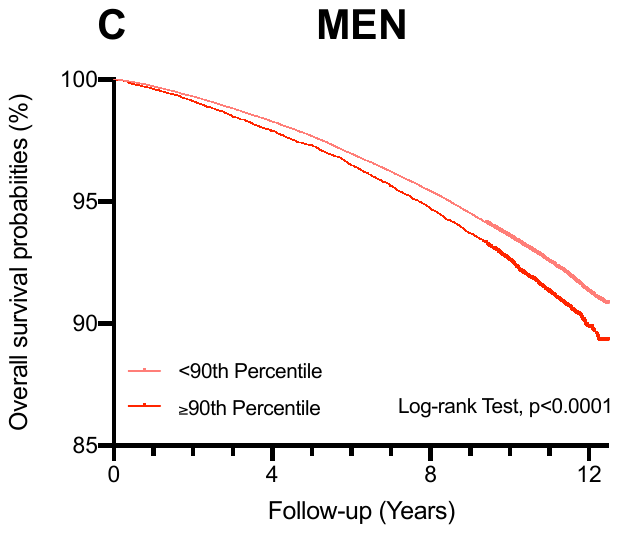

### **SUPPLEMENTAL REFERENCES**

1.         Goff DC, Lloyd-Jones DM, Bennett G, Coady S, D’Agostino RB, Gibbons R, et al. 2013 ACC/AHA Guideline on the Assessment of Cardiovascular Risk. Journal of the American College of Cardiology 2014;63:2935–2959. doi:10.1016/j.jacc.2013.11.005.

2.         Bycroft C, Freeman C, Petkova D, Band G, Elliott LT, Sharp K, et al. The UK Biobank resource with deep phenotyping and genomic data. Nature 2018;562:203–209. doi:10.1038/s41586-018-0579-z.

3.         Vilhjálmsson BJ, Yang J, Finucane HK, Gusev A, Lindström S, Ripke S, et al. Modeling Linkage Disequilibrium Increases Accuracy of Polygenic Risk Scores. The American Journal of Human Genetics 2015;97:576–592. doi:10.1016/j.ajhg.2015.09.001.

4.         Nikpay M, Goel A, Won HH, Hall LM, Willenborg C, Kanoni S, et al. A comprehensive 1000 Genomes-based genome-wide association meta-analysis of coronary artery disease. Nature Genetics 2015;47:1121–1130. doi:10.1038/ng.3396.

5.         Auton A, Abecasis GR, Altshuler DM, Durbin RM, Bentley DR, Chakravarti A, et al. A global reference for human genetic variation. Nature 2015;526:68–74. doi:10.1038/nature15393.

6.         Pencina MJ, D’Agostino RB, Steyerberg EW. Extensions of net reclassification improvement calculations to measure usefulness of new biomarkers. Statistics in Medicine 2011;30:11–21. doi:10.1002/sim.4085.

7.         Kerr KF, Wang Z, Janes H, McClelland RL, Psaty BM, Pepe MS. Net Reclassification Indices for Evaluating Risk Prediction Instruments: A critical review. Epidemiology 2014;25:114–121. doi:10.1097/EDE.0000000000000018.
